# How does the local area deprivation influence life chances for children in poverty in Wales: A record linkage cohort study

**DOI:** 10.1101/2021.06.21.21259276

**Authors:** Amrita Bandyopadhyay, Tony Whiffen, Richard Fry, Sinead Brophy

## Abstract

**Background:** Children growing up in poverty are less likely to achieve in school and more likely to experience mental health problems. This study examined factors in the local area that can help a child overcome the negative impact of poverty.

**Method:** This retrospective cohort study included 159,131 children who lived in Wales and completed their age 16 exams (Key Stage 4 (KS4)) between 2009 and 2016. Free School Meal (FSM) provision was used an indicator of household level deprivation. Area level deprivation was measured by the Welsh Index of Multiple Deprivation 2011. An encrypted unique Anonymous Linking Field was used to link the children with their health and educational records. The outcome variable of ‘Overall doing well’ was comprised of achieved at KS4, no mental health condition, no substance, and alcohol abuse records. Bidirectional logistic regression models were used to investigate the association between local areas deprivation and child’s outcome.

**Results:** 22% of children on FSM were overall doing well compared to 54.9% of non-FSM children. FSM Children who lived in the least deprived areas are significantly more likely to do well (adjusted odds ratio (aOR) -2.20 (1.93-2.51)) than children lived in the most deprived areas. FSM children, living in areas with higher community safety, higher relative income, higher access to services, are more likely to do well than their peers.

**Conclusion:** This study highlights that investing in community development (e.g., safety, back to work schemes, connectivity) helps in child’s education attainment, mental health and reduce risk-taking behaviours.

**What is already known on this subject?:**

1. In Wales, 29.3% of children are living in poverty
2. Welsh Index of Multiple Deprivation (WIMD) 2019 from Welsh Government highlighted the areas in Wales which remained as the topmost deprived areas for more than last 15 years and indicated a lack of social mobility in most of these areas.
3. Growing up in a disadvantaged area increases the risk of adverse health and social outcomes in children’s life.

**What this study adds?:**

1. Children from disadvantaged households are significantly more likely to do well if they are living in least deprived areas than the children from most deprived areas.
2. The findings suggest that investing in community development and local area improvements such as promoting community safety, improving the access to public services and return to work schemes, can also help local children to do well in terms of education, mental health and reducing risk-taking behaviours (alcohol/drug use).

## INTRODUCTION

Latest figures suggest that in 2020, 29.3% of children aged between 0 and 19 are living in poverty (i.e. family income below 60% of the median income) in Wales, which is a 1% rise compare to the previous year.[1] Living in persistent poverty has a detrimental impact on child health, cognitive and behavioural outcomes.[2] Child poverty has caused an unprecedented increase in infant mortality in recent years in the UK.[3] While infant mortality rate in many similar European countries (such as Finland, Sweden) has consistently declined, the rate for the UK has plateaued and started to increase in the past few years.[4] Compared to countries in Western Europe (e.g. France, Germany), UK has the highest mortality rate for children under age 5 years.[5] After a steady fall in the last decade (post-2010), the child poverty rate has also now started to increase in the UK.[3,6] In the post-recession recovery period (i.e. since 2008) inequality increased.[7,8] because of disproportionately slow recovery for low-income families. This is due to real-term cuts in benefits, increasing housing costs and restricted possibilities to improve income from work (e.g. due to salary reductions, freeze in promotions).[9] As a result, relative child poverty is now more prevalent in working families as opposed to workless households.[10] Currently 67% of poor children are living in households where at least one person is working.[11] A report from *End Child Poverty* carried out by Loughborough University has shown that child poverty is disproportionately rising in the UK’s most impoverished areas.[12] The report shows that in some parts of Wales, children from deprived families are six times more likely to be growing up in poverty than their neighbours even if they are living in less deprived areas. The latest report from the Welsh Index of Multiple Deprivation (WIMD) 2019 from Welsh Government highlighted ‘deep-rooted’ deprivation by highlighting the areas in Wales which have remained as the top most deprived areas for more than last 15 years, which indicates a lack of social mobility in most of these areas.[13]

A child growing up in a deprived area implies that they are more likely to be provided with insufficient educational support, lack of recreational space (no safe park or playground) and receive poor quality childcare and health support.[14] This has numerous inevitable long-term consequences such as poorer mental and physical health, lower school achievement, and worse outcomes in adulthood.[2,14–16] Children living in deprived neighbourhoods are less likely to complete high school and achieve higher educational attainment. This creates a significant difference in their earning levels in later life compared to their peers [14]. Local areas with community safety issues often restrict children from after school outdoor activities and increases their sedentary behaviours. This significantly contributes to childhood obesity amongst children living in poor neighbourhoods.[17] Family and area level disadvantageous socio-economic conditions often lead to teenage pregnancy,[18] which is significantly associated with adverse health outcomes and social consequences.[19]

It follows from the above that growing up in a deprived family and local area increase the possibility of a dysfunctional environment which contributes to maladaptive development of children. But, despite this adversity some children beat the odds and achieve better than their peers despite coming from disadvantage.[20] Studies have investigated various factors that can be linked with overcoming odds, such as moving to a more affluent area in early childhood,[21] living in an area with better access to green space,[22] safer community areas so the parents allow and encourage their children to be involved in outdoor physical activity,[23] and neighbourhood safety that enhances collective socialisation.[24,25] Though such evidence is fragmented, it indicates that improvement of the quality, facility and environment of the local area can help the children to build resilience and overcome adversity. Hence it is necessary to develop a holistic understanding of neighbourhoods and prioritise the various aspects of a local area which can help children and their parents to improve their life and overcome poverty.

This study investigates which different socio-economic determinants of a local area are associated with the resilience in children using a linked routine data framework. This study has used the Welsh Index of Multiple Deprivation (WIMD 2011) to identify the concentrations and variations of several domains of deprivation for small areas in Wales and its impact on children. The findings of the study can provide important insights for targeted policy development and intervention.

## METHOD

### Sample

The study population was comprised of children who completed their age 16 exams, Key Stage 4 (KS4), between 2009 and 2016 and had a valid Free School Meal (FSM) record (eligible or not eligible). The children were born or resident of Wales until they completed KS4. The participants were derived by linking Wales Demographic Service Dataset (WDSD) (a Wales-wide administrative register for all individuals with a general practitioner (GP)) and education datasets. The data linkage was performed with the help of an anonymised encrypted linkage key known as Anonymised Lining Filed (ALF) provided by trusted third party in the Secure Anonymised Information Linkage (SAIL) databank platform at Swansea University.[26,27] To enable individuals living in the same household to be anonymously linked, residential anonymised linking fields (RALFs) were created by encrypting individual’s address identifiers for the study period.[28] The children who did not have a continuous residential record (valid RALF) in WDS between age six months and KS4 exam (to ensure they lived in Wales throughout the childhood and we had valid measures of exposures) and primary care record in Welsh Longitudinal General Practice (WLGP) dataset between age 11 and KS4 (when the outcome variable was observed) were excluded from the analysis to ensure the complete data coverage and follow-up period. A detailed consort diagram of the study population is provided in Figure 1.

**Figure 1:**
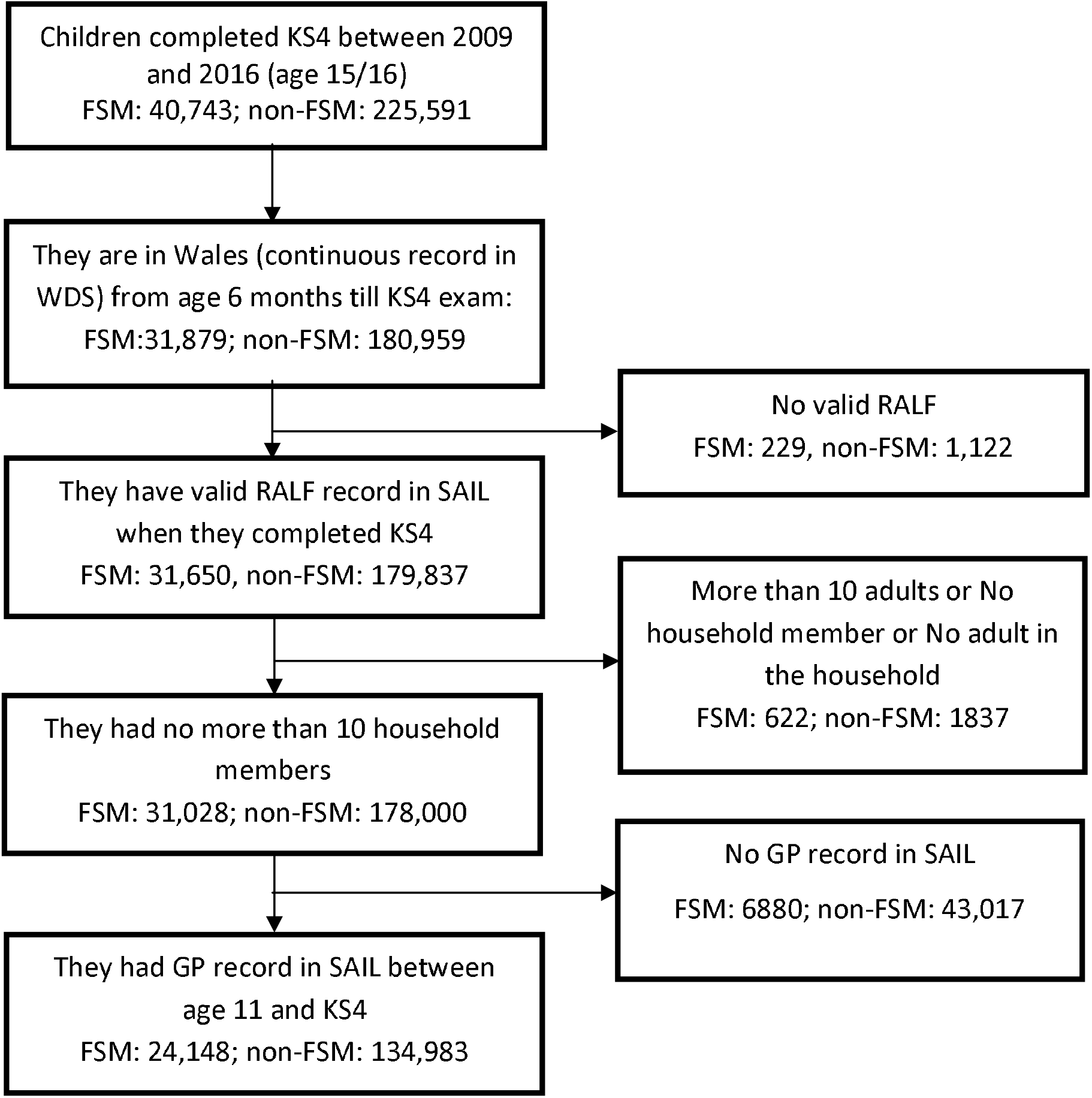
Participants consort diagram (based on Free School Meal eligibility)

### Exposure variables

In this study local area deprivation was measured by using WIMD 2011 [29] which is the official measure of relative deprivation for small areas in Wales. An Individual’s encrypted household identifier RALF was linked to a statistical geography known as a Lower layer Super Output Area (LSOA). Each LSOA is linked with a WIMD rank aggregated into a quintile scale where a lower value denotes greater deprivation. In this study along with overall WIMD rank component scores for WIMD domains such as income, community safety, health, access to services, physical environment, housing, but not education have been considered as main exposure variables.

Individual level socio-economic disadvantage has been measured by FSM incidence.[30] Eligibility for FSM at KS4 was used to determine the household level deprivation of the children.

### Covariates

The other covariates that were included in the study are -living in urban or rural area, number of adults and number of children in the household, living with someone who had depression (diagnosis and/or medication), any household member diagnosed with serious mental illness such as schizophrenia, bipolar disorder (for ICD10 and READ codes see Supplementary material Codes 1), household member who had an alcohol related hospitalisation record (for ICD10 codes see Supplementary material Codes 2), whether the child needs special education support, and year of their KS4 assessment.

### Outcome variable

The outcome variable – ‘Overall doing well’ has been derived based on 4 criteria such as

a) achieved KS4: if they have achieved L2EWM (level 2 English/Welsh Maths– A* to C in 5 GCSE subjects including Maths and English/Welsh)

b) no mental health condition: they have no records of the following conditions -Attention Deficit Hyperactive Disorder, Conduct Disorder, Depression, Serious Mental Illness, Self-harm between age 11 and KS4 assessment

c) no substance abuse: they have no substance misuse record between age 11 and KS4 assessment

d) no alcohol abuse: they have no alcohol related records between age 11 and KS4 assessment

The study population has been linked with their education data to obtain the KS4 record. The mental health, substance misuse and alcohol record were derived from PEDW, WLGP and substance misuse dataset. The ICD10 and RAED codes for all mental health conditions, substance abuse and alcohol are mentioned the Supplementary material Codes 3, 4 & 5. The children who satisfied all the 4 above-mentioned conditions were considered as ‘Overall doing well’. Those who did not satisfy any one of the conditions were considered as ‘Overall, not doing well’.

### Statistical analysis

The analysis has been stratified based on eligibility for FSM. Children who are eligible for FSM are considered as living in high individual level socio-economic deprivation as opposed to those not eligible for FSM (non-FSM). Stepwise bidirectional (forward and backward) logistic regression models have been developed to determine the association between local area deprivation measured by WIMD and Overall doing well amongst the children in Wales. [31] This method determines the best model with minimum Akaike Information Criterion (AIC) and least significant features are excluded at each iteration step. The data preparation including extraction, cleaning and linkage was performed in Structured Query Language (SQL) on IBM DB2 platform and the analyses were performed in the R statistical language version 3.3.2.[32]

### Ethical approval

This study was approved by the SAIL Databank independent Information Governance Review Panel (IGRP) (project number 0916 – WECC Phase 4).

## RESULTS

Characteristics of the study population by family level poverty as assessed using FSM are presented in Table 1. Those receiving FSM were more likely (compared to non-FSM) to live in a single parent household (29.2% compared to 13.1%, respectively), live with 3 or more other children (18.5% compared to 5.8%, respectively) in the same household or to have special educational needs (36.5% compared to 17.6%). They were also more likely to live with a household member who had an alcohol problem (11% compared to 3.8%), depression (63.3% compared to 39.8%), or a serious mental illness (4.6% compared to 1.2%).

**Table 1:**
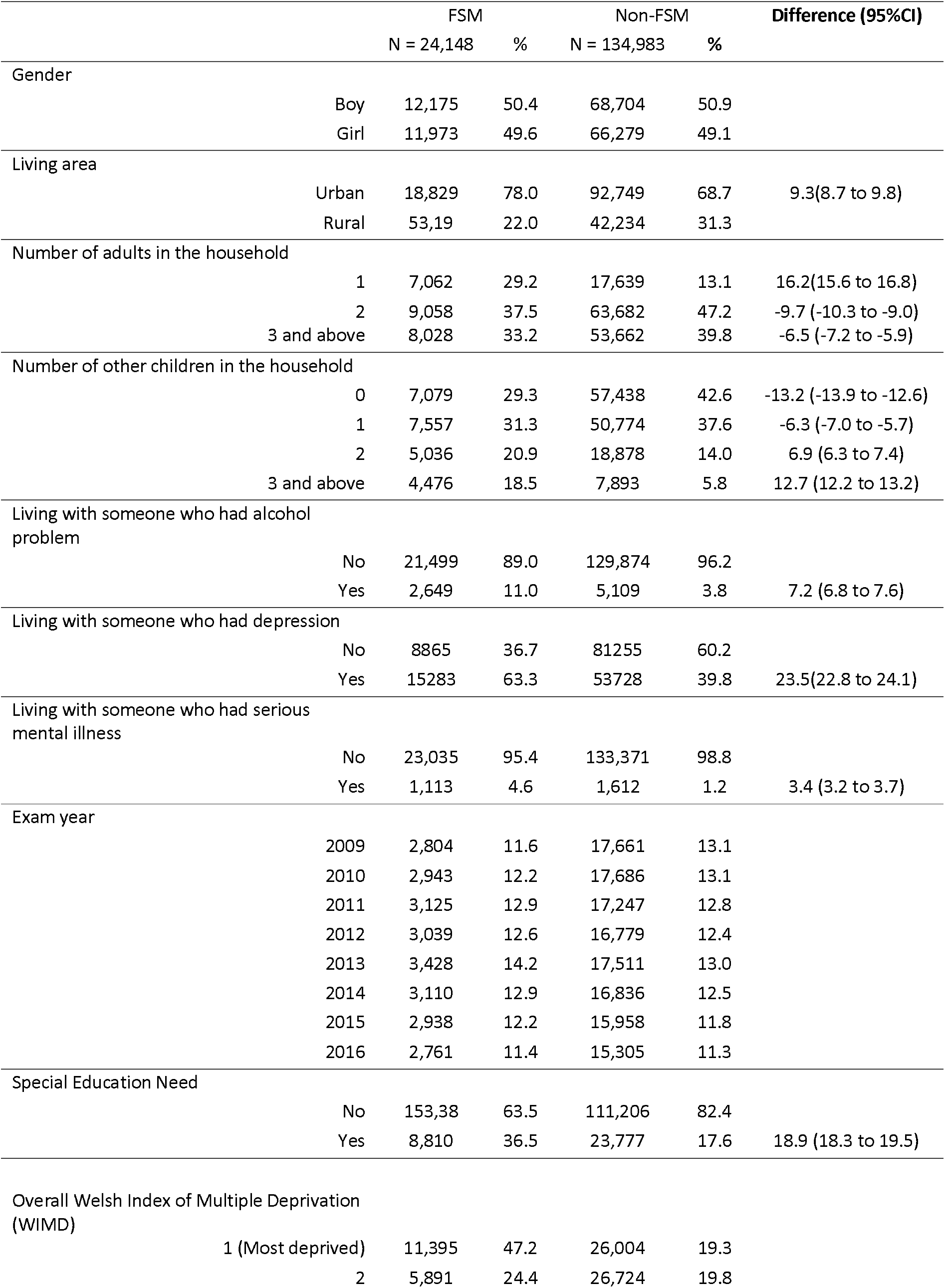
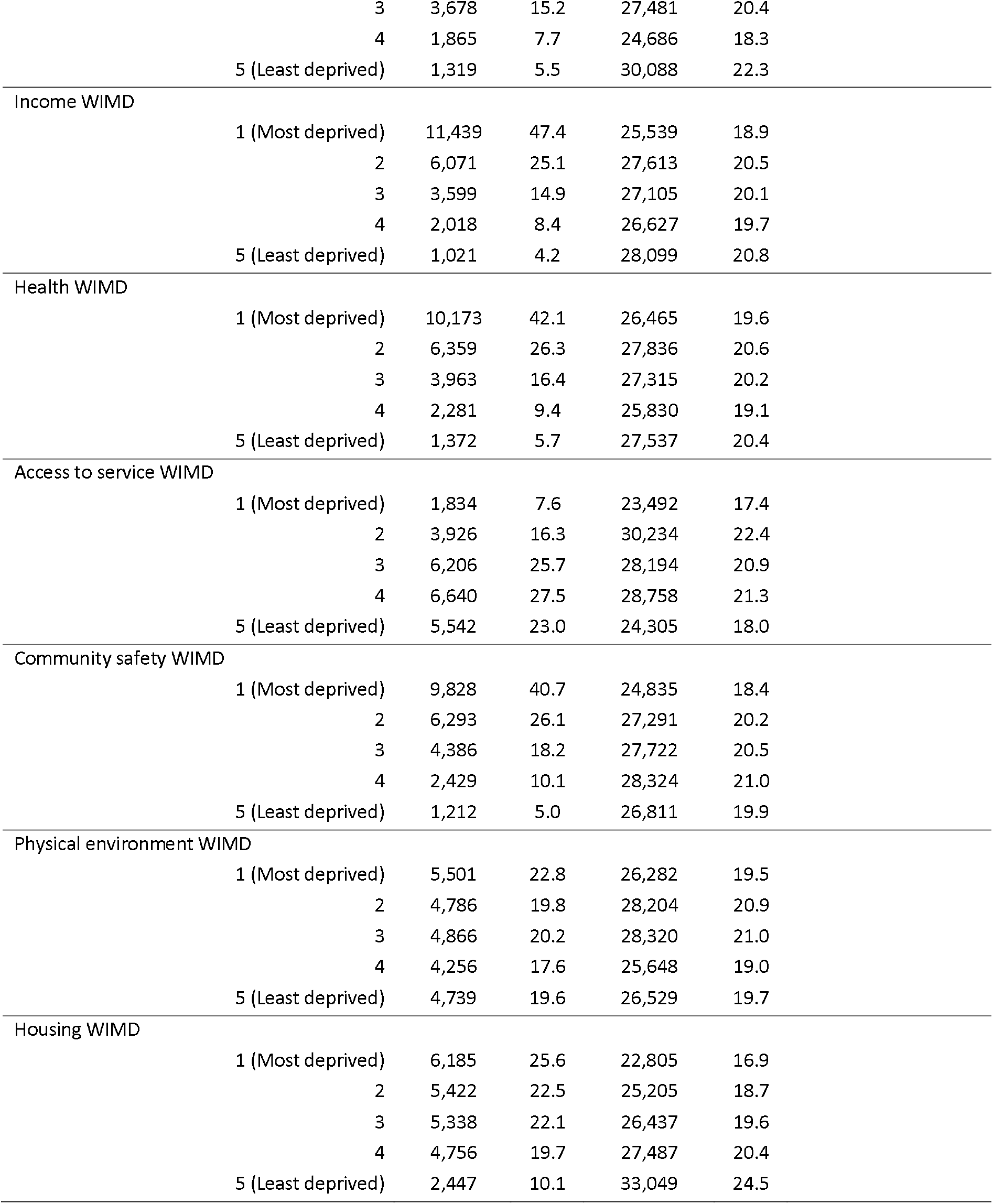
Characteristics of study population by FSM eligibility.

|  |  | FSM |  | Non-FSM |  | Difference (95%CI) |
| --- | --- | --- | --- | --- | --- | --- |
|  |  | N = 24,148 | % | N = 134,983 | % |  |
| Gender |  |  |  |  |  |  |
|  | Boy | 12,175 | 50.4 | 68,704 | 50.9 |  |
|  | Girl | 11,973 | 49.6 | 66,279 | 49.1 |  |
| Living area |  |  |  |  |  |  |
|  | Urban | 18,829 | 78.0 | 92,749 | 68.7 | 9.3(8.7 to 9.8) |
|  | Rural | 53,19 | 22.0 | 42,234 | 31.3 |  |
| Number of adults in the household |  |  |  |  |  |  |
|  | 1 | 7,062 | 29.2 | 17,639 | 13.1 | 16.2(15.6 to 16.8) |
|  | 2 | 9,058 | 37.5 | 63,682 | 47.2 | -9.7 (-10.3 to -9.0) |
|  | 3 and above | 8,028 | 33.2 | 53,662 | 39.8 | -6.5 (-7.2 to -5.9) |
| Number of other children in the household |  |  |  |  |  |  |
|  | 0 | 7,079 | 29.3 | 57,438 | 42.6 | -13.2 (-13.9 to -12.6) |
|  | 1 | 7,557 | 31.3 | 50,774 | 37.6 | -6.3 (-7.0 to -5.7) |
|  | 2 | 5,036 | 20.9 | 18,878 | 14.0 | 6.9 (6.3 to 7.4) |
|  | 3 and above | 4,476 | 18.5 | 7,893 | 5.8 | 12.7 (12.2 to 13.2) |
| Living with someone who had alcohol problem |  |  |  |  |  |  |
|  | No | 21,499 | 89.0 | 129,874 | 96.2 |  |
|  | Yes | 2,649 | 11.0 | 5,109 | 3.8 | 7.2 (6.8 to 7.6) |
| Living with someone who had depression |  |  |  |  |  |  |
|  | No | 8865 | 36.7 | 81255 | 60.2 |  |
|  | Yes | 15283 | 63.3 | 53728 | 39.8 | 23.5(22.8 to 24.1) |
| Living with someone who had serious mental illness |  |  |  |  |  |  |
|  | No | 23,035 | 95.4 | 133,371 | 98.8 |  |
|  | Yes | 1,113 | 4.6 | 1,612 | 1.2 | 3.4 (3.2 to 3.7) |
| Exam year |  |  |  |  |  |  |
|  | 2009 | 2,804 | 11.6 | 17,661 | 13.1 |  |
|  | 2010 | 2,943 | 12.2 | 17,686 | 13.1 |  |
|  | 2011 | 3,125 | 12.9 | 17,247 | 12.8 |  |
|  | 2012 | 3,039 | 12.6 | 16,779 | 12.4 |  |
|  | 2013 | 3,428 | 14.2 | 17,511 | 13.0 |  |
|  | 2014 | 3,110 | 12.9 | 16,836 | 12.5 |  |
|  | 2015 | 2,938 | 12.2 | 15,958 | 11.8 |  |
|  | 2016 | 2,761 | 11.4 | 15,305 | 11.3 |  |
| Special Education Need |  |  |  |  |  |  |
|  | No | 153,38 | 63.5 | 111,206 | 82.4 |  |
|  | Yes | 8,810 | 36.5 | 23,777 | 17.6 | 18.9 (18.3 to 19.5) |
| Overall Welsh Index of Multiple Deprivation (WIMD) |  |  |  |  |  |  |
|  | 1 (Most deprived) | 11,395 | 47.2 | 26,004 | 19.3 |  |
|  | 2 | 5,891 | 24.4 | 26,724 | 19.8 |  |

|  |  |  |  |  |  |
| --- | --- | --- | --- | --- | --- |
|  | 3 | 3,678 | 15.2 | 27,481 | 20.4 |
|  | 4 | 1,865 | 7.7 | 24,686 | 18.3 |
|  | 5 (Least deprived) | 1,319 | 5.5 | 30,088 | 22.3 |
| Income WIMD |  |  |  |  |  |
|  | 1 (Most deprived) | 11,439 | 47.4 | 25,539 | 18.9 |
|  | 2 | 6,071 | 25.1 | 27,613 | 20.5 |
|  | 3 | 3,599 | 14.9 | 27,105 | 20.1 |
|  | 4 | 2,018 | 8.4 | 26,627 | 19.7 |
|  | 5 (Least deprived) | 1,021 | 4.2 | 28,099 | 20.8 |
| Health WIMD |  |  |  |  |  |
|  | 1 (Most deprived) | 10,173 | 42.1 | 26,465 | 19.6 |
|  | 2 | 6,359 | 26.3 | 27,836 | 20.6 |
|  | 3 | 3,963 | 16.4 | 27,315 | 20.2 |
|  | 4 | 2,281 | 9.4 | 25,830 | 19.1 |
|  | 5 (Least deprived) | 1,372 | 5.7 | 27,537 | 20.4 |
| Access to service WIMD |  |  |  |  |  |
|  | 1 (Most deprived) | 1,834 | 7.6 | 23,492 | 17.4 |
|  | 2 | 3,926 | 16.3 | 30,234 | 22.4 |
|  | 3 | 6,206 | 25.7 | 28,194 | 20.9 |
|  | 4 | 6,640 | 27.5 | 28,758 | 21.3 |
|  | 5 (Least deprived) | 5,542 | 23.0 | 24,305 | 18.0 |
| Community safety WIMD |  |  |  |  |  |
|  | 1 (Most deprived) | 9,828 | 40.7 | 24,835 | 18.4 |
|  | 2 | 6,293 | 26.1 | 27,291 | 20.2 |
|  | 3 | 4,386 | 18.2 | 27,722 | 20.5 |
|  | 4 | 2,429 | 10.1 | 28,324 | 21.0 |
|  | 5 (Least deprived) | 1,212 | 5.0 | 26,811 | 19.9 |
| Physical environment WIMD |  |  |  |  |  |
|  | 1 (Most deprived) | 5,501 | 22.8 | 26,282 | 19.5 |
|  | 2 | 4,786 | 19.8 | 28,204 | 20.9 |
|  | 3 | 4,866 | 20.2 | 28,320 | 21.0 |
|  | 4 | 4,256 | 17.6 | 25,648 | 19.0 |
|  | 5 (Least deprived) | 4,739 | 19.6 | 26,529 | 19.7 |
| Housing WIMD |  |  |  |  |  |
|  | 1 (Most deprived) | 6,185 | 25.6 | 22,805 | 16.9 |
|  | 2 | 5,422 | 22.5 | 25,205 | 18.7 |
|  | 3 | 5,338 | 22.1 | 26,437 | 19.6 |
|  | 4 | 4,756 | 19.7 | 27,487 | 20.4 |
|  | 5 (Least deprived) | 2,447 | 10.1 | 33,049 | 24.5 |

### Outcomes for children on FSM

There were 22% FSM children who were classified as ‘Overall doing well’ compared to 54.9% of non-FSM children (difference: 32.9% (95%CI: 32.3% to 33.5%)). The children who were classified as ‘Overall not doing well’ were mainly due to them not achieving KS4 (75.1% of children on FSM) and due to having a mental health condition (11% of FSM children) (see Table 2).

**Table 2:** Breakdown of Overall Doing Well outcome variable.

|  |  | FSM |  | Non-FSM |  | Difference (P |
| --- | --- | --- | --- | --- | --- | --- |
|  |  | N = 24,148 | % | N = 134,983 | % |  |
| Overall Doing Well |  |  |  |  |  |  |
|  | yes | 5311 | 22.0 | 74060 | 54.9 | -32.9 (95%CI: -33.5 to -32.3) |
|  | No | 18837 | 78.0 | 60923 | 45.1 |  |
| KS4 not achieved: |  |  |  |  |  |  |
|  | Achieved | 6005 | 24.9 | 79083 | 58.6 | -33.7- (95%CI:-33.1 to -34.3) |
|  | Not achieved | 18143 | 75.1 | 55900 | 41.4 |  |
| Alcohol record |  |  |  |  |  |  |
|  | No | 22645 | 93.8 | 129441 | 95.9 | -2.1- (95CI: -2.5 to -1.8) |
|  | yes | 1503 | 6.2 | 5542 | 4.1 |  |
| Substance misuse record |  |  |  |  |  |  |
|  | No | 23709 | 98.2 | 134130 | 99.4 | 1.2- (95%CI: -1.4 to -1.0) |
|  | yes | 439 | 1.8 | 853 | 0.6 |  |
| Any mental health condition |  |  |  |  |  |  |
|  | No | 21487 | 89.0 | 128172 | 95.0 | 6.0- (95%CI: -6.4 to -5.6) |
|  | yes | 2661 | 11.0 | 6811 | 5.0 |  |

### Factors associated with ‘Overall doing well’ for children who are on Free School Meals

Children who live in a deprived household but who live in the least deprived areas are significantly more likely to be ‘Overall doing well’ (adjusted odds ratio (aOR) -2.20 (1.93-2.51)) compared to children who are living in the most deprived areas. Living in a household containing less than 3 children, and not living with someone with alcohol problem and depression were also associated with doing well for children living in high individual level socio-economic deprivation (see Table 3). For the FSM children gender, number of adult household member and living with someone who had serious mental illness were not so significant, as a result bidirectional model removed them in the iteration steps.

**Table 3:**
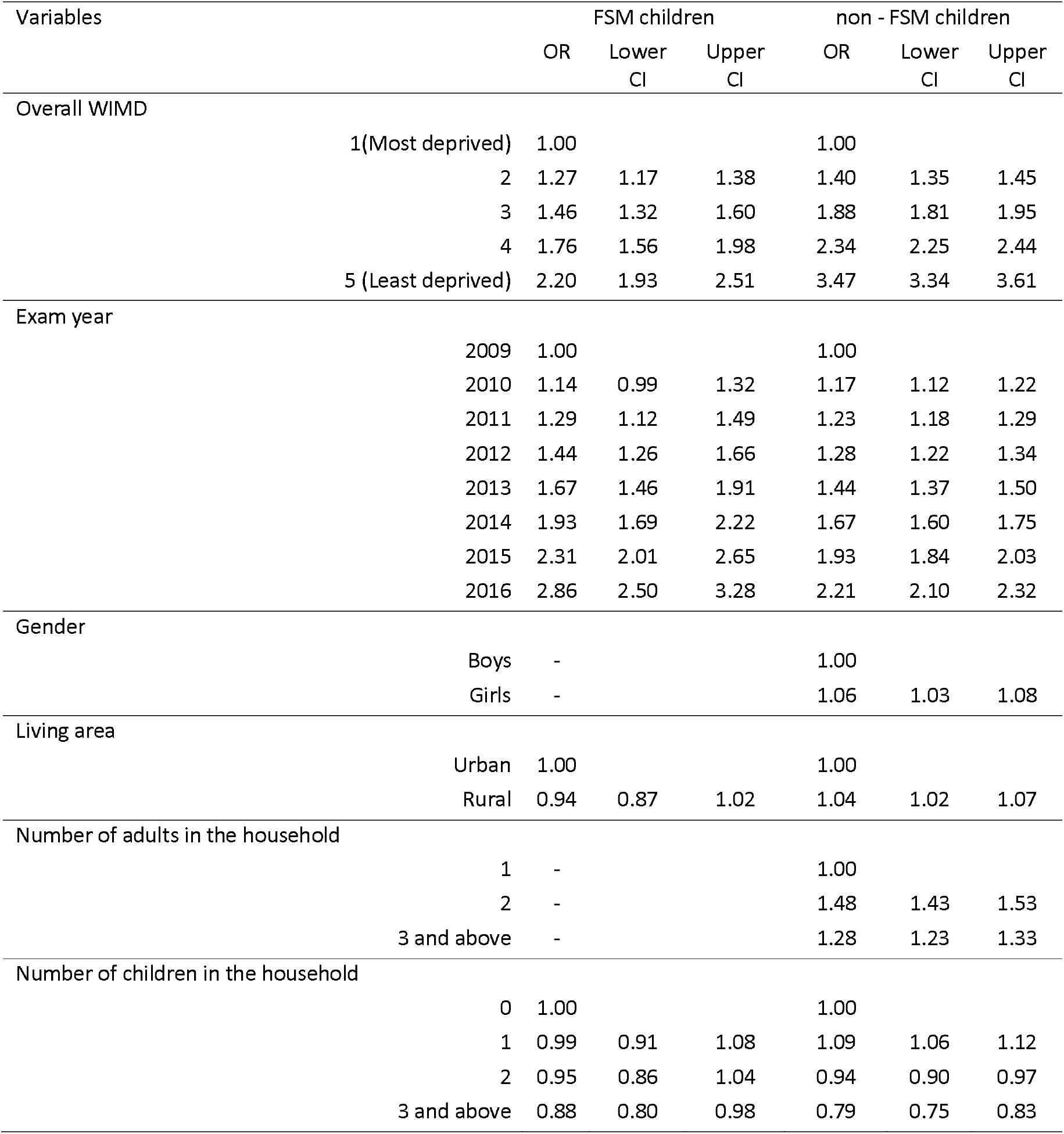
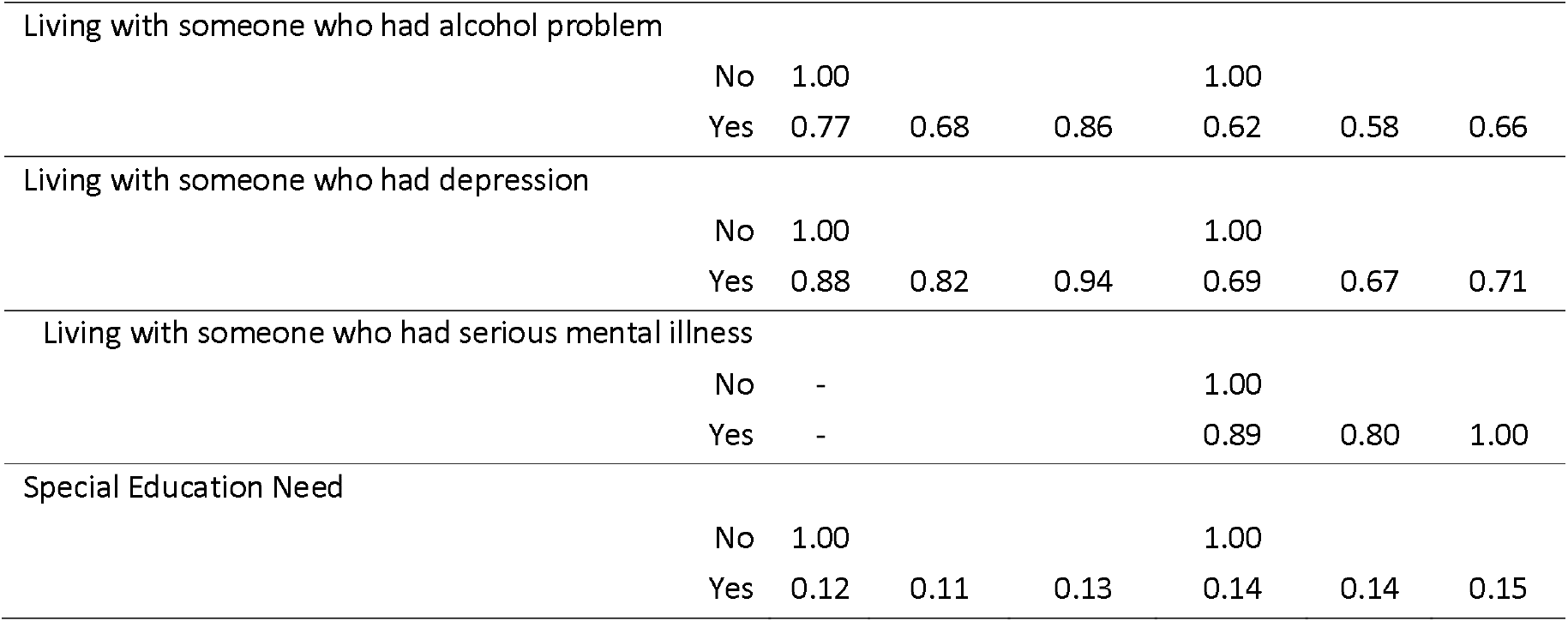
Logistic regression model of the association between overall WIMD and Overall Doing Well for the FSM and non-FSM children.

### Factors associated with ‘Overall doing well’ for children who are not on Free School Meals

Like FSM children, non-FSM children who are living in the least deprived areas are also significantly more likely to do well (adjusted odds ratio (aOR) – 3.47 (3.34-3.61)) compared to children who are living in deprived areas. Non-FSM girls are doing better than boys. The other most significant factors that support these children to do well are -not living in a single adult household, living with 1 other child in the household and not living with someone with alcohol and mental health conditions.

### The impact of different aspects of area on ‘Overall doing well’ for children on Free School Meals

Children who are on FSM and living in deprived areas are significantly less likely to do well than children who are on FSM but living in less derived area (18.32% compared to 34.54%) (see Supplementary material Figure 1). The area components that make most difference to children doing well are higher community safety (1.95 times more likely to do well for FSM children living in the safest areas compared to the least safe areas), higher relative income in the area (e.g. few people on benefits and more people in work,1.61 times more likely to do well if living in highest income area compared to the lowest), and relatively higher access to services (1.26 times more likely to do well if living in highly accessibility compared to those with lowest accessibility). After adjusting for WIMD domains children from urban areas were more likely to do well compared to children from rural areas (see table 4). Area characteristics that did not impact on doing well included general health of people in the area or physical environment (e.g., pollution levels). Supplementary material Figure 2 and 3 graphically depicts the significant indicators that are associated with ‘Overall doing well’ for both FSM and non-FSM children.

**Table 4:**
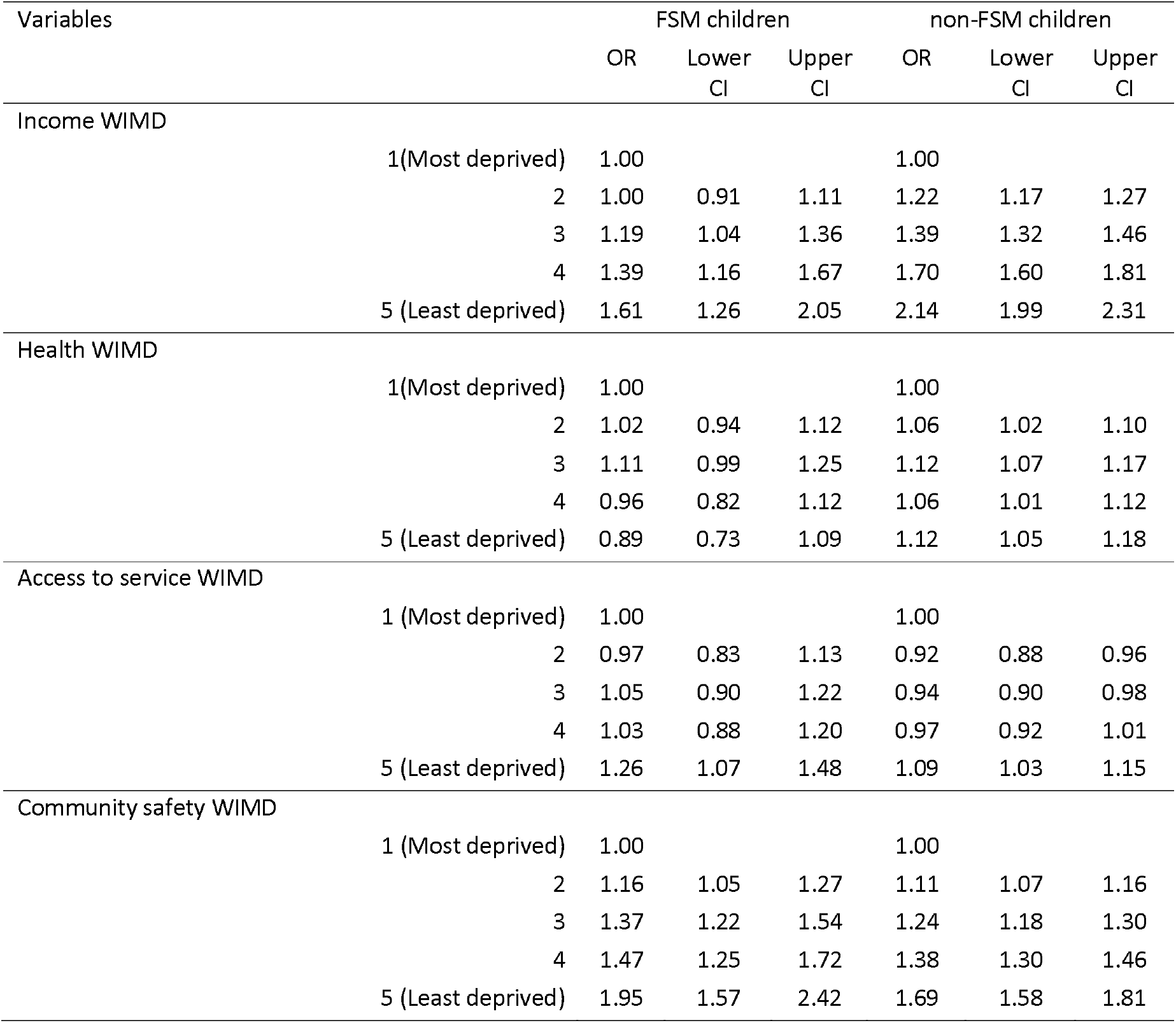
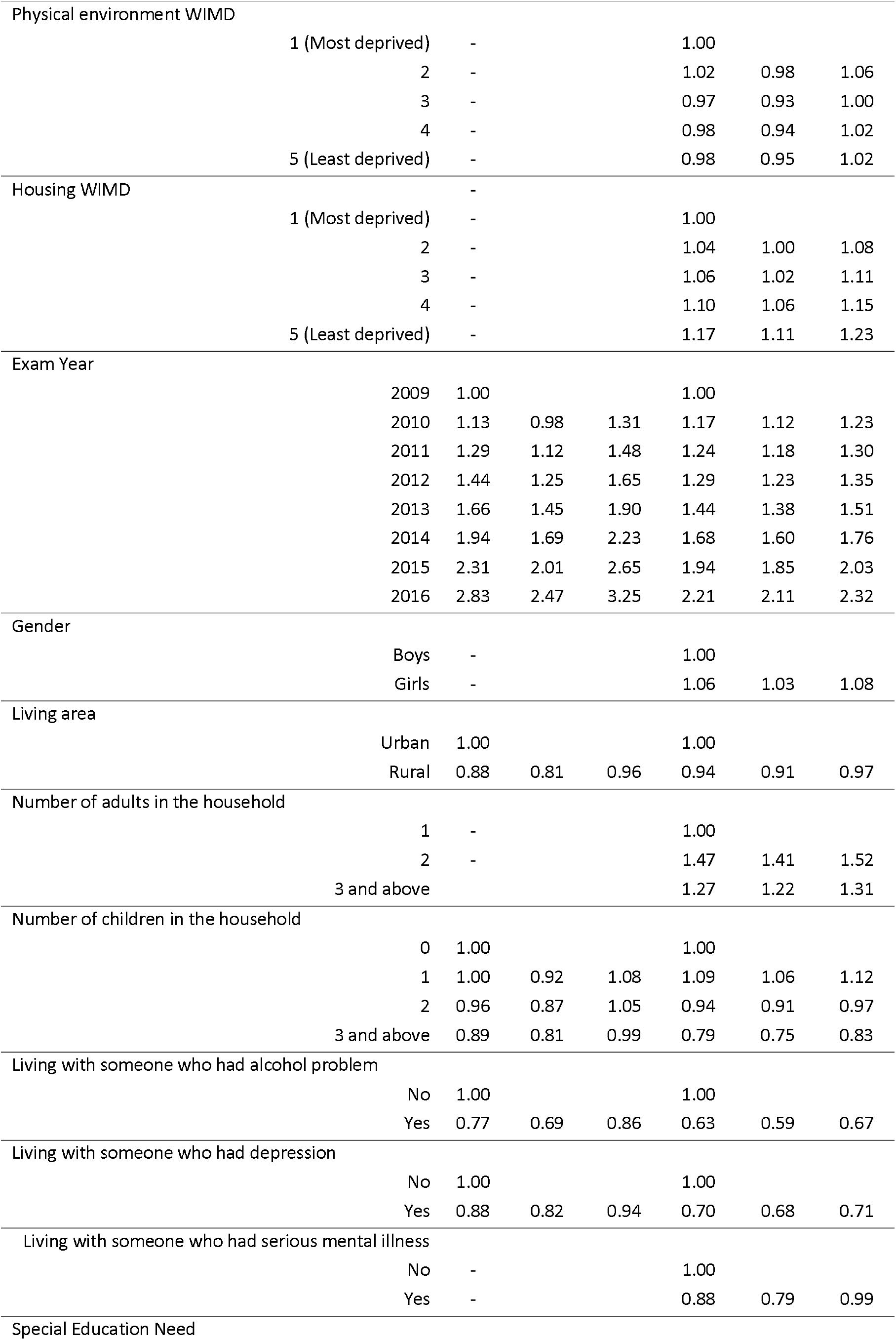
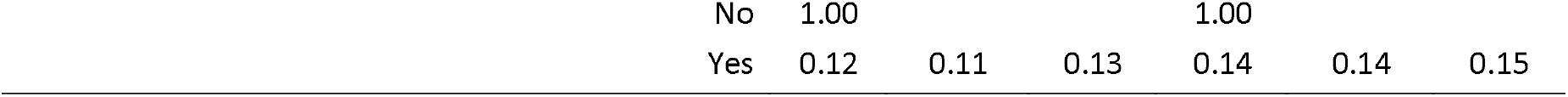
Logistic regression model of the association between WIMD components and Overall Doing Well for the FSM and non-FSM children.

| Variables |  | FSM children |  |  | non-FSM children |  |  |
| --- | --- | --- | --- | --- | --- | --- | --- |
|  |  | OR | Lower<br>CI | Upper<br>CI | OR | Lower<br>CI | Upper<br>CI |
| Income WIMD |  |  |  |  |  |  |  |
|  | 1(Most deprived) | 1.00 |  |  | 1.00 |  |  |
|  | 2 | 1.00 | 0.91 | 1.11 | 1.22 | 1.17 | 1.27 |
|  | 3 | 1.19 | 1.04 | 1.36 | 1.39 | 1.32 | 1.46 |
|  | 4 | 1.39 | 1.16 | 1.67 | 1.70 | 1.60 | 1.81 |
|  | 5 (Least deprived) | 1.61 | 1.26 | 2.05 | 2.14 | 1.99 | 2.31 |
| Health WIMD |  |  |  |  |  |  |  |
|  | 1(Most deprived) | 1.00 |  |  | 1.00 |  |  |
|  | 2 | 1.02 | 0.94 | 1.12 | 1.06 | 1.02 | 1.10 |
|  | 3 | 1.11 | 0.99 | 1.25 | 1.12 | 1.07 | 1.17 |
|  | 4 | 0.96 | 0.82 | 1.12 | 1.06 | 1.01 | 1.12 |
|  | 5 (Least deprived) | 0.89 | 0.73 | 1.09 | 1.12 | 1.05 | 1.18 |
| Access to service WIMD |  |  |  |  |  |  |  |
|  | 1 (Most deprived) | 1.00 |  |  | 1.00 |  |  |
|  | 2 | 0.97 | 0.83 | 1.13 | 0.92 | 0.88 | 0.96 |
|  | 3 | 1.05 | 0.90 | 1.22 | 0.94 | 0.90 | 0.98 |
|  | 4 | 1.03 | 0.88 | 1.20 | 0.97 | 0.92 | 1.01 |
|  | 5 (Least deprived) | 1.26 | 1.07 | 1.48 | 1.09 | 1.03 | 1.15 |
| Community safety WIMD |  |  |  |  |  |  |  |
|  | 1 (Most deprived) | 1.00 |  |  | 1.00 |  |  |
|  | 2 | 1.16 | 1.05 | 1.27 | 1.11 | 1.07 | 1.16 |
|  | 3 | 1.37 | 1.22 | 1.54 | 1.24 | 1.18 | 1.30 |
|  | 4 | 1.47 | 1.25 | 1.72 | 1.38 | 1.30 | 1.46 |
|  | 5 (Least deprived) | 1.95 | 1.57 | 2.42 | 1.69 | 1.58 | 1.81 |
| Physical environment WIMD |  |  |  |  |  |  |  |
|  | 1 (Most deprived) | - |  |  | 1.00 |  |  |
|  | 2 | - |  |  | 1.02 | 0.98 | 1.06 |
|  | 3 | - |  |  | 0.97 | 0.93 | 1.00 |
|  | 4 | - |  |  | 0.98 | 0.94 | 1.02 |
|  | 5 (Least deprived) | - |  |  | 0.98 | 0.95 | 1.02 |
| Housing WIMD |  |  |  |  |  |  |  |
|  | 1 (Most deprived) | - |  |  | 1.00 |  |  |
|  | 2 | - |  |  | 1.04 | 1.00 | 1.08 |
|  | 3 | - |  |  | 1.06 | 1.02 | 1.11 |
|  | 4 | - |  |  | 1.10 | 1.06 | 1.15 |
|  | 5 (Least deprived) | - |  |  | 1.17 | 1.11 | 1.23 |
| Exam Year |  |  |  |  |  |  |  |
|  | 2009 | 1.00 |  |  | 1.00 |  |  |
|  | 2010 | 1.13 | 0.98 | 1.31 | 1.17 | 1.12 | 1.23 |
|  | 2011 | 1.29 | 1.12 | 1.48 | 1.24 | 1.18 | 1.30 |
|  | 2012 | 1.44 | 1.25 | 1.65 | 1.29 | 1.23 | 1.35 |
|  | 2013 | 1.66 | 1.45 | 1.90 | 1.44 | 1.38 | 1.51 |
|  | 2014 | 1.94 | 1.69 | 2.23 | 1.68 | 1.60 | 1.76 |
|  | 2015 | 2.31 | 2.01 | 2.65 | 1.94 | 1.85 | 2.03 |
|  | 2016 | 2.83 | 2.47 | 3.25 | 2.21 | 2.11 | 2.32 |
| Gender |  |  |  |  |  |  |  |
|  | Boys | - |  |  | 1.00 |  |  |
|  | Girls | - |  |  | 1.06 | 1.03 | 1.08 |
| Living area |  |  |  |  |  |  |  |
|  | Urban | 1.00 |  |  | 1.00 |  |  |
|  | Rural | 0.88 | 0.81 | 0.96 | 0.94 | 0.91 | 0.97 |
| Number of adults in the household |  |  |  |  |  |  |  |
|  | 1 | - |  |  | 1.00 |  |  |
|  | 2 | - |  |  | 1.47 | 1.41 | 1.52 |
|  | 3 and above |  |  |  | 1.27 | 1.22 | 1.31 |
| Number of children in the household |  |  |  |  |  |  |  |
|  | 0 | 1.00 |  |  | 1.00 |  |  |
|  | 1 | 1.00 | 0.92 | 1.08 | 1.09 | 1.06 | 1.12 |
|  | 2 | 0.96 | 0.87 | 1.05 | 0.94 | 0.91 | 0.97 |
|  | 3 and above | 0.89 | 0.81 | 0.99 | 0.79 | 0.75 | 0.83 |
| Living with someone who had alcohol problem |  |  |  |  |  |  |  |
|  | No | 1.00 |  |  | 1.00 |  |  |
|  | Yes | 0.77 | 0.69 | 0.86 | 0.63 | 0.59 | 0.67 |
| Living with someone who had depression |  |  |  |  |  |  |  |
|  | No | 1.00 |  |  | 1.00 |  |  |
|  | Yes | 0.88 | 0.82 | 0.94 | 0.70 | 0.68 | 0.71 |
| Living with someone who had serious mental illness |  |  |  |  |  |  |  |
|  | No | - |  |  | 1.00 |  |  |
|  | Yes | - |  |  | 0.88 | 0.79 | 0.99 |
| Special Education Need |  |  |  |  |  |  |  |

|  |  |  |  |  |  |  |
| --- | --- | --- | --- | --- | --- | --- |
| No | 1.00 |  |  | 1.00 |  |  |
| Yes | 0.12 | 0.11 | 0.13 | 0.14 | 0.14 | 0.15 |

## DISCUSSION

This study found that the area in which a person grows up has an important impact on how well they do, especially at school, suggesting neighbourhood effect on education irrespective or parental educational attainment.[33] Previous research suggested a weak association between neighbourhood effects and educational attainment and that family background is more of a factor.[34] However, this study highlights specific aspects of neighbourhood characteristics e.g. community safety, area income and connectivity, which impact on children overcoming the negative aspects of poverty. In terms of community safety, the previous studies have showed that children are more able to undertake outdoor physical activity if they are living in a safer place and this directly contributes to their resilience.[22,23] Other evidence indicates that concerns over community safety are a growing reason for dissatisfaction with green spaces.[35] Residents of deprived areas are more likely to report poorer safety in green spaces and visit less frequently [36] with potential indirect consequences for physical development of children. Living in an area which feels unsafe due to high crime levels, has a detrimental effect on residents in general,[37] hence living in an area with minimal crime risk becomes beneficial for child development. Evidence based measures that improve area safely include; neighbourhood watch, street lighting, CCTV, hotspots policing and alley gating (Crime Reduction Toolkit | College of Policing). In addition, this study also found that good access to services such as public transport, food shops, schools, leisure centres and health services is another important aspect of the local area that helps children who are in poverty to “do well” in their life. There has been evidence that children living in an area with good access to necessary services in day-to-day life has a positive influence on their overall development.[38] An area with good public transport and good social connectivity is an advantageous environment for the children. This might explain why children in rural areas do less well compared to children in urban areas. The Income domain of WIMD reflects the proportion of the people who are living in the area who are claiming income-related benefits and qualitative evidence indicates that poverty also has an effect on children’s experiences at school.[39] This study found that children who are in poverty (indicated by eligibility for FSM) do better when living in an area where fewer people are claiming benefits (e.g., less income-related deprivation in the area). This is also supported by a previous study which shows that if children in poverty have relocated to a less poor areas at an early stage, there is a decrease in the risk of adverse consequences in their later life.[21] If the social norm is to be in employment this may make it also the ‘norm’ for children to remain in education or seek employment.

A limitation of this study is that it uses person-level data to identify possible impact of non-income-based factors on child development and education outcomes. Aside from proxies of child poverty such as FSM eligibility the results indicate the effects of community safety, higher relative income, and access to services in an area on children’s ability to do well. In doing so this study utilises small-area level measures from WIMD which are linked to ONS census geographies. ONS census geographies are designed to maintain best practice is disclosure controls for UK census data and therefore necessarily mask household level variations in WIMD characteristics. This will introduce an ecological inference fallacy where aggregated data were used as a basis for individual to make an inference.[40] The aggregated level data might not be necessarily always a true reflection of an individual; hence this can be a limitation of the study. However, findings highlight the impact of broader area-related factors on child development compared to micro-level household income. In addition, more than 20% of eligible children did not have a full GP record between age 11-16 and so were excluded from the study, these children may have different characteristics of those included in the study and we cannot extrapolate to children who may have moved into Wales or out of Wales in their early teens, and the impact this has on ‘doing well’.

In summary, children who grow up in poverty but are ‘Overall doing well’ as defined by; achieve qualifications at age 16, do not have a mental health diagnosis or substance abuse (including alcohol) problems, are those who live in an area with good community safety, have good public transport and access to services and live in an area where people are employed rather than on benefits. This study highlights that investing in community development and local area improvements such as promoting neighbourhood watch, improving public transport and return to work schemes, would help local children to do well in terms of education, mental health and reducing risk-taking behaviours (alcohol/drug use).

## Supporting information

Supplementary material codes 1

Supplementary material codes 2

Supplementary material codes 3

Supplementary material codes 4

Supplementary material codes 5

## Data Availability

All data have been archived in Secure Annonimised Information Linkage Dtabank (https://saildatabank.com/)

## Funding

This research has been carried out as part of the ADR Wales programme of work. The ADR Wales programme of work is aligned to the priority themes as identified in the Welsh Government’s national strategy: Prosperity for All. ADR Wales brings together data science experts at Swansea University Medical School, staff from the Wales Institute of Social and Economic Research, Data and Methods (WISERD) at Cardiff University and specialist teams within the Welsh Government to develop new evidence which supports Prosperity for All by using the SAIL Databank at Swansea University, to link and analyse anonymised data. ADR Wales is part of the Economic and Social Research Council (part of UK Research and Innovation) funded ADR UK (grant ES/S007393/1).

This work was also supported by the National Centre for Population Health and Well-Being Research (NCPHWR) which is funded by Health and Care Research Wales. This work was supported by Health Data Research UK which receives its funding from HDR UK Ltd (NIWA1) funded by the UK Medical Research Council, Engineering and Physical Sciences Research Council, Economic and Social Research Council, Department of Health and Social Care (England), Chief Scientist Office of the Scottish Government Health and Social Care Directorates, Health and Social Care Research and Development Division (Welsh Government), Public Health Agency (Northern Ireland), British Heart Foundation (BHF) and the Welcome Trust.

This work uses data provided by patients and collected by the NHS as part of their care and support. This study used anonymised data held in the Secure Anonymised Information Linkage (SAIL) Databank. We would like to acknowledge all the data providers who enable SAIL to make anonymised data available for research.

## Contributorship statement

All authors contributed to the study conception and design. Data collection, preparation, and analysis were performed by Amrita Bandyopadhyay. The first draft of the manuscript was written by Amrita Bandyopadhyay. Tony Whiffen, Richard Fry and Sinead Brophy reviewed and edited the drafts. All authors read and approved the final manuscript. Conceptualization: Sinead Brophy and Amrita Bandyopadhyay; Methodology: Amrita Bandyopadhyay and Sinead Brophy; Formal analysis and investigation: Amrita Bandyopadhyay Writing -original draft preparation: Amrita Bandyopadhyay; Writing -review and editing: Tony Whiffen, Richard Fry and Sinead Brophy, Supervision: Sinead Brophy.

## Conflict of interest

The authors declare that they have no conflict of interest.

## Notes

### Competing Interest Statement

The authors have declared no competing interest.

### Funding Statement

This research has been carried out as part of the ADR Wales programme of work. ADR Wales is part of the Economic and Social Research Council (part of UK Research and Innovation) funded ADR UK (grant ES/S007393/1).

### Author Declarations

This study was approved by the SAIL Databank independent Information Governance Review Panel (IGRP) (project number 0916 - WECC Phase 4).

