## Supplementary material codes 1 for "How does the local area deprivation influence life chances for children in poverty in Wales: A record linkage cohort study"

### Table 1: Depression related ICD10 codes

| ICD10 codes | Descriptions |
|---|---|
| F251 | Schizoaffective disorder depressive type |
| F32 | Depressive episode |
| F320 | Mild depressive episode |
| F321 | Moderate depressive episode |
| F322 | Severe depressive episode without psychotic symptoms |
| F323 | Severe depressive episode with psychotic symptoms |
| F328 | Other depressive episodes |
| F329 | Depressive episode, unspecified |
| F33 | Recurrent depressive disorder |
| F330 | Recurrent depressive disorder, current episode mild |
| F331 | Recurrent depressive disorder, current episode moderate |
| F332 | Recurrent depressive disorder, current episode severe without psychotic symptoms |
| F333 | Recurrent depressive disorder, current episode severe with psychotic symptoms |
| F334 | Recurrent depressive disorder, currently in remission |
| F338 | Other recurrent depressive disorders |
| F339 | Recurrent depressive disorder, unspecified |
| F341 | Dysthymia |

### Table 2: Depression related READ codes

| RAED codes | Descriptions |
|---|---|
| 1B17. | Depressed |
| 1B1U. | Symptoms of depression |
| 1BQ.. | Loss of capacity for enjoyment |
| 1BT.. | Depressed mood |
| 1BU.. | Loss of hope for the future |
| 2257 | O/E - depressed |
| E0013 | Presenile dementia with depression |
| E0021 | Senile dementia with depression |
| E112. | Single major depressive episode |
| E1120 | Single major depressive episode, unspecified |
| E1121 | Single major depressive episode, mild |
| E1122 | Single major depressive episode, moderate |
| E1123 | Single major depressive episode, severe, without psychosis |
| E1124 | Single major depressive episode, severe, with psychosis |
| E1125 | Single major depressive episode, partial or unspec remission |
| E1126 | Single major depressive episode, in full remission |
| E112z | Single major depressive episode NOS |
| E113. | Recurrent major depressive episode |
| E1130 | Recurrent major depressive episodes, unspecified |
| E1131 | Recurrent major depressive episodes, mild |

| | |
|---|---|
| E1132 | Recurrent major depressive episodes, moderate |
| E1133 | Recurrent major depressive episodes, severe, no psychosis |
| E1134 | Recurrent major depressive episodes, severe, with psychosis |
| E1135 | Recurrent major depressive episodes,partial/unspec remission |
| E1136 | Recurrent major depressive episodes, in full remission |
| E1137 | Recurrent depression |
| E113z | Recurrent major depressive episode NOS |
| E118. | Seasonal affective disorder |
| E11y2 | Atypical depressive disorder |
| E11z2 | Masked depression |
| E130. | Reactive depressive psychosis |
| E135. | Agitated depression |
| E2003 | Anxiety with depression |
| E204. | Neurotic depression reactive type |
| E291. | Prolonged depressive reaction |
| E2B.. | Depressive disorder NEC |
| E2B0. | Postviral depression |
| E2B1. | Chronic depression |
| Eu204 | [X]Post-schizophrenic depression |
| Eu251 | [X]Schizoaffective disorder, depressive type |
| Eu32. | [X]Depressive episode |
| Eu320 | [X]Mild depressive episode |
| Eu321 | [X]Moderate depressive episode |
| Eu322 | [X]Severe depressive episode without psychotic symptoms |
| Eu323 | [X]Severe depressive episode with psychotic symptoms |
| Eu324 | [X]Mild depression |
| Eu325 | [X]Major depression, mild |
| Eu326 | [X]Major depression, moderately severe |
| Eu327 | [X]Major depression, severe without psychotic symptoms |
| Eu328 | [X]Major depression, severe with psychotic symptoms |
| Eu32y | [X]Other depressive episodes |
| Eu32z | [X]Depressive episode, unspecified |
| Eu33. | [X]Recurrent depressive disorder |
| Eu330 | [X]Recurrent depressive disorder, current episode mild |
| Eu331 | [X]Recurrent depressive disorder, current episode moderate |
| Eu332 | [X]Recurr depress disorder cur epi severe without psyc sympt |
| Eu333 | [X]Recurrent depress disorder cur epi severe with psyc symp |
| Eu334 | [X]Recurrent depressive disorder, currently in remission |
| Eu33y | [X]Other recurrent depressive disorders |
| Eu33z | [X]Recurrent depressive disorder, unspecified |
| Eu341 | [X]Dysthymia |
| Eu412 | [X]Mixed anxiety and depressive disorder |

### Table 3: Depression - medication READ codes

| RAED codes | Descriptions |
|---|---|
| d11.. | CHLORAL HYDRATE |
| d12.. | CLOMETHIAZOLE EDISYLATE [HYPNOTIC] |
| d13.. | *DICHLORALPHENAZONE |
| d14.. | *FLUNITRAZEPAM |
| d15.. | FLURAZEPAM |
| d16.. | LOPRAZOLAM |
| d17.. | LORMETAZEPAM |
| d18.. | NITRAZEPAM |
| d1a.. | TEMAZEPAM [HYPNOTIC] |
| d1b.. | *TRIAZOLAM |
| d1c.. | TRICLOFOS SODIUM |
| d1d.. | ZOPICLONE |
| d1f.. | ZOLPIDEM |
| d1g.. | ZALEPLON |
| d1h.. | MELATONIN |
| d21.. | DIAZEPAM [ANXIOLYTIC] |
| d22.. | ALPRAZOLAM |
| d23.. | BROMAZEPAM |
| d24.. | CHLORDIAZEPOXIDE |
| d25.. | CHLORMEZANONE |
| d26.. | CLOBAZAM |
| d27.. | CLORAZEPATE DIPOTASSIUM |
| d28.. | HYDROXYZINE HCL [ANXIOLYTIC] |
| d29.. | *KETAZOLAM |
| d2a.. | LORAZEPAM [ANXIOLYTIC] |
| d2b.. | *MEDAZEPAM |
| d2c.. | MEPROBAMATE |
| d2d.. | OXAZEPAM |
| d2e.. | *PRAZEPAM |
| d2f.. | BUSPIRONE HYDROCHLORIDE |
| d2g.. | FLUMAZENIL |
| d71.. | AMITRIPTYLINE HYDROCHLORIDE [ANTIDEPRESSANT] |
| d72.. | *BUTRIPTYLINE |
| d73.. | CLOMIPRAMINE HYDROCHLORIDE |
| d74.. | DESIPRAMINE HYDROCHLORIDE |
| d75.. | DOSULEPIN HYDROCHLORIDE |
| d76.. | DOXEPIN |
| d77.. | IMIPRAMINE HYDROCHLORIDE [ANTIDEPRESSANT] |
| d78.. | IPRINDOLE |
| d79.. | LOFEPRAMINE |
| d7a.. | MAPROTILINE HYDROCHLORIDE |
| d7b.. | MIANSERIN HYDROCHLORIDE |

| | |
|---|---|
| d7c.. | NORTRIPTYLINE |
| d7d.. | PROTRIPTYLINE HYDROCHLORIDE |
| d7e.. | TRAZODONE HYDROCHLORIDE |
| d7f.. | TRIMIPRAMINE |
| d7g.. | VILOXAZINE HYDROCHLORIDE |
| d7h.. | AMOXAPINE |
| d81.. | PHENELZINE |
| d83.. | ISOCARBOXAZID |
| d84.. | TRANYLCYPROMINE |
| d85.. | MOCLOBEMIDE |
| d91.. | COMPOUND ANTIDEPRESSANTS A-Z |
| da1.. | FLUPENTIXOL [ANTIDEPRESSANT] |
| da2.. | TRYPTOPHAN |
| da3.. | FLUVOXAMINE MALEATE |
| da4.. | FLUOXETINE HYDROCHLORIDE |
| da5.. | SERTRALINE HYDROCHLORIDE |
| da6.. | PAROXETINE HYDROCHLORIDE |
| da7.. | VENLAFAXINE |
| da9.. | CITALOPRAM |
| daA.. | REBOXETINE |
| daB.. | MIRTAZAPINE |
| daC.. | ESCITALOPRAM |
| daD.. | AGOMELATINE |
| gde.. | DULOXETINE |

#### Table 4: Serious Mental Illness related ICD10 codes

| **ICD10 codes** | **Descriptions** |
|---|---|
| F200 | Paranoid schizophrenia |
| F201 | Hebephrenic schizophrenia |
| F202 | Catatonic schizophrenia |
| F203 | Undifferentiated schizophrenia |
| F204 | Post-schizophrenic depression |
| F205 | Residual schizophrenia |
| F206 | Simple schizophrenia |
| F208 | Other schizophrenia |
| F209 | Schizophreniaunspecified |
| F21X | Schizotypal disorder |
| F220 | Delusional disorder |
| F228 | Other persistent delusional disorders |
| F229 | Persistent delusional disorderunspecified |
| F230 | Acute polymorphic psychot disord without symp of schizoph'a |
| F231 | Acute polymorphic psychot disord with symp of schizophrenia |
| F232 | Acute schizophrenia-like psychotic disorder |

| | |
|---|---|
| F233 | Other acute predominantly delusional psychotic disorders |
| F238 | Other acute and transient psychotic disorders |
| F239 | Acute and transient psychotic disorderunspecified |
| F24X | Induced delusional disorder |
| F250 | Schizoaffective disordermanic type |
| F251 | Schizoaffective disorderdepressive type |
| F252 | Schizoaffective disordermixed type |
| F258 | Other schizoaffective disorders |
| F259 | Schizoaffective disorderunspecified |
| F28X | Other nonorganic psychotic disorders |
| F29X | Unspecified nonorganic psychosis |
| F300 | Hypomania |
| F301 | Mania without psychotic symptoms |
| F302 | Mania with psychotic symptoms |
| F308 | Other manic episodes |
| F309 | Manic episodeunspecified |
| F310 | Bipolar affective disordercurrent episode hypomanic |
| F311 | Bipolar affect disord cur epi manic without psychotic symp |
| F312 | Bipolar affect disorder cur epi manic with psychotic symp |
| F313 | Bipolar affect disorder cur epi mild or moderate depression |
| F314 | Bipolar affect disord cur epi sev depres without psyc symp |
| F315 | Bipolar affect disord cur epi severe depres with psyc symp |
| F316 | Bipolar affective disordercurrent episode mixed |
| F317 | Bipolar affective disordercurrently in remission |
| F318 | Other bipolar affective disorders |
| F319 | Bipolar affective disorderunspecified |
| F323 | Severe depressive episode with psychotic symptoms |
| F333 | Recurrent depress disorder cur epi severe with psyc symp |
| F39X | Unspecified mood [affective] disorder |

### Table 5: Serious mental Illness related READ codes

| **RAED codes** | **Descriptions** |
|---|---|
| E10.. | Schizophrenic disorders |
| E100. | Simple schizophrenia |
| E1000 | Unspecified schizophrenia |
| E1001 | Subchronic schizophrenia |
| E1002 | Chronic schizophrenic |
| E1003 | Acute exacerbation of subchronic schizophrenia |
| E1004 | Acute exacerbation of chronic schizophrenia |
| E1005 | Schizophrenia in remission |
| E100z | Simple schizophrenia NOS |
| E101. | Hebephrenic schizophrenia |
| E1010 | Unspecified hebephrenic schizophrenia |

| | |
|---|---|
| E1011 | Subchronic hebephrenic schizophrenia |
| E1012 | Chronic hebephrenic schizophrenia |
| E1013 | Acute exacerbation of subchronic hebephrenic schizophrenia |
| E1014 | Acute exacerbation of chronic hebephrenic schizophrenia |
| E1015 | Hebephrenic schizophrenia in remission |
| E101z | Hebephrenic schizophrenia NOS |
| E102. | Catatonic schizophrenia |
| E1020 | Unspecified catatonic schizophrenia |
| E1021 | Subchronic catatonic schizophrenia |
| E1022 | Chronic catatonic schizophrenia |
| E1023 | Acute exacerbation of subchronic catatonic schizophrenia |
| E1024 | Acute exacerbation of chronic catatonic schizophrenia |
| E1025 | Catatonic schizophrenia in remission |
| E102z | Catatonic schizophrenia NOS |
| E103. | Paranoid schizophrenia |
| E1030 | Unspecified paranoid schizophrenia |
| E1031 | Subchronic paranoid schizophrenia |
| E1032 | Chronic paranoid schizophrenia |
| E1033 | Acute exacerbation of subchronic paranoid schizophrenia |
| E1034 | Acute exacerbation of chronic paranoid schizophrenia |
| E1035 | Paranoid schizophrenia in remission |
| E103z | Paranoid schizophrenia NOS |
| E104. | Acute schizophrenic episode |
| E105. | Latent schizophrenia |
| E1050 | Unspecified latent schizophrenia |
| E1051 | Subchronic latent schizophrenia |
| E1052 | Chronic latent schizophrenia |
| E1053 | Acute exacerbation of subchronic latent schizophrenia |
| E1054 | Acute exacerbation of chronic latent schizophrenia |
| E1055 | Latent schizophrenia in remission |
| E105z | Latent schizophrenia NOS |
| E106. | Residual schizophrenia |
| E107. | Schizo-affective schizophrenia |
| E1070 | Unspecified schizo-affective schizophrenia |
| E1071 | Subchronic schizo-affective schizophrenia |
| E1072 | Chronic schizo-affective schizophrenia |
| E1073 | Acute exacerbation subchronic schizo-affective schizophrenia |
| E1074 | Acute exacerbation of chronic schizo-affective schizophrenia |
| E1075 | Schizo-affective schizophrenia in remission |
| E107z | Schizo-affective schizophrenia NOS |
| E10y. | Other schizophrenia |
| E10y0 | Atypical schizophrenia |
| E10y1 | Coenesthopathic schizophrenia |
| E10yz | Other schizophrenia NOS |

| | |
|---|---|
| E10z. | Schizophrenia NOS |
| E110. | Manic disorder, single episode |
| E1100 | Single manic episode, unspecified |
| E1101 | Single manic episode, mild |
| E1102 | Single manic episode, moderate |
| E1103 | Single manic episode, severe without mention of psychosis |
| E1104 | Single manic episode, severe, with psychosis |
| E1105 | Single manic episode in partial or unspecified remission |
| E1106 | Single manic episode in full remission |
| E110z | Manic disorder, single episode NOS |
| E111. | Recurrent manic episodes |
| E1110 | Recurrent manic episodes, unspecified |
| E1111 | Recurrent manic episodes, mild |
| E1112 | Recurrent manic episodes, moderate |
| E1113 | Recurrent manic episodes, severe without mention psychosis |
| E1114 | Recurrent manic episodes, severe, with psychosis |
| E1115 | Recurrent manic episodes, partial or unspecified remission |
| E1116 | Recurrent manic episodes, in full remission |
| E111z | Recurrent manic episode NOS |
| E1124 | Single major depressive episode, severe, with psychosis |
| E1134 | Recurrent major depressive episodes, severe, with psychosis |
| E114. | Bipolar affective disorder, currently manic |
| E1140 | Bipolar affective disorder, currently manic, unspecified |
| E1141 | Bipolar affective disorder, currently manic, mild |
| E1142 | Bipolar affective disorder, currently manic, moderate |
| E1143 | Bipolar affect disord, currently manic, severe, no psychosis |
| E1144 | Bipolar affect disord, currently manic,severe with psychosis |
| E1145 | Bipolar affect disord,currently manic, part/unspec remission |
| E1146 | Bipolar affective disorder, currently manic, full remission |
| E114z | Bipolar affective disorder, currently manic, NOS |
| E115. | Bipolar affective disorder, currently depressed |
| E1150 | Bipolar affective disorder, currently depressed, unspecified |
| E1151 | Bipolar affective disorder, currently depressed, mild |
| E1152 | Bipolar affective disorder, currently depressed, moderate |
| E1153 | Bipolar affect disord, now depressed, severe, no psychosis |
| E1154 | Bipolar affect disord, now depressed, severe with psychosis |
| E1155 | Bipolar affect disord, now depressed, part/unspec remission |
| E1156 | Bipolar affective disorder, now depressed, in full remission |
| E115z | Bipolar affective disorder, currently depressed, NOS |
| E116. | Mixed bipolar affective disorder |
| E1160 | Mixed bipolar affective disorder, unspecified |
| E1161 | Mixed bipolar affective disorder, mild |
| E1162 | Mixed bipolar affective disorder, moderate |
| E1163 | Mixed bipolar affective disorder, severe, without psychosis |

| | |
|---|---|
| E1164 | Mixed bipolar affective disorder, severe, with psychosis |
| E1165 | Mixed bipolar affective disorder, partial/unspec remission |
| E1166 | Mixed bipolar affective disorder, in full remission |
| E116z | Mixed bipolar affective disorder, NOS |
| E117. | Unspecified bipolar affective disorder |
| E1170 | Unspecified bipolar affective disorder, unspecified |
| E1171 | Unspecified bipolar affective disorder, mild |
| E1172 | Unspecified bipolar affective disorder, moderate |
| E1173 | Unspecified bipolar affective disorder, severe, no psychosis |
| E1174 | Unspecified bipolar affective disorder,severe with psychosis |
| E1175 | Unspecified bipolar affect disord, partial/unspec remission |
| E1176 | Unspecified bipolar affective disorder, in full remission |
| E117z | Unspecified bipolar affective disorder, NOS |
| E11y. | Other and unspecified manic-depressive psychoses |
| E11y0 | Unspecified manic-depressive psychoses |
| E11y1 | Atypical manic disorder |
| E11y3 | Other mixed manic-depressive psychoses |
| E11yz | Other and unspecified manic-depressive psychoses NOS |
| E11z. | Other and unspecified affective psychoses |
| E11z0 | Unspecified affective psychoses NOS |
| E11zz | Other affective psychosis NOS |
| E12.. | Paranoid states |
| E120. | Simple paranoid state |
| E121. | Chronic paranoid psychosis |
| E122. | Paraphrenia |
| E123. | Shared paranoid disorder |
| E12y. | Other paranoid states |
| E12y0 | Paranoia querulans |
| E12yz | Other paranoid states NOS |
| E12z. | Paranoid psychosis NOS |
| E13.. | Other nonorganic psychoses |
| E130. | Reactive depressive psychosis |
| E131. | Acute hysterical psychosis |
| E132. | Reactive confusion |
| E133. | Acute paranoid reaction |
| E134. | Psychogenic paranoid psychosis |
| E13y. | Other reactive psychoses |
| E13y0 | Psychogenic stupor |
| E13y1 | Brief reactive psychosis |
| E13yz | Other reactive psychoses NOS |
| E13z. | Nonorganic psychosis NOS |
| E2122 | Schizotypal personality |
| Eu2.. | [X]Schizophrenia, schizotypal and delusional disorders |
| Eu20. | [X]Schizophrenia |

| | |
|---|---|
| Eu200 | [X]Paranoid schizophrenia |
| Eu201 | [X]Hebephrenic schizophrenia |
| Eu202 | [X]Catatonic schizophrenia |
| Eu203 | [X]Undifferentiated schizophrenia |
| Eu204 | [X]Post-schizophrenic depression |
| Eu205 | [X]Residual schizophrenia |
| Eu206 | [X]Simple schizophrenia |
| Eu20y | [X]Other schizophrenia |
| Eu20z | [X]Schizophrenia, unspecified |
| Eu21. | [X]Schizotypal disorder |
| Eu22. | [X]Persistent delusional disorders |
| Eu220 | [X]Delusional disorder |
| Eu221 | [X]Delusional misidentification syndrome |
| Eu222 | [X]Cotard syndrome |
| Eu223 | [X]Paranoid state in remission |
| Eu22y | [X]Other persistent delusional disorders |
| Eu22z | [X]Persistent delusional disorder, unspecified |
| Eu23. | [X]Acute and transient psychotic disorders |
| Eu230 | [X]Acute polymorphic psychot disord without symp of schizoph |
| Eu231 | [X]Acute polymorphic psychot disord with symp of schizophren |
| Eu232 | [X]Acute schizophrenia-like psychotic disorder |
| Eu233 | [X]Other acute predominantly delusional psychotic disorders |
| Eu23y | [X]Other acute and transient psychotic disorders |
| Eu23z | [X]Acute and transient psychotic disorder, unspecified |
| Eu24. | [X]Induced delusional disorder |
| Eu25. | [X]Schizoaffective disorders |
| Eu250 | [X]Schizoaffective disorder, manic type |
| Eu251 | [X]Schizoaffective disorder, depressive type |
| Eu252 | [X]Schizoaffective disorder, mixed type |
| Eu25y | [X]Other schizoaffective disorders |
| Eu25z | [X]Schizoaffective disorder, unspecified |
| Eu26. | [X]Nonorganic psychosis in remission |
| Eu2y. | [X]Other nonorganic psychotic disorders |
| Eu2z. | [X]Unspecified nonorganic psychosis |
| Eu30. | [X]Manic episode |
| Eu300 | [X]Hypomania |
| Eu301 | [X]Mania without psychotic symptoms |
| Eu302 | [X]Mania with psychotic symptoms |
| Eu30y | [X]Other manic episodes |
| Eu30z | [X]Manic episode, unspecified |
| Eu31. | [X]Bipolar affective disorder |
| Eu310 | [X]Bipolar affective disorder, current episode hypomanic |
| Eu311 | [X]Bipolar affect disorder cur epi manic wout psychotic symp |
| Eu312 | [X]Bipolar affect disorder cur epi manic with psychotic symp |

| | |
|---|---|
| Eu313 | [X]Bipolar affect disorder cur epi mild or moderate depressn |
| Eu314 | [X]Bipol aff disord, curr epis sev depress, no psychot symp |
| Eu315 | [X]Bipolar affect dis cur epi severe depres with psyc symp |
| Eu316 | [X]Bipolar affective disorder, current episode mixed |
| Eu317 | [X]Bipolar affective disorder, currently in remission |
| Eu318 | [X]Bipolar affective disorder type I |
| Eu319 | [X]Bipolar affective disorder type II |
| Eu31y | [X]Other bipolar affective disorders |
| Eu31z | [X]Bipolar affective disorder, unspecified |
| Eu323 | [X]Severe depressive episode with psychotic symptoms |
| Eu333 | [X]Recurrent depress disorder cur epi severe with psyc symp |
