## Supplementary material codes 2 for "How does the local area deprivation influence life chances for children in poverty in Wales: A record linkage cohort study"

### Table 1: Alcohol related ICD10 codes

| ICD10 Codes | Descriptions |
|---|---|
| E244 | Alcohol-induced pseudo-Cushing's syndrome |
| E512 | Wernicke's encephalopathy |
| F10 | Mental and behavioural disorders due to use of alcohol |
| F100 | Mental and behavioural disorders due to use of alcohol |
| F101 | Mental and behavioural disorders due to use of alcohol |
| F102 | Mental and behavioural disorders due to use of alcohol |
| F103 | Mental and behavioural disorders due to use of alcohol |
| F104 | Mental and behavioural disorders due to use of alcohol |
| F105 | Mental and behavioural disorders due to use of alcohol |
| F106 | Mental and behavioural disorders due to use of alcohol |
| F107 | Mental and behavioural disorders due to use of alcohol |
| F108 | Mental and behavioural disorders due to use of alcohol |
| F109 | Mental and behavioural disorders due to use of alcohol |
| G312 | Degeneration of nervous system due to alcohol |
| G621 | Alcoholic polyneuropathy |
| G721 | Alcoholic myopathy |
| I426 | Alcoholic cardiomyopathy |
| K292 | Alcoholic gastritis |
| K70 | Alcoholic liver disease |
| K700 | Alcoholic fatty liver |
| K701 | Alcoholic hepatitis |
| K702 | Alcoholic fibrosis and sclerosis of liver |
| K703 | Alcoholic cirrhosis of liver |
| K704 | Alcoholic hepatic failure |
| K709 | Alcoholic liver disease, unspecified |
| K852 | Alcohol-induced acute pancreatitis |
| K860 | Alcohol-induced chronic pancreatitis |
| O354 | Maternal care for (suspected) damage to fetus from alcohol |
| R780 | Finding of alcohol in blood |
| T51 | Toxic effect of alcohol |
| T510 | Toxic effect: Ethanol |
| T511 | Toxic effect: Methanol |
| T512 | Toxic effect: 2-Propanol |
| T513 | Toxic effect: Fusel oil |
| T518 | Toxic effect: Other alcohols |
| T519 | Toxic effect: Alcohol, unspecified |
| X45 | Accidental poisoning by and exposure to alcohol |
| X450 | Accidental poisoning by and exposure to alcohol |
| X451 | Accidental poisoning by and exposure to alcohol |
| X452 | Accidental poisoning by and exposure to alcohol |
| X453 | Accidental poisoning by and exposure to alcohol |
| X454 | Accidental poisoning by and exposure to alcohol |

| | |
|---|---|
| X455 | Accidental poisoning by and exposure to alcohol |
| X456 | Accidental poisoning by and exposure to alcohol |
| X457 | Accidental poisoning by and exposure to alcohol |
| X458 | Accidental poisoning by and exposure to alcohol |
| X459 | Accidental poisoning by and exposure to alcohol |
| X65 | Intentional self-poisoning by and exposure to alcohol |
| X650 | Intentional self-poisoning by and exposure to alcohol |
| X651 | Intentional self-poisoning by and exposure to alcohol |
| X652 | Intentional self-poisoning by and exposure to alcohol |
| X653 | Intentional self-poisoning by and exposure to alcohol |
| X654 | Intentional self-poisoning by and exposure to alcohol |
| X655 | Intentional self-poisoning by and exposure to alcohol |
| X656 | Intentional self-poisoning by and exposure to alcohol |
| X657 | Intentional self-poisoning by and exposure to alcohol |
| X658 | Intentional self-poisoning by and exposure to alcohol |
| X659 | Intentional self-poisoning by and exposure to alcohol |
| Y15 | Poisoning by and exposure to alcohol, undetermined intent |
| Y150 | Poisoning by and exposure to alcohol, undetermined intent |
| Y151 | Poisoning by and exposure to alcohol, undetermined intent |
| Y152 | Poisoning by and exposure to alcohol, undetermined intent |
| Y153 | Poisoning by and exposure to alcohol, undetermined intent |
| Y154 | Poisoning by and exposure to alcohol, undetermined intent |
| Y155 | Poisoning by and exposure to alcohol, undetermined intent |
| Y156 | Poisoning by and exposure to alcohol, undetermined intent |
| Y157 | Poisoning by and exposure to alcohol, undetermined intent |
| Y158 | Poisoning by and exposure to alcohol, undetermined intent |
| Y159 | Poisoning by and exposure to alcohol, undetermined intent |
| Y573 | Alcohol deterrents |
| Y900 | Blood alcohol level of less than 20 mg/100 ml |
| Y901 | Blood alcohol level of 20-39 mg/100 ml |
| Y902 | Blood alcohol level of 40-59 mg/100 ml |
| Y903 | Blood alcohol level of 60-79 mg/100 ml |
| Y904 | Blood alcohol level of 80-99 mg/100 ml |
| Y905 | Blood alcohol level of 100-119 mg/100 ml |
| Y906 | Blood alcohol level of 120-199 mg/100 ml |
| Y907 | Blood alcohol level of 200-239 mg/100 ml |
| Y908 | Blood alcohol level of 240 mg/100 ml or more |
| Y909 | Presence of alcohol in blood, level not specified |
| Y910 | Mild alcohol intoxication |
| Y911 | Moderate alcohol intoxication |
| Y912 | Severe alcohol intoxication |
| Y913 | Very severe alcohol intoxication |
| Y919 | Alcohol involvement, not otherwise specified |
| Z502 | Alcohol rehabilitation |

| | |
|---|---|
| Z714 | Alcohol abuse counselling and surveillance |
| Z721 | Alcohol use |
