## Supplementary material codes 3 for "How does the local area deprivation influence life chances for children in poverty in Wales: A record linkage cohort study"

### Table 1: ADHD related READ codes

| READ codes | Descriptions |
|---|---|
| 6A61. | ADHD annual review |
| 8BPT. | Drug therapy for ADHD |
| 8BPT0 | Stimulant drug therapy for ADHD |
| 8BPT1 | Non-stimulant drug therapy for ADHD |
| 9Ngp. | On drug ther ADHD (attention deficit hyperactivity disorder) |
| 9Ngp0 | On stim drug ther ADHD (attention def hyperactivity disordr) |
| 9Ngp1 | On non-stimulant drug therapy for ADHD |
| 9Ol8. | ADHD monitoring invitation first letter |
| 9Ol9. | ADHD monitoring invitation second letter |
| 9OlA. | ADHD monitoring invitation third letter |
| E2E.. | Childhood hyperkinetic syndrome |
| E2E0. | Child attention deficit disorder |
| E2E00 | Attention deficit without hyperactivity |
| E2E01 | Attention deficit with hyperactivity |
| E2E0z | Child attention deficit disorder NOS |
| E2E2. | Hyperkinetic conduct disorder |
| E2Ez. | Hyperkinetic syndrome NOS |
| Eu90. | [X]Hyperkinetic disorders |
| Eu900 | [X]Disturbance activity/attntn |
| Eu901 | [X]Hyperkinetic conduct disord |
| Eu90y | [X]Oth hyperkinetic disorders |
| Eu90z | [X]Hyperkinetic disorder, unsp |
| Eu9y7 | [X]Attention deficit disorder |
| dc1.. | DEXAMFETAMINE SULPHATE |
| dc11. | *DEXEDRINE 5mg tablets |
| dc1v. | DEXAMFET SULF 1mg/mL oral soln |
| dc1w. | DEXAMFETAMINE SULPH 5mg tabs |
| dc1x. | *DEXAMPHETAMINE 7.5mg m/r caps |
| dc1y. | *DEXAMPHETAMINE 12.5mg caps |
| dc1z. | *DEXAMPHETAMINE 20mg m/r caps |
| dw... | DRUGS USED TO TREAT HYPERACTIVITY DISORDERS |
| dw1.. | METHYLPHENIDATE |
| dw11. | METHYLPHENIDATE HCL 10mg tabs |
| dw12. | RITALIN 10mg tablets |
| dw13. | *EQUASYM 5mg tablets |
| dw14. | *EQUASYM 20mg tablets |
| dw15. | *EQUASYM 10mg tablets |
| dw16. | EQUASYM XL 20mg m/r capsules |
| dw17. | CONCERTA XL 18mg m/r tablets |
| dw18. | CONCERTA XL 36mg m/r tablets |
| dw1C. | EQUASYM XL 10mg m/r capsules |
| dw1D. | EQUASYM XL 30mg m/r capsules |

| | |
|---|---|
| dw1E. | MEDIKINET XL 10mg m/r capsules |
| dw1F. | MEDIKINET XL 20mg m/r capsules |
| dw1G. | MEDIKINET XL 30mg m/r capsules |
| dw1H. | MEDIKINET XL 40mg m/r capsules |
| dw1I. | CONCERTA XL 27mg m/r tablets |
| dw1J. | MEDIKINET 5mg tablets |
| dw1K. | MEDIKINET 10mg tablets |
| dw1L. | MEDIKINET 20mg tablets |
| dw1M. | MEDIKINET XL 5mg m/r capsules |
| dw1N. | MEDIKINET XL 50mg m/r capsules |
| dw1O. | MEDIKINET XL 60mg m/r capsules |
| dw1U. | CONCERTA XL 54mg m/r tablets |
| dw1n. | METHYLPHENIDATE 54mg m/r tabs |
| dw1o. | METHYLPHENIDATE 50mg m/r caps |
| dw1p. | METHYLPHENIDATE 60mg m/r caps |
| dw1q. | METHYLPHENIDATE 5mg m/r caps |
| dw1r. | METHYLPHENIDATE 27mg m/r tabs |
| dw1s. | METHYLPHENIDATE 40mg m/r caps |
| dw1t. | METHYLPHENIDATE 10mg m/r caps |
| dw1u. | METHYLPHENIDATE 30mg m/r caps |
| dw1v. | METHYLPHENIDATE 36mg m/r tabs |
| dw1w. | METHYLPHENIDATE 18mg m/r tabs |
| dw1x. | METHYLPHENIDATE 20mg m/r caps |
| dw1y. | METHYLPHENIDATE HCL 5mg tabs |
| dw1z. | METHYLPHENIDATE HCL 20mg tabs |
| dw2.. | ATOMOXETINE |
| dw21. | STRATTERA 10mg capsules |
| dw22. | STRATTERA 18mg capsules |
| dw23. | STRATTERA 25mg capsules |
| dw24. | STRATTERA 40mg capsules |
| dw25. | STRATTERA 60mg capsules |
| dw26. | STRATTERA 80mg capsules |
| dw27. | STRATTERA 100mg capsules |
| dw28. | STRATTERA 4mg/mL oral solution |
| dw2s. | ATOMOXETINE 4mg/mL oral soln |
| dw2t. | ATOMOXETINE 100mg capsules |
| dw2u. | ATOMOXETINE 80mg capsules |
| dw2v. | ATOMOXETINE 60mg capsules |
| dw2w. | ATOMOXETINE 40mg capsules |
| dw2x. | ATOMOXETINE 25mg capsules |
| dw2y. | ATOMOXETINE 18mg capsules |
| dw2z. | ATOMOXETINE 10mg capsules |
| dw3.. | LISDEXAMFETAMINE |
| dw3x. | LISDEXAMFETAMINE DIM 70mg caps |

| | |
|---|---|
| dw3y. | LISDEXAMFETAMINE DIM 50mg caps |
| dw3z. | LISDEXAMFETAMINE DIM 30mg caps |

#### Table 2: Conduct Disorder related READ codes

| READ codes | Descriptions |
|---|---|
| E2731 | Head-banging |
| E293z | Adjustment reaction with predominant disturbance conduct NOS |
| E2C.. | Disturbance of conduct NEC |
| E2C0. | Aggressive unsocial conduct disorder |
| E2C00 | Aggressive outburst |
| E2C01 | Anger reaction |
| E2C0z | Aggressive unsocial conduct disorder NOS |
| E2C1. | Nonaggressive unsocial conduct disorder |
| E2C10 | Unsocial childhood truancy |
| E2C11 | Solitary stealing |
| E2C12 | Tantrums |
| E2C1z | Nonaggressive unsocial conduct disorder NOS |
| E2C2. | Socialised conduct disorder |
| E2C2z | Socialised conduct disorder NOS |
| E2C4z | Mixed disturbance of conduct and emotion NOS |
| E2Cy. | Other conduct disturbances |
| E2Cyz | Other conduct disturbances NOS |
| E2Cz. | Unspecified disturbance of conduct |
| E2Czz | Disturbance of conduct NOS |
| E2Dy0 | Childhood and adolescent oppositional disorder |
| E2E2. | Hyperkinetic conduct disorder |
| Eu9.. | [X]Behavioural/emotional disords onset childhood/adolescence |
| Eu901 | [X]Hyperkinetic conduct disorder |
| Eu91. | [X]Conduct disorders |
| Eu910 | [X]Conduct disorder confined to the family context |
| Eu911 | [X]Unsocialized conduct disorder |
| Eu912 | [X]Socialized conduct disorder |
| Eu913 | [X]Oppositional defiant disorder |
| Eu91y | [X]Other conduct disorders |
| Eu91z | [X]Conduct disorder, unspecified |
| Eu92. | [X]Mixed disorders of conduct and emotions |
| Eu920 | [X]Depressive conduct disorder |
| Eu92y | [X]Other mixed disorders of conduct and emotions |
| Eu92z | [X]Mixed disorder of conduct and emotions, unspecified |
| Eu941 | [X]Reactive attachment disorder of childhood |

**Table 3: Self harm related ICD10 codes**

| ICD10 codes | Descriptions |
|---|---|
| X60 | Inten self pois/expos to nonopiod analges antipy & antirheum |
| X600 | Inten self pois/expos to nonopiod analges antipy & antirheum |
| X601 | Inten self pois/expos to nonopiod analges antipy & antirheum |
| X602 | Inten self pois/expos to nonopiod analges antipy & antirheum |
| X603 | Inten self pois/expos to nonopiod analges antipy & antirheum |
| X604 | Inten self pois/expos to nonopiod analges antipy & antirheum |
| X605 | Inten self pois/expos to nonopiod analges antipy & antirheum |
| X606 | Inten self pois/expos to nonopiod analges antipy & antirheum |
| X607 | Inten self pois/expos to nonopiod analges antipy & antirheum |
| X608 | Inten self pois/expos to nonopiod analges antipy & antirheum |
| X609 | Inten self pois/expos to nonopiod analges antipy & antirheum |
| X61 | Int sf pois/expo ant-epi sed-hyp ant park & psy'trop dgs NEC |
| X610 | Int sf pois/expo ant-epi sed-hyp ant park & psy'trop dgs NEC |
| X611 | Int sf pois/expo ant-epi sed-hyp ant park & psy'trop dgs NEC |
| X612 | Int sf pois/expo ant-epi sed-hyp ant park & psy'trop dgs NEC |
| X613 | Int sf pois/expo ant-epi sed-hyp ant park & psy'trop dgs NEC |
| X614 | Int sf pois/expo ant-epi sed-hyp ant park & psy'trop dgs NEC |
| X615 | Int sf pois/expo ant-epi sed-hyp ant park & psy'trop dgs NEC |
| X616 | Int sf pois/expo ant-epi sed-hyp ant park & psy'trop dgs NEC |
| X617 | Int sf pois/expo ant-epi sed-hyp ant park & psy'trop dgs NEC |
| X618 | Int sf pois/expo ant-epi sed-hyp ant park & psy'trop dgs NEC |
| X619 | Int sf pois/expo ant-epi sed-hyp ant park & psy'trop dgs NEC |
| X62 | Inten sf pois/expos narcots & psy'dysleptics [halluc'ns]NEC |
| X620 | Inten sf pois/expos narcots & psy'dysleptics [halluc'ns]NEC |
| X621 | Inten sf pois/expos narcots & psy'dysleptics [halluc'ns]NEC |
| X622 | Inten sf pois/expos narcots & psy'dysleptics [halluc'ns]NEC |
| X623 | Inten sf pois/expos narcots & psy'dysleptics [halluc'ns]NEC |
| X624 | Inten sf pois/expos narcots & psy'dysleptics [halluc'ns]NEC |
| X625 | Inten sf pois/expos narcots & psy'dysleptics [halluc'ns]NEC |
| X626 | Inten sf pois/expos narcots & psy'dysleptics [halluc'ns]NEC |
| X627 | Inten sf pois/expos narcots & psy'dysleptics [halluc'ns]NEC |
| X628 | Inten sf pois/expos narcots & psy'dysleptics [halluc'ns]NEC |
| X629 | Inten sf pois/expos narcots & psy'dysleptics [halluc'ns]NEC |
| X63 | Intent self-poison/expos oth drug act on autonom nervous sys |
| X630 | Intent self-poison/expos oth drug act on autonom nervous sys |
| X631 | Intent self-poison/expos oth drug act on autonom nervous sys |
| X632 | Intent self-poison/expos oth drug act on autonom nervous sys |
| X633 | Intent self-poison/expos oth drug act on autonom nervous sys |
| X634 | Intent self-poison/expos oth drug act on autonom nervous sys |
| X635 | Intent self-poison/expos oth drug act on autonom nervous sys |

| | |
|---|---|
| X636 | Intent self-poison/expos oth drug act on autonom nervous sys |
| X637 | Intent self-poison/expos oth drug act on autonom nervous sys |
| X638 | Intent self-poison/expos oth drug act on autonom nervous sys |
| X639 | Intent self-poison/expos oth drug act on autonom nervous sys |
| X64 | Int self-poison/expos to oth & unsp drugs medics & biol subs |
| X640 | Int self-poison/expos to oth & unsp drugs medics & biol subs |
| X641 | Int self-poison/expos to oth & unsp drugs medics & biol subs |
| X642 | Int self-poison/expos to oth & unsp drugs medics & biol subs |
| X643 | Int self-poison/expos to oth & unsp drugs medics & biol subs |
| X644 | Int self-poison/expos to oth & unsp drugs medics & biol subs |
| X645 | Int self-poison/expos to oth & unsp drugs medics & biol subs |
| X646 | Int self-poison/expos to oth & unsp drugs medics & biol subs |
| X647 | Int self-poison/expos to oth & unsp drugs medics & biol subs |
| X648 | Int self-poison/expos to oth & unsp drugs medics & biol subs |
| X649 | Int self-poison/expos to oth & unsp drugs medics & biol subs |
| X65 | Intentional self-poisoning by and exposure to alcohol |
| X650 | Intentional self-poisoning by and exposure to alcohol |
| X651 | Intentional self-poisoning by and exposure to alcohol |
| X652 | Intentional self-poisoning by and exposure to alcohol |
| X653 | Intentional self-poisoning by and exposure to alcohol |
| X654 | Intentional self-poisoning by and exposure to alcohol |
| X655 | Intentional self-poisoning by and exposure to alcohol |
| X656 | Intentional self-poisoning by and exposure to alcohol |
| X657 | Intentional self-poisoning by and exposure to alcohol |
| X658 | Intentional self-poisoning by and exposure to alcohol |
| X659 | Intentional self-poisoning by and exposure to alcohol |
| X66 | Intent self-poison/expos org solvs+halogen hydrocarbon vapor |
| X660 | Intent self-poison/expos org solvs+halogen hydrocarbon vapor |
| X661 | Intent self-poison/expos org solvs+halogen hydrocarbon vapor |
| X662 | Intent self-poison/expos org solvs+halogen hydrocarbon vapor |
| X663 | Intent self-poison/expos org solvs+halogen hydrocarbon vapor |
| X664 | Intent self-poison/expos org solvs+halogen hydrocarbon vapor |
| X665 | Intent self-poison/expos org solvs+halogen hydrocarbon vapor |
| X666 | Intent self-poison/expos org solvs+halogen hydrocarbon vapor |
| X667 | Intent self-poison/expos org solvs+halogen hydrocarbon vapor |
| X668 | Intent self-poison/expos org solvs+halogen hydrocarbon vapor |
| X669 | Intent self-poison/expos org solvs+halogen hydrocarbon vapor |
| X67 | Intent self-poisoning by and expos to oth gases and vapours |
| X670 | Intent self-poisoning by and expos to oth gases and vapours |
| X671 | Intent self-poisoning by and expos to oth gases and vapours |
| X672 | Intent self-poisoning by and expos to oth gases and vapours |
| X673 | Intent self-poisoning by and expos to oth gases and vapours |
| X674 | Intent self-poisoning by and expos to oth gases and vapours |
| X675 | Intent self-poisoning by and expos to oth gases and vapours |

| | |
|---|---|
| X676 | Intent self-poisoning by and expos to oth gases and vapours |
| X677 | Intent self-poisoning by and expos to oth gases and vapours |
| X678 | Intent self-poisoning by and expos to oth gases and vapours |
| X679 | Intent self-poisoning by and expos to oth gases and vapours |
| X68 | Intentional self-poisoning by and exposure to pesticides |
| X680 | Intentional self-poisoning by and exposure to pesticides |
| X681 | Intentional self-poisoning by and exposure to pesticides |
| X682 | Intentional self-poisoning by and exposure to pesticides |
| X683 | Intentional self-poisoning by and exposure to pesticides |
| X684 | Intentional self-poisoning by and exposure to pesticides |
| X685 | Intentional self-poisoning by and exposure to pesticides |
| X686 | Intentional self-poisoning by and exposure to pesticides |
| X687 | Intentional self-poisoning by and exposure to pesticides |
| X688 | Intentional self-poisoning by and exposure to pesticides |
| X689 | Intentional self-poisoning by and exposure to pesticides |
| X69 | Intent self-poison/expos oth and unspec chems and nox subs |
| X690 | Intent self-poison/expos oth and unspec chems and nox subs |
| X691 | Intent self-poison/expos oth and unspec chems and nox subs |
| X692 | Intent self-poison/expos oth and unspec chems and nox subs |
| X693 | Intent self-poison/expos oth and unspec chems and nox subs |
| X694 | Intent self-poison/expos oth and unspec chems and nox subs |
| X695 | Intent self-poison/expos oth and unspec chems and nox subs |
| X696 | Intent self-poison/expos oth and unspec chems and nox subs |
| X697 | Intent self-poison/expos oth and unspec chems and nox subs |
| X698 | Intent self-poison/expos oth and unspec chems and nox subs |
| X699 | Intent self-poison/expos oth and unspec chems and nox subs |
| X70 | Intent self-harm by hanging strangulation and suffocation |
| X700 | Intent self-harm by hanging strangulation and suffocation |
| X701 | Intent self-harm by hanging strangulation and suffocation |
| X702 | Intent self-harm by hanging strangulation and suffocation |
| X703 | Intent self-harm by hanging strangulation and suffocation |
| X704 | Intent self-harm by hanging strangulation and suffocation |
| X705 | Intent self-harm by hanging strangulation and suffocation |
| X706 | Intent self-harm by hanging strangulation and suffocation |
| X707 | Intent self-harm by hanging strangulation and suffocation |
| X708 | Intent self-harm by hanging strangulation and suffocation |
| X709 | Intent self-harm by hanging strangulation and suffocation |
| X71 | Intentional self-harm by drowning and submersion |
| X710 | Intentional self-harm by drowning and submersion |
| X711 | Intentional self-harm by drowning and submersion |
| X712 | Intentional self-harm by drowning and submersion |
| X713 | Intentional self-harm by drowning and submersion |
| X714 | Intentional self-harm by drowning and submersion |
| X715 | Intentional self-harm by drowning and submersion |

| | |
|---|---|
| X716 | Intentional self-harm by drowning and submersion |
| X717 | Intentional self-harm by drowning and submersion |
| X718 | Intentional self-harm by drowning and submersion |
| X719 | Intentional self-harm by drowning and submersion |
| X72 | Intentional self-harm by handgun discharge |
| X720 | Intentional self-harm by handgun discharge |
| X721 | Intentional self-harm by handgun discharge |
| X722 | Intentional self-harm by handgun discharge |
| X723 | Intentional self-harm by handgun discharge |
| X724 | Intentional self-harm by handgun discharge |
| X725 | Intentional self-harm by handgun discharge |
| X726 | Intentional self-harm by handgun discharge |
| X727 | Intentional self-harm by handgun discharge |
| X728 | Intentional self-harm by handgun discharge |
| X729 | Intentional self-harm by handgun discharge |
| X73 | Intent self-harm by rifle shotgun & larger firearm discharge |
| X730 | Intent self-harm by rifle shotgun & larger firearm discharge |
| X731 | Intent self-harm by rifle shotgun & larger firearm discharge |
| X732 | Intent self-harm by rifle shotgun & larger firearm discharge |
| X733 | Intent self-harm by rifle shotgun & larger firearm discharge |
| X734 | Intent self-harm by rifle shotgun & larger firearm discharge |
| X735 | Intent self-harm by rifle shotgun & larger firearm discharge |
| X736 | Intent self-harm by rifle shotgun & larger firearm discharge |
| X737 | Intent self-harm by rifle shotgun & larger firearm discharge |
| X738 | Intent self-harm by rifle shotgun & larger firearm discharge |
| X739 | Intent self-harm by rifle shotgun & larger firearm discharge |
| X74 | Intent self-harm by other and unspecified firearm discharge |
| X740 | Intent self-harm by other and unspecified firearm discharge |
| X741 | Intent self-harm by other and unspecified firearm discharge |
| X742 | Intent self-harm by other and unspecified firearm discharge |
| X743 | Intent self-harm by other and unspecified firearm discharge |
| X744 | Intent self-harm by other and unspecified firearm discharge |
| X745 | Intent self-harm by other and unspecified firearm discharge |
| X746 | Intent self-harm by other and unspecified firearm discharge |
| X747 | Intent self-harm by other and unspecified firearm discharge |
| X748 | Intent self-harm by other and unspecified firearm discharge |
| X749 | Intent self-harm by other and unspecified firearm discharge |
| X75 | Intentional self-harm by explosive material |
| X750 | Intentional self-harm by explosive material |
| X751 | Intentional self-harm by explosive material |
| X752 | Intentional self-harm by explosive material |
| X753 | Intentional self-harm by explosive material |
| X754 | Intentional self-harm by explosive material |
| X755 | Intentional self-harm by explosive material |

| | |
|---|---|
| X756 | Intentional self-harm by explosive material |
| X757 | Intentional self-harm by explosive material |
| X758 | Intentional self-harm by explosive material |
| X759 | Intentional self-harm by explosive material |
| X76 | Intentional self-harm by smoke fire and flames |
| X760 | Intentional self-harm by smoke fire and flames |
| X761 | Intentional self-harm by smoke fire and flames |
| X762 | Intentional self-harm by smoke fire and flames |
| X763 | Intentional self-harm by smoke fire and flames |
| X764 | Intentional self-harm by smoke fire and flames |
| X765 | Intentional self-harm by smoke fire and flames |
| X766 | Intentional self-harm by smoke fire and flames |
| X767 | Intentional self-harm by smoke fire and flames |
| X768 | Intentional self-harm by smoke fire and flames |
| X769 | Intentional self-harm by smoke fire and flames |
| X77 | Intentional self-harm by steam hot vapours and hot objects |
| X770 | Intentional self-harm by steam hot vapours and hot objects |
| X771 | Intentional self-harm by steam hot vapours and hot objects |
| X772 | Intentional self-harm by steam hot vapours and hot objects |
| X773 | Intentional self-harm by steam hot vapours and hot objects |
| X774 | Intentional self-harm by steam hot vapours and hot objects |
| X775 | Intentional self-harm by steam hot vapours and hot objects |
| X776 | Intentional self-harm by steam hot vapours and hot objects |
| X777 | Intentional self-harm by steam hot vapours and hot objects |
| X778 | Intentional self-harm by steam hot vapours and hot objects |
| X779 | Intentional self-harm by steam hot vapours and hot objects |
| X78 | Intentional self-harm by sharp object |
| X780 | Intentional self-harm by sharp object |
| X781 | Intentional self-harm by sharp object |
| X782 | Intentional self-harm by sharp object |
| X783 | Intentional self-harm by sharp object |
| X784 | Intentional self-harm by sharp object |
| X785 | Intentional self-harm by sharp object |
| X786 | Intentional self-harm by sharp object |
| X787 | Intentional self-harm by sharp object |
| X788 | Intentional self-harm by sharp object |
| X789 | Intentional self-harm by sharp object |
| X79 | Intentional self-harm by blunt object |
| X790 | Intentional self-harm by blunt object |
| X791 | Intentional self-harm by blunt object |
| X792 | Intentional self-harm by blunt object |
| X793 | Intentional self-harm by blunt object |
| X794 | Intentional self-harm by blunt object |
| X795 | Intentional self-harm by blunt object |

| | |
|---|---|
| X796 | Intentional self-harm by blunt object |
| X797 | Intentional self-harm by blunt object |
| X798 | Intentional self-harm by blunt object |
| X799 | Intentional self-harm by blunt object |
| X80 | Intentional self-harm by jumping from a high place |
| X800 | Intentional self-harm by jumping from a high place |
| X801 | Intentional self-harm by jumping from a high place |
| X802 | Intentional self-harm by jumping from a high place |
| X803 | Intentional self-harm by jumping from a high place |
| X804 | Intentional self-harm by jumping from a high place |
| X805 | Intentional self-harm by jumping from a high place |
| X806 | Intentional self-harm by jumping from a high place |
| X807 | Intentional self-harm by jumping from a high place |
| X808 | Intentional self-harm by jumping from a high place |
| X809 | Intentional self-harm by jumping from a high place |
| X81 | Intent self-harm by jumping or lying before moving object |
| X810 | Intent self-harm by jumping or lying before moving object |
| X811 | Intent self-harm by jumping or lying before moving object |
| X812 | Intent self-harm by jumping or lying before moving object |
| X813 | Intent self-harm by jumping or lying before moving object |
| X814 | Intent self-harm by jumping or lying before moving object |
| X815 | Intent self-harm by jumping or lying before moving object |
| X816 | Intent self-harm by jumping or lying before moving object |
| X817 | Intent self-harm by jumping or lying before moving object |
| X818 | Intent self-harm by jumping or lying before moving object |
| X819 | Intent self-harm by jumping or lying before moving object |
| X82 | Intentional self-harm by crashing of motor vehicle |
| X820 | Intentional self-harm by crashing of motor vehicle |
| X821 | Intentional self-harm by crashing of motor vehicle |
| X822 | Intentional self-harm by crashing of motor vehicle |
| X823 | Intentional self-harm by crashing of motor vehicle |
| X824 | Intentional self-harm by crashing of motor vehicle |
| X825 | Intentional self-harm by crashing of motor vehicle |
| X826 | Intentional self-harm by crashing of motor vehicle |
| X827 | Intentional self-harm by crashing of motor vehicle |
| X828 | Intentional self-harm by crashing of motor vehicle |
| X829 | Intentional self-harm by crashing of motor vehicle |
| X83 | Intentional self-harm by other specified means |
| X830 | Intentional self-harm by other specified means |
| X831 | Intentional self-harm by other specified means |
| X832 | Intentional self-harm by other specified means |
| X833 | Intentional self-harm by other specified means |
| X834 | Intentional self-harm by other specified means |
| X835 | Intentional self-harm by other specified means |

| | |
|---|---|
| X836 | Intentional self-harm by other specified means |
| X837 | Intentional self-harm by other specified means |
| X838 | Intentional self-harm by other specified means |
| X839 | Intentional self-harm by other specified means |
| X84 | Intentional self-harm by unspecified means |
| X840 | Intentional self-harm by unspecified means |
| X841 | Intentional self-harm by unspecified means |
| X842 | Intentional self-harm by unspecified means |
| X843 | Intentional self-harm by unspecified means |
| X844 | Intentional self-harm by unspecified means |
| X845 | Intentional self-harm by unspecified means |
| X846 | Intentional self-harm by unspecified means |
| X847 | Intentional self-harm by unspecified means |
| X848 | Intentional self-harm by unspecified means |
| X849 | Intentional self-harm by unspecified means |
| Y10 | Pois/expos nonopiod analges antipy & antirheum undet intent |
| Y100 | Pois/expos nonopiod analges antipy & antirheum undet intent |
| Y101 | Pois/expos nonopiod analges antipy & antirheum undet intent |
| Y102 | Pois/expos nonopiod analges antipy & antirheum undet intent |
| Y103 | Pois/expos nonopiod analges antipy & antirheum undet intent |
| Y104 | Pois/expos nonopiod analges antipy & antirheum undet intent |
| Y105 | Pois/expos nonopiod analges antipy & antirheum undet intent |
| Y106 | Pois/expos nonopiod analges antipy & antirheum undet intent |
| Y107 | Pois/expos nonopiod analges antipy & antirheum undet intent |
| Y108 | Pois/expos nonopiod analges antipy & antirheum undet intent |
| Y109 | Pois/expos nonopiod analges antipy & antirheum undet intent |
| Y11 | Pois/expos ant-epi sed-hyp ant-park/psy'trp dgs undet intent |
| Y110 | Pois/expos ant-epi sed-hyp ant-park/psy'trp dgs undet intent |
| Y111 | Pois/expos ant-epi sed-hyp ant-park/psy'trp dgs undet intent |
| Y112 | Pois/expos ant-epi sed-hyp ant-park/psy'trp dgs undet intent |
| Y113 | Pois/expos ant-epi sed-hyp ant-park/psy'trp dgs undet intent |
| Y114 | Pois/expos ant-epi sed-hyp ant-park/psy'trp dgs undet intent |
| Y115 | Pois/expos ant-epi sed-hyp ant-park/psy'trp dgs undet intent |
| Y116 | Pois/expos ant-epi sed-hyp ant-park/psy'trp dgs undet intent |
| Y117 | Pois/expos ant-epi sed-hyp ant-park/psy'trp dgs undet intent |
| Y118 | Pois/expos ant-epi sed-hyp ant-park/psy'trp dgs undet intent |
| Y119 | Pois/expos ant-epi sed-hyp ant-park/psy'trp dgs undet intent |
| Y12 | Pois/expos narcotics & psy'dys'tics [halluc'ns] undet intent |
| Y120 | Pois/expos narcotics & psy'dys'tics [halluc'ns] undet intent |
| Y121 | Pois/expos narcotics & psy'dys'tics [halluc'ns] undet intent |
| Y122 | Pois/expos narcotics & psy'dys'tics [halluc'ns] undet intent |
| Y123 | Pois/expos narcotics & psy'dys'tics [halluc'ns] undet intent |
| Y124 | Pois/expos narcotics & psy'dys'tics [halluc'ns] undet intent |
| Y125 | Pois/expos narcotics & psy'dys'tics [halluc'ns] undet intent |

| | |
|---|---|
| Y126 | Pois/expos narcotics & psy'dys'tics [halluc'ns] undet intent |
| Y127 | Pois/expos narcotics & psy'dys'tics [halluc'ns] undet intent |
| Y128 | Pois/expos narcotics & psy'dys'tics [halluc'ns] undet intent |
| Y129 | Pois/expos narcotics & psy'dys'tics [halluc'ns] undet intent |
| Y13 | Pois/expos oth drgs acting on autonom nerv syst undet intent |
| Y130 | Pois/expos oth drgs acting on autonom nerv syst undet intent |
| Y131 | Pois/expos oth drgs acting on autonom nerv syst undet intent |
| Y132 | Pois/expos oth drgs acting on autonom nerv syst undet intent |
| Y133 | Pois/expos oth drgs acting on autonom nerv syst undet intent |
| Y134 | Pois/expos oth drgs acting on autonom nerv syst undet intent |
| Y135 | Pois/expos oth drgs acting on autonom nerv syst undet intent |
| Y136 | Pois/expos oth drgs acting on autonom nerv syst undet intent |
| Y137 | Pois/expos oth drgs acting on autonom nerv syst undet intent |
| Y138 | Pois/expos oth drgs acting on autonom nerv syst undet intent |
| Y139 | Pois/expos oth drgs acting on autonom nerv syst undet intent |
| Y14 | Pois/expos oth /unsp drugs medics/biol subs undet intent |
| Y140 | Pois/expos oth /unsp drugs medics/biol subs undet intent |
| Y141 | Pois/expos oth /unsp drugs medics/biol subs undet intent |
| Y142 | Pois/expos oth /unsp drugs medics/biol subs undet intent |
| Y143 | Pois/expos oth /unsp drugs medics/biol subs undet intent |
| Y144 | Pois/expos oth /unsp drugs medics/biol subs undet intent |
| Y145 | Pois/expos oth /unsp drugs medics/biol subs undet intent |
| Y146 | Pois/expos oth /unsp drugs medics/biol subs undet intent |
| Y147 | Pois/expos oth /unsp drugs medics/biol subs undet intent |
| Y148 | Pois/expos oth /unsp drugs medics/biol subs undet intent |
| Y149 | Pois/expos oth /unsp drugs medics/biol subs undet intent |
| Y15 | Poisoning by and exposure to alcohol undetermined intent |
| Y150 | Poisoning by and exposure to alcohol undetermined intent |
| Y151 | Poisoning by and exposure to alcohol undetermined intent |
| Y152 | Poisoning by and exposure to alcohol undetermined intent |
| Y153 | Poisoning by and exposure to alcohol undetermined intent |
| Y154 | Poisoning by and exposure to alcohol undetermined intent |
| Y155 | Poisoning by and exposure to alcohol undetermined intent |
| Y156 | Poisoning by and exposure to alcohol undetermined intent |
| Y157 | Poisoning by and exposure to alcohol undetermined intent |
| Y158 | Poisoning by and exposure to alcohol undetermined intent |
| Y159 | Poisoning by and exposure to alcohol undetermined intent |
| Y16 | Poison/expos org solvs halogen h'carb & vaps undet intent |
| Y160 | Poison/expos org solvs halogen h'carb & vaps undet intent |
| Y161 | Poison/expos org solvs halogen h'carb & vaps undet intent |
| Y162 | Poison/expos org solvs halogen h'carb & vaps undet intent |
| Y163 | Poison/expos org solvs halogen h'carb & vaps undet intent |
| Y164 | Poison/expos org solvs halogen h'carb & vaps undet intent |
| Y165 | Poison/expos org solvs halogen h'carb & vaps undet intent |

| | |
|---|---|
| Y166 | Poison/expos org solvs halogen h'carb & vaps undet intent |
| Y167 | Poison/expos org solvs halogen h'carb & vaps undet intent |
| Y168 | Poison/expos org solvs halogen h'carb & vaps undet intent |
| Y169 | Poison/expos org solvs halogen h'carb & vaps undet intent |
| Y17 | Poison'g/expos to oth gases and vapours undetermined intent |
| Y170 | Poison'g/expos to oth gases and vapours undetermined intent |
| Y171 | Poison'g/expos to oth gases and vapours undetermined intent |
| Y172 | Poison'g/expos to oth gases and vapours undetermined intent |
| Y173 | Poison'g/expos to oth gases and vapours undetermined intent |
| Y174 | Poison'g/expos to oth gases and vapours undetermined intent |
| Y175 | Poison'g/expos to oth gases and vapours undetermined intent |
| Y176 | Poison'g/expos to oth gases and vapours undetermined intent |
| Y177 | Poison'g/expos to oth gases and vapours undetermined intent |
| Y178 | Poison'g/expos to oth gases and vapours undetermined intent |
| Y179 | Poison'g/expos to oth gases and vapours undetermined intent |
| Y18 | Poisoning by and exposure to pesticides undetermined intent |
| Y180 | Poisoning by and exposure to pesticides undetermined intent |
| Y181 | Poisoning by and exposure to pesticides undetermined intent |
| Y182 | Poisoning by and exposure to pesticides undetermined intent |
| Y183 | Poisoning by and exposure to pesticides undetermined intent |
| Y184 | Poisoning by and exposure to pesticides undetermined intent |
| Y185 | Poisoning by and exposure to pesticides undetermined intent |
| Y186 | Poisoning by and exposure to pesticides undetermined intent |
| Y187 | Poisoning by and exposure to pesticides undetermined intent |
| Y188 | Poisoning by and exposure to pesticides undetermined intent |
| Y189 | Poisoning by and exposure to pesticides undetermined intent |
| Y19 | Poison/expos oth/unsp chemicals & noxious subs undet intent |
| Y190 | Poison/expos oth/unsp chemicals & noxious subs undet intent |
| Y191 | Poison/expos oth/unsp chemicals & noxious subs undet intent |
| Y192 | Poison/expos oth/unsp chemicals & noxious subs undet intent |
| Y193 | Poison/expos oth/unsp chemicals & noxious subs undet intent |
| Y194 | Poison/expos oth/unsp chemicals & noxious subs undet intent |
| Y195 | Poison/expos oth/unsp chemicals & noxious subs undet intent |
| Y196 | Poison/expos oth/unsp chemicals & noxious subs undet intent |
| Y197 | Poison/expos oth/unsp chemicals & noxious subs undet intent |
| Y198 | Poison/expos oth/unsp chemicals & noxious subs undet intent |
| Y199 | Poison/expos oth/unsp chemicals & noxious subs undet intent |
| Y20 | Hanging strangulation and suffocation undetermined intent |
| Y200 | Hanging strangulation and suffocation undetermined intent |
| Y201 | Hanging strangulation and suffocation undetermined intent |
| Y202 | Hanging strangulation and suffocation undetermined intent |
| Y203 | Hanging strangulation and suffocation undetermined intent |
| Y204 | Hanging strangulation and suffocation undetermined intent |
| Y205 | Hanging strangulation and suffocation undetermined intent |

| | |
|---|---|
| Y206 | Hanging strangulation and suffocation undetermined intent |
| Y207 | Hanging strangulation and suffocation undetermined intent |
| Y208 | Hanging strangulation and suffocation undetermined intent |
| Y209 | Hanging strangulation and suffocation undetermined intent |
| Y21 | Drowning and submersion undetermined intent |
| Y210 | Drowning and submersion undetermined intent |
| Y211 | Drowning and submersion undetermined intent |
| Y212 | Drowning and submersion undetermined intent |
| Y213 | Drowning and submersion undetermined intent |
| Y214 | Drowning and submersion undetermined intent |
| Y215 | Drowning and submersion undetermined intent |
| Y216 | Drowning and submersion undetermined intent |
| Y217 | Drowning and submersion undetermined intent |
| Y218 | Drowning and submersion undetermined intent |
| Y219 | Drowning and submersion undetermined intent |
| Y22 | Handgun discharge undetermined intent |
| Y220 | Handgun discharge undetermined intent |
| Y221 | Handgun discharge undetermined intent |
| Y222 | Handgun discharge undetermined intent |
| Y223 | Handgun discharge undetermined intent |
| Y224 | Handgun discharge undetermined intent |
| Y225 | Handgun discharge undetermined intent |
| Y226 | Handgun discharge undetermined intent |
| Y227 | Handgun discharge undetermined intent |
| Y228 | Handgun discharge undetermined intent |
| Y229 | Handgun discharge undetermined intent |
| Y23 | Rifle shotgun & larger firearm discharge undetermined intent |
| Y230 | Rifle shotgun & larger firearm discharge undetermined intent |
| Y231 | Rifle shotgun & larger firearm discharge undetermined intent |
| Y232 | Rifle shotgun & larger firearm discharge undetermined intent |
| Y233 | Rifle shotgun & larger firearm discharge undetermined intent |
| Y234 | Rifle shotgun & larger firearm discharge undetermined intent |
| Y235 | Rifle shotgun & larger firearm discharge undetermined intent |
| Y236 | Rifle shotgun & larger firearm discharge undetermined intent |
| Y237 | Rifle shotgun & larger firearm discharge undetermined intent |
| Y238 | Rifle shotgun & larger firearm discharge undetermined intent |
| Y239 | Rifle shotgun & larger firearm discharge undetermined intent |
| Y24 | Other and unspecified firearm discharge undetermined intent |
| Y240 | Other and unspecified firearm discharge undetermined intent |
| Y241 | Other and unspecified firearm discharge undetermined intent |
| Y242 | Other and unspecified firearm discharge undetermined intent |
| Y243 | Other and unspecified firearm discharge undetermined intent |
| Y244 | Other and unspecified firearm discharge undetermined intent |
| Y245 | Other and unspecified firearm discharge undetermined intent |

| | |
|---|---|
| Y246 | Other and unspecified firearm discharge undetermined intent |
| Y247 | Other and unspecified firearm discharge undetermined intent |
| Y248 | Other and unspecified firearm discharge undetermined intent |
| Y249 | Other and unspecified firearm discharge undetermined intent |
| Y25 | Contact with explosive material undetermined intent |
| Y250 | Contact with explosive material undetermined intent |
| Y251 | Contact with explosive material undetermined intent |
| Y252 | Contact with explosive material undetermined intent |
| Y253 | Contact with explosive material undetermined intent |
| Y254 | Contact with explosive material undetermined intent |
| Y255 | Contact with explosive material undetermined intent |
| Y256 | Contact with explosive material undetermined intent |
| Y257 | Contact with explosive material undetermined intent |
| Y258 | Contact with explosive material undetermined intent |
| Y259 | Contact with explosive material undetermined intent |
| Y26 | Exposure to smoke fire and flames undetermined intent |
| Y260 | Exposure to smoke fire and flames undetermined intent |
| Y261 | Exposure to smoke fire and flames undetermined intent |
| Y262 | Exposure to smoke fire and flames undetermined intent |
| Y263 | Exposure to smoke fire and flames undetermined intent |
| Y264 | Exposure to smoke fire and flames undetermined intent |
| Y265 | Exposure to smoke fire and flames undetermined intent |
| Y266 | Exposure to smoke fire and flames undetermined intent |
| Y267 | Exposure to smoke fire and flames undetermined intent |
| Y268 | Exposure to smoke fire and flames undetermined intent |
| Y269 | Exposure to smoke fire and flames undetermined intent |
| Y27 | Contact with steam hot vapours/objects undetermined intent |
| Y270 | Contact with steam hot vapours/objects undetermined intent |
| Y271 | Contact with steam hot vapours/objects undetermined intent |
| Y272 | Contact with steam hot vapours/objects undetermined intent |
| Y273 | Contact with steam hot vapours/objects undetermined intent |
| Y274 | Contact with steam hot vapours/objects undetermined intent |
| Y275 | Contact with steam hot vapours/objects undetermined intent |
| Y276 | Contact with steam hot vapours/objects undetermined intent |
| Y277 | Contact with steam hot vapours/objects undetermined intent |
| Y278 | Contact with steam hot vapours/objects undetermined intent |
| Y279 | Contact with steam hot vapours/objects undetermined intent |
| Y28 | Contact with sharp object undetermined intent |
| Y280 | Contact with sharp object undetermined intent |
| Y281 | Contact with sharp object undetermined intent |
| Y282 | Contact with sharp object undetermined intent |
| Y283 | Contact with sharp object undetermined intent |
| Y284 | Contact with sharp object undetermined intent |
| Y285 | Contact with sharp object undetermined intent |

| | |
|---|---|
| Y286 | Contact with sharp object undetermined intent |
| Y287 | Contact with sharp object undetermined intent |
| Y288 | Contact with sharp object undetermined intent |
| Y289 | Contact with sharp object undetermined intent |
| Y29 | Contact with blunt object undetermined intent |
| Y290 | Contact with blunt object undetermined intent |
| Y291 | Contact with blunt object undetermined intent |
| Y292 | Contact with blunt object undetermined intent |
| Y293 | Contact with blunt object undetermined intent |
| Y294 | Contact with blunt object undetermined intent |
| Y295 | Contact with blunt object undetermined intent |
| Y296 | Contact with blunt object undetermined intent |
| Y297 | Contact with blunt object undetermined intent |
| Y298 | Contact with blunt object undetermined intent |
| Y299 | Contact with blunt object undetermined intent |
| Y30 | Falling jumping/pushed from high place undetermined intent |
| Y300 | Falling jumping/pushed from high place undetermined intent |
| Y301 | Falling jumping/pushed from high place undetermined intent |
| Y302 | Falling jumping/pushed from high place undetermined intent |
| Y303 | Falling jumping/pushed from high place undetermined intent |
| Y304 | Falling jumping/pushed from high place undetermined intent |
| Y305 | Falling jumping/pushed from high place undetermined intent |
| Y306 | Falling jumping/pushed from high place undetermined intent |
| Y307 | Falling jumping/pushed from high place undetermined intent |
| Y308 | Falling jumping/pushed from high place undetermined intent |
| Y309 | Falling jumping/pushed from high place undetermined intent |
| Y31 | Falling lying running before/into moving obj undet intent |
| Y310 | Falling lying running before/into moving obj undet intent |
| Y311 | Falling lying running before/into moving obj undet intent |
| Y312 | Falling lying running before/into moving obj undet intent |
| Y313 | Falling lying running before/into moving obj undet intent |
| Y314 | Falling lying running before/into moving obj undet intent |
| Y315 | Falling lying running before/into moving obj undet intent |
| Y316 | Falling lying running before/into moving obj undet intent |
| Y317 | Falling lying running before/into moving obj undet intent |
| Y318 | Falling lying running before/into moving obj undet intent |
| Y319 | Falling lying running before/into moving obj undet intent |
| Y32 | Crashing of motor vehicle undetermined intent |
| Y320 | Crashing of motor vehicle undetermined intent |
| Y321 | Crashing of motor vehicle undetermined intent |
| Y322 | Crashing of motor vehicle undetermined intent |
| Y323 | Crashing of motor vehicle undetermined intent |
| Y324 | Crashing of motor vehicle undetermined intent |
| Y325 | Crashing of motor vehicle undetermined intent |

| | |
|---|---|
| Y326 | Crashing of motor vehicle undetermined intent |
| Y327 | Crashing of motor vehicle undetermined intent |
| Y328 | Crashing of motor vehicle undetermined intent |
| Y329 | Crashing of motor vehicle undetermined intent |
| Y33 | Other specified events undetermined intent |
| Y330 | Other specified events undetermined intent |
| Y331 | Other specified events undetermined intent |
| Y332 | Other specified events undetermined intent |
| Y333 | Other specified events undetermined intent |
| Y334 | Other specified events undetermined intent |
| Y335 | Other specified events undetermined intent |
| Y336 | Other specified events undetermined intent |
| Y337 | Other specified events undetermined intent |
| Y338 | Other specified events undetermined intent |
| Y339 | Other specified events undetermined intent |
| Y34 | Unspecified event undetermined intent |
| Y340 | Unspecified event undetermined intent |
| Y341 | Unspecified event undetermined intent |
| Y342 | Unspecified event undetermined intent |
| Y343 | Unspecified event undetermined intent |
| Y344 | Unspecified event undetermined intent |
| Y345 | Unspecified event undetermined intent |
| Y346 | Unspecified event undetermined intent |
| Y347 | Unspecified event undetermined intent |
| Y348 | Unspecified event undetermined intent |
| Y349 | Unspecified event undetermined intent |
| Y870 | Sequelae of intentional self-harm |
| Y872 | Sequelae of events of undetermined intent |
| Z915 | Personal history of self-harm |

### Table 4: Self harm related READ codes

| READ codes | Descriptions |
|---|---|
| 14K1. | Intentional overdose of prescription only medication |
| SL... | Poisoning |
| SL90. | Antidepressant poisoning |
| SL900 | Amitriptyline poisoning |
| SL901 | Imipramine poisoning |
| SL902 | Monoamine oxidase inhibitor poisoning |
| SL903 | Trazodone poisoning |
| SL90z | Anti-depressant poisoning NOS |
| TK... | Suicide and selfinflicted injury |
| TK0.. | Suicide + selfinflicted poisoning by solid/liquid substances |
| TK01. | Suicide + selfinflicted poisoning by barbiturates |

| | |
|---|---|
| TK010 | Suicide and self inflicted injury by Amylobarbitone |
| TK011 | Suicide and self inflicted injury by Barbitone |
| TK014 | Suicide and self inflicted injury by Phenobarbitone |
| TK02. | Suicide + selfinflicted poisoning by oth sedatives/hypnotics |
| TK03. | Suicide + selfinflicted poisoning tranquilliser/psychotropic |
| TK04. | Suicide + selfinflicted poisoning by other drugs/medicines |
| TK05. | Suicide + selfinflicted poisoning by drug or medicine NOS |
| TK06. | Suicide + selfinflicted poisoning by agricultural chemical |
| TK07. | Suicide + selfinflicted poisoning by corrosive/caustic subst |
| TK0z. | Suicide + selfinflicted poisoning by solid/liquid subst NOS |
| TK1.. | Suicide + selfinflicted poisoning by gases in domestic use |
| TK10. | Suicide + selfinflicted poisoning by gas via pipeline |
| TK11. | Suicide + selfinflicted poisoning by liquified petrol gas |
| TK1y. | Suicide and selfinflicted poisoning by other utility gas |
| TK1z. | Suicide + selfinflicted poisoning by domestic gases NOS |
| TK2.. | Suicide + selfinflicted poisoning by other gases and vapours |
| TK20. | Suicide + selfinflicted poisoning by motor veh exhaust gas |
| TK21. | Suicide and selfinflicted poisoning by other carbon monoxide |
| TK2z. | Suicide + selfinflicted poisoning by gases and vapours NOS |
| TK3.. | Suicide + selfinflicted injury by hang/strangulate/suffocate |
| TK30. | Suicide and selfinflicted injury by hanging |
| TK31. | Suicide + selfinflicted injury by suffocation by plastic bag |
| TK3y. | Suicide + selfinflicted inj oth mean hang/strangle/suffocate |
| TK3z. | Suicide + selfinflicted inj by hang/strangle/suffocate NOS |
| TK4.. | Suicide and selfinflicted injury by drowning |
| TK5.. | Suicide and selfinflicted injury by firearms and explosives |
| TK51. | Suicide and selfinflicted injury by shotgun |
| TK52. | Suicide and selfinflicted injury by hunting rifle |
| TK53. | Suicide and selfinflicted injury by military firearms |
| TK54. | Suicide and selfinflicted injury by other firearm |
| TK5z. | Suicide and selfinflicted injury by firearms/explosives NOS |
| TK6.. | Suicide and selfinflicted injury by cutting and stabbing |
| TK60. | Suicide and selfinflicted injury by cutting |
| TK601 | Self inflicted lacerations to wrist |
| TK61. | Suicide and selfinflicted injury by stabbing |
| TK6z. | Suicide and selfinflicted injury by cutting and stabbing NOS |
| TK7.. | Suicide and selfinflicted injury by jumping from high place |
| TK70. | Suicide+selfinflicted injury-jump from residential premises |
| TK71. | Suicide+selfinflicted injury-jump from oth manmade structure |
| TK72. | Suicide+selfinflicted injury-jump from natural sites |
| TK7z. | Suicide+selfinflicted injury-jump from high place NOS |
| TKx.. | Suicide and selfinflicted injury by other means |
| TKx0. | Suicide + selfinflicted injury-jump/lie before moving object |
| TKx00 | Suicide + selfinflicted injury-jumping before moving object |

| | |
|---|---|
| TKx1. | Suicide and selfinflicted injury by burns or fire |
| TKx2. | Suicide and selfinflicted injury by scald |
| TKx3. | Suicide and selfinflicted injury by extremes of cold |
| TKx4. | Suicide and selfinflicted injury by electrocution |
| TKx5. | Suicide and selfinflicted injury by crashing motor vehicle |
| TKx6. | Suicide and selfinflicted injury by crashing of aircraft |
| TKx7. | Suicide and selfinflicted injury caustic subst, excl poison |
| TKxy. | Suicide and selfinflicted injury by other specified means |
| TKxz. | Suicide and selfinflicted injury by other means NOS |
| TKz.. | Suicide and selfinflicted injury NOS |
| U2... | [X]Intentional self-harm |
| U20.. | [X]Intentional self poisoning/exposure to noxious substances |
| U200. | [X]Intent self poison/exposure to nonopioid analgesic |
| U2000 | [X]Int self poison/exposure to nonopioid analgesic at home |
| U2001 | [X]Intent self poison nonopioid analgesic at res institut |
| U2002 | [X]Int self poison nonopioid analges school/pub admin area |
| U2003 | [X]Int self poison nonopioid analges in sport/athletic area |
| U2004 | [X]Intent self pois nonopioid analgesic in street/highway |
| U2005 | [X]Intent self pois nonopioid analgesic trade/service area |
| U2006 | [X]Int self pois nonopioid analgesic indust/construct area |
| U2007 | [X]Int self poison/exposure to nonopioid analgesic on farm |
| U200y | [X]Int self poison nonopioid analgesic other spec place |
| U200z | [X]Intent self poison nonopioid analgesic unspecif place |
| U201. | [X]Intent self poison/exposure to antiepileptic |
| U2010 | [X]Int self poison/exposure to antiepileptic at home |
| U2011 | [X]Intent self poison antiepileptic at res institut |
| U2012 | [X]Intent self pois nonopioid analges school/pub admin area |
| U2013 | [X]Int self poison antiepileptic in sport/athletic area |
| U2014 | [X]Intent self pois antiepileptic in street/highway |
| U2015 | [X]Intent self pois antiepileptic trade/service area |
| U2016 | [X]Int self poison antiepileptic indust/construct area |
| U2017 | [X]Int self poison/exposure to antiepileptic on farm |
| U201y | [X]Intent self poison antiepileptic other spec place |
| U201z | [X]Intent self poison antiepileptic unspecif place |
| U202. | [X]Intent self poison/exposure to sedative hypnotic |
| U2020 | [X]Int self poison/exposure to sedative hypnotic at home |
| U2021 | [X]Intent self poison sedative hypnotic at res institut |
| U2022 | [X]Int self poison sedative hypnotic school/pub admin area |
| U2023 | [X]Int self poison sedative hypnotic in sport/athletic area |
| U2024 | [X]Intent self pois sedative hypnotic in street/highway |
| U2025 | [X]Intent self pois sedative hypnotic trade/service area |
| U2026 | [X]Int self pois sedative hypnotic indust/construct area |
| U2027 | [X]Int self poison/exposure to sedative hypnotic on farm |
| U202y | [X]Int self poison sedative hypnotic other spec place |

| | |
|---|---|
| U202z | [X]Intent self poison sedative hypnotic unspecif place |
| U203. | [X]Intent self poison/exposure to antiparkinson drug |
| U2030 | [X]Int self poison/exposure to antiparkinson drug at home |
| U2031 | [X]Intent self poison antiparkinson drug at res institut |
| U2032 | [X]Int self poison antparkinson drug school/pub admin area |
| U2033 | [X]Int self poison antparkinson drug in sport/athletic area |
| U2034 | [X]Intent self pois antiparkinson drug in street/highway |
| U2035 | [X]Intent self pois antiparkinson drug trade/service area |
| U2036 | [X]Int self pois antiparkinson drug indust/construct area |
| U2037 | [X]Int self poison/exposure to antiparkinson drug on farm |
| U203y | [X]Int self poison antiparkinson drug other spec place |
| U203z | [X]Intent self poison antiparkinson drug unspecif place |
| U204. | [X]Intent self poison/exposure to psychotropic drug |
| U2040 | [X]Int self poison/exposure to psychotropic drug at home |
| U2041 | [X]Intent self poison psychotropic drug at res institut |
| U2042 | [X]Int self poison psychotropic drug school/pub admin area |
| U2043 | [X]Int self poison psychotropic drug in sport/athletic area |
| U2044 | [X]Intent self pois psychotropic drug in street/highway |
| U2045 | [X]Intent self pois psychotropic drug trade/service area |
| U2046 | [X]Int self pois psychotropic drug indust/construct area |
| U2047 | [X]Int self poison/exposure to psychotropic drug on farm |
| U204y | [X]Int self poison psychotropic drug other spec place |
| U204z | [X]Intent self poison psychotropic drug unspecif place |
| U205. | [X]Intent self poison/exposure to narcotic drug |
| U2050 | [X]Int self poison/exposure to narcotic drug at home |
| U2051 | [X]Intent self poison narcotic drug at res institut |
| U2052 | [X]Int self poison narcotic drug school/pub admin area |
| U2053 | [X]Int self poison narcotic drug in sport/athletic area |
| U2054 | [X]Intent self pois narcotic drug in street/highway |
| U2055 | [X]Intent self pois narcotic drug trade/service area |
| U2056 | [X]Int self pois narcotic drug indust/construct area |
| U2057 | [X]Int self poison/exposure to narcotic drug on farm |
| U205y | [X]Int self poison narcotic drug other spec place |
| U205z | [X]Intent self poison narcotic drug unspecif place |
| U206. | [X]Intent self poison/exposure to hallucinogen |
| U2060 | [X]Int self poison/exposure to hallucinogen at home |
| U2061 | [X]Intent self poison hallucinogen at res institut |
| U2062 | [X]Int self poison hallucinogenschool/pub admin area |
| U2063 | [X]Int self poison hallucinogenin sport/athletic area |
| U2064 | [X]Intent self pois hallucinogen in street/highway |
| U2065 | [X]Intent self pois hallucinogen trade/service area |
| U2066 | [X]Int self pois hallucinogen indust/construct area |
| U2067 | [X]Int self poison/exposure to hallucinogen on farm |
| U206y | [X]Int self poison hallucinogen other spec place |

| | |
|---|---|
| U206z | [X]Intent self poison hallucinogen unspecif place |
| U207. | [X]Intent self poison/exposure to oth autonomic drug |
| U2070 | [X]Int self poison/exposure to oth autonomic drug at home |
| U2071 | [X]Intent self poison oth autonomic drug at res institut |
| U2072 | [X]Int self poison oth autonom drug school/pub admin area |
| U2073 | [X]Int self poison oth autonom drug in sport/athletic area |
| U2074 | [X]Intent self pois oth autonomic drug in street/highway |
| U2075 | [X]Intent self pois oth autonomic drug trade/service area |
| U2076 | [X]Int self pois oth autonomic drug indust/construct area |
| U2077 | [X]Int self poison/exposure to oth autonomic drug on farm |
| U207y | [X]Int self poison oth autonomic drug other spec place |
| U207z | [X]Intent self poison oth autonomic drug unspecif place |
| U208. | [X]Int self poison/exposure to other/unspec drug/medicament |
| U2080 | [X]Int self poison/exposure to oth/unsp drug/medicam home |
| U2081 | [X]Intent self poison oth/unsp drug/medicam res institut |
| U2082 | [X]Int self poison oth/uns drug/med school/pub admin area |
| U2083 | [X]Int self poison oth/uns drug/med in sport/athletic area |
| U2084 | [X]Intent self pois oth/unsp drug/medic in street/highway |
| U2085 | [X]Intent self pois oth/unsp drug/medic trade/service area |
| U2086 | [X]Int self pois oth/unsp drug/medic indust/construct area |
| U2087 | [X]Int self poison/exposure to oth/unsp drug/medic on farm |
| U208y | [X]Int self poison oth/unsp drug/medic other spec place |
| U208z | [X]Intent self poison oth/unsp drug/medic unspecif place |
| U20A. | [X]Intentional self poison organ solvent,halogen hydrocarb |
| U20A0 | [X]Intent self pois organ solvent,halogen hydrocarb, home |
| U20A1 | [X]Int self poison org solvent,halogen hydrocarb,res instit |
| U20A2 | [X]Int self poison org solvent,halogen hydrocarb, school |
| U20A3 | [X]Int self poison org solvent,halogen hydrocarb,sport area |
| U20A4 | [X]Int self poison org solvent,halogen hydrocarb,in highway |
| U20A5 | [X]Int self poison org solvent,halogen hydrocarb,trade area |
| U20A6 | [X]Int self pois org solvent,halogen hydrocarb,indust area |
| U20A7 | [X]Int self poison org solvent,halogen hydrocarb,on farm |
| U20Ay | [X]Int self pois org solv,halogen hydrocarb,oth spec place |
| U20Az | [X]Int self pois org solv,halogen hydrocarb, unspec place |
| U20B. | [X]Intent self poison/exposure to other gas/vapour |
| U20B0 | [X]Int self poison/exposure to other gas/vapour at home |
| U20B1 | [X]Intent self poison other gas/vapour at res institut |
| U20B2 | [X]Int self poison other gas/vapour school/pub admin area |
| U20B3 | [X]Int self poison other gas/vapour in sport/athletic area |
| U20B4 | [X]Intent self pois other gas/vapour in street/highway |
| U20B5 | [X]Intent self pois other gas/vapour trade/service area |
| U20B6 | [X]Int self pois other gas/vapour indust/construct area |
| U20B7 | [X]Int self poison/exposure to other gas/vapour on farm |
| U20By | [X]Int self poison other gas/vapour other spec place |

| | |
|---|---|
| U20Bz | [X]Intent self poison other gas/vapour unspecif place |
| U20C. | [X]Intent self poison/exposure to pesticide |
| U20C0 | [X]Int self poison/exposure to pesticide at home |
| U20C1 | [X]Intent self poison pesticide at res institut |
| U20C2 | [X]Int self poison pesticide school/pub admin area |
| U20C3 | [X]Int self poison pesticide in sport/athletic area |
| U20C4 | [X]Intent self pois pesticide in street/highway |
| U20C5 | [X]Intent self pois pesticide trade/service area |
| U20C6 | [X]Int self pois pesticide indust/construct area |
| U20C7 | [X]Int self poison/exposure to pesticide on farm |
| U20Cy | [X]Int self poison pesticide other spec place |
| U20Cz | [X]Intent self poison pesticide unspecif place |
| U20y. | [X]Intent self poison/exposure to unspecif chemical |
| U20y0 | [X]Int self poison/exposure to unspecif chemical at home |
| U20y1 | [X]Intent self poison unspecif chemical at res institut |
| U20y2 | [X]Int self poison unspecif chemical school/pub admin area |
| U20y3 | [X]Int self poison unspecif chemical in sport/athletic area |
| U20y4 | [X]Intent self pois unspecif chemical in street/highway |
| U20y5 | [X]Intent self pois unspecif chemical trade/service area |
| U20y6 | [X]Int self pois unspecif chemical indust/construct area |
| U20y7 | [X]Int self poison/exposure to unspecif chemical on farm |
| U20yy | [X]Int self poison unspecif chemical other spec place |
| U20yz | [X]Intent self poison unspecif chemical unspecif place |
| U21.. | [X]Intent self harm by hanging strangulation / suffocation |
| U210. | [X]Intent self harm by hanging strangulat/suffocat occ home |
| U211. | [X]Intent self harm by hangng strangult/suffoct resid instit |
| U212. | [X]Inten slf harm hang strang/suffc sch oth ins/pub adm area |
| U213. | [X]Intent self harm by hang strangl/suffc sport/athlet area |
| U214. | [X]Intent self harm by hangng strangult/suffoct street/h'way |
| U215. | [X]Intent self harm by hang strangl/suffc trade/service area |
| U216. | [X]Intent self harm by hang strangl/suffc indust/constr area |
| U217. | [X]Intent self harm by hanging strangulat/suffocat occ farm |
| U21y. | [X]Intent self harm by hangng strangul/suffoct oth spec plce |
| U21z. | [X]Intent self harm by hangng strangul/suffoct unspecif plce |
| U22.. | [X]Intentional self harm by drowning and submersion |
| U220. | [X]Intent self harm by drowning/submersion occurrn at home |
| U221. | [X]Intent self harm by drowning/submersn occ resid instit'n |
| U222. | [X]Intent self harm drown/submers occ sch/ins/pub adm area |
| U223. | [X]Intent self harm by drown/submersn occ sport/athlet area |
| U224. | [X]Intent self harm by drowning/submersn occ street/highway |
| U225. | [X]Intent self harm by drown/submersn occ trade/servce area |
| U226. | [X]Intent self harm by drown/submers occ indust/constr area |
| U227. | [X]Intent self harm by drowning/submersion occurrn on farm |
| U22y. | [X]Intent self harm by drown/submersn occ oth specif place |

| | |
|---|---|
| U22z. | [X]Intent self harm by drown/submersn occ unspecified place |
| U23.. | [X]Intentional self harm by handgun discharge |
| U230. | [X]Intention self harm by handgun discharge occurrn at home |
| U231. | [X]Intent self harm by handgun disch occ in resid instit'n |
| U232. | [X]Intent self harm h'gun disch occ sch oth ins/pub adm area |
| U233. | [X]Intent self harm by handgun disch occ sport/athlet area |
| U234. | [X]Intent self harm by handgun disch occ on street/highway |
| U235. | [X]Intent self harm by handgun disch occ trade/service area |
| U236. | [X]Intent self harm by handgun disch occ indust/constr area |
| U237. | [X]Intention self harm by handgun discharge occurrn on farm |
| U23y. | [X]Intent self harm by handgun disch occ at oth specif plce |
| U23z. | [X]Intent self harm by handgun disch occ at unspecif place |
| U24.. | [X]Intent self harm by rifle shotgun/larger firearm disch |
| U240. | [X]Intent self harm rifle sh'gun/largr firarm disch occ home |
| U241. | [X]Int self harm rifl s'gun/lrg frarm disch occ resid instit |
| U242. | [X]Int slf hrm rifl s'gun/lrg frarm dis sch/ins/pub adm area |
| U243. | [X]Int self harm rifl s'gun/lrg frarm disch sprt/athlet area |
| U244. | [X]Int self harm rifl s'gun/lrg frarm disch occ street/h'way |
| U245. | [X]Int self harm rifl s'gun/lrg frarm disch trad/servce area |
| U246. | [X]Int slf hrm rifl s'gun/lrg frarm disch indust/constr area |
| U247. | [X]Intent self harm rifle sh'gun/largr firarm disch occ farm |
| U24y. | [X]Int self harm rifl s'gun/lrg frarm disch oth specif place |
| U24z. | [X]Int self harm rifl s'gun/lrg frarm disch occ unspec place |
| U25.. | [X]Intent self harm by other/unspecified firearm discharge |
| U250. | [X]Intent self harm oth/unspecif firearm disch occ at home |
| U251. | [X]Intent self harm oth/unsp firearm disch occ resid instit |
| U252. | [X]Inten self harm oth/uns firarm disch sch/ins/pub adm area |
| U253. | [X]Inten self harm oth/uns firearm disch occ sprt/athl area |
| U254. | [X]Intent self harm oth/unsp firearm disch occ street/h'way |
| U255. | [X]Intent self harm oth/uns firearm disch trade/servce area |
| U256. | [X]Inten self harm oth/uns firearm disch indust/constr area |
| U257. | [X]Intent self harm oth/unspecif firearm disch occ on farm |
| U25y. | [X]Intent self harm oth/unsp firearm disch oth specif place |
| U25z. | [X]Intent self harm oth/unsp firearm disch occ unspecif plce |
| U26.. | [X]Intentional self harm by explosive material |
| U260. | [X]Intention self harm by explosive material occurrn home |
| U261. | [X]Intention self harm by explosiv materl occ resid instit |
| U262. | [X]Intent self harm by explosiv materl sch/ins/pub adm area |
| U263. | [X]Intent self harm by explosv materl occ sport/athlet area |
| U264. | [X]Intention self harm by explosiv materl occ street/highway |
| U265. | [X]Intent self harm by explosv materl occ trade/servce area |
| U266. | [X]Intent self harm by explosv materl occ indust/constr area |
| U267. | [X]Intention self harm by explosive material occurrn farm |
| U26y. | [X]Intent self harm by explosiv materl occ oth specif place |

| | |
|---|---|
| U26z. | [X]Intent self harm by explosiv materl occ unspecif place |
| U27.. | [X]Intentional self harm by smoke, fire and flames |
| U270. | [X]Intention self harm by smoke fire/flames occurrn at home |
| U271. | [X]Intent self harm by smoke fire/flame occ resid instit'n |
| U272. | [X]Intent self harm by smoke fire/flame sch/ins/pub adm area |
| U273. | [X]Intent self harm by smok fire/flam occ sport/athlet area |
| U274. | [X]Intent self harm by smoke fire/flame occ street/highway |
| U275. | [X]Intent self harm by smok fire/flam occ trade/servce area |
| U276. | [X]Intent self harm by smok fire/flam occ indust/constr area |
| U277. | [X]Intention self harm by smoke fire/flames occurrn on farm |
| U27y. | [X]Intent self harm by smoke fire/flame occ oth specif plce |
| U27z. | [X]Intent self harm by smoke fire/flames occ unspecif place |
| U28.. | [X]Intentional self harm by steam hot vapours / hot objects |
| U280. | [X]Intent self harm by steam hot vapour/hot obj occ at home |
| U281. | [X]Intent self harm by steam hot vapour/obj occ resid instit |
| U282. | [X]Int self harm by steam hot vapor/obj sch/ins/pub adm area |
| U283. | [X]Int self harm by steam hot vapour/obj occ sport/athl area |
| U284. | [X]Intent self harm by steam hot vapour/obj occ street/h'way |
| U285. | [X]Int self harm by steam hot vapour/obj trade/service area |
| U286. | [X]Int self harm by steam hot vapour/obj indust/constr area |
| U287. | [X]Intent self harm by steam hot vapour/hot obj occ on farm |
| U28y. | [X]Intent self harm by steam hot vapour/obj oth specif place |
| U28z. | [X]Intent self harm by steam hot vapour/obj occ unspec place |
| U29.. | [X]Intentional self harm by sharp object |
| U290. | [X]Intentional self harm by sharp object occurrence at home |
| U291. | [X]Intent self harm by sharp object occ resident instit'n |
| U292. | [X]Intent self harm sharp obj occ sch oth ins/pub adm area |
| U293. | [X]Intent self harm by sharp object occ sports/athlet area |
| U294. | [X]Intention self harm by sharp object occ street/highway |
| U295. | [X]Intent self harm by sharp object occ trade/service area |
| U296. | [X]Intent self harm by sharp object occ indust/constr area |
| U297. | [X]Intentional self harm by sharp object occurrence on farm |
| U29y. | [X]Intention self harm by sharp object occ oth specif place |
| U29z. | [X]Intentional self harm by sharp object occ unspecif place |
| U2A.. | [X]Intentional self harm by blunt object |
| U2A0. | [X]Intentional self harm by blunt object occurrence at home |
| U2A1. | [X]Intent self harm by blunt object occ resident instit'n |
| U2A2. | [X]Intent self harm blunt obj occ sch oth ins/pub adm area |
| U2A3. | [X]Intent self harm by blunt object occ sports/athlet area |
| U2A4. | [X]Intention self harm by blunt object occ street/highway |
| U2A5. | [X]Intent self harm by blunt object occ trade/service area |
| U2A6. | [X]Intent self harm by blunt object occ indust/constr area |
| U2A7. | [X]Intentional self harm by blunt object occurrence on farm |
| U2Ay. | [X]Intention self harm by blunt object occ oth specif place |

| | |
|---|---|
| U2Az. | [X]Intentional self harm by blunt object occ unspecif place |
| U2B.. | [X]Intentional self harm by jumping from a high place |
| U2B0. | [X]Intent self harm by jumping from high place occ at home |
| U2B1. | [X]Intent self harm by jump from high place occ resid instit |
| U2B2. | [X]Int self harm jump fr high place sch oth ins/pub adm area |
| U2B3. | [X]Intent self harm by jump from high place sport/athl area |
| U2B4. | [X]Intent self harm by jump from high place occ street/h'way |
| U2B5. | [X]Intent self harm by jump from high place trad/servce area |
| U2B6. | [X]Int self harm by jump from high place indust/constr area |
| U2B7. | [X]Intent self harm by jumping from high place occ on farm |
| U2By. | [X]Int self harm by jump from high place occ oth specif plce |
| U2Bz. | [X]Int self harm by jump from high place occ unspecif place |
| U2C.. | [X]Intent self harm by jumping / lying before moving object |
| U2C0. | [X]Intent self harm by jump/lying befor moving obj occ home |
| U2C1. | [X]Int self harm jump/lying befr mov obje occ resid instit'n |
| U2C2. | [X]Int self harm jump/lying bef mov obj sch/ins/pub adm area |
| U2C3. | [X]Int self harm jump/lying bef mov obj occ sprt/athlet area |
| U2C4. | [X]Int self harm jump/lying befr mov obje occ street/highway |
| U2C5. | [X]Int self harm jump/lying bef mov obj occ trad/servce area |
| U2C6. | [X]Int self harm jump/lying befr mov obj indust/constr area |
| U2C7. | [X]Intent self harm by jump/lying befor moving obj occ farm |
| U2Cy. | [X]Int self harm jump/lying bef mov obje occ oth specif plce |
| U2Cz. | [X]Int self harm jump/lying bef mov obje occ unspecif place |
| U2D.. | [X]Intentional self harm by crashing of motor vehicle |
| U2D0. | [X]Intent self harm by crash of motor vehicl occurrn at home |
| U2D1. | [X]Intent self harm by crash motor vehicl occ resid instit'n |
| U2D2. | [X]Int self harm crash motor vehicl occ sch/ins/pub adm area |
| U2D3. | [X]Intent self harm by crash motor vehicl occ sprt/athl area |
| U2D4. | [X]Intent self harm by crash motor vehicl occ street/highway |
| U2D5. | [X]Intent self harm crash motor vehicl occ trade/servce area |
| U2D6. | [X]Intent self harm crash motor vehic occ indust/constr area |
| U2D7. | [X]Intent self harm by crash of motor vehicl occurrn on farm |
| U2Dy. | [X]Intent self harm by crash motor vehic occ oth specif plce |
| U2Dz. | [X]Intent self harm by crash motor vehic occ unspecif place |
| U2E.. | [X]Self mutilation |
| U2y.. | [X]Intentional self harm by other specified means |
| U2y0. | [X]Intentionl self harm by oth specif means occurrn at home |
| U2y1. | [X]Intent self harm by oth specif means occ resid instit'n |
| U2y2. | [X]Intent self harm oth specif mean occ sch/ins/pub adm area |
| U2y3. | [X]Intent self harm by oth specif means occ sport/athl area |
| U2y4. | [X]Intent self harm by oth specif means occ street/highway |
| U2y5. | [X]Intent self harm by oth specif means occ trad/servce area |
| U2y6. | [X]Intent self harm oth specif means occ indust/constr area |
| U2y7. | [X]Intentionl self harm by oth specif means occurrn on farm |

| | |
|---|---|
| U2yy. | [X]Intent self harm oth specif means occ oth specif place |
| U2yz. | [X]Intent self harm by oth specif means occ unspecif place |
| U2z.. | [X]Intentional self harm by unspecified means |
| U2z0. | [X]Intentional self harm by unspecif means occurrn at home |
| U2z1. | [X]Intent self harm by unspecif means occurrn resid instit'n |
| U2z2. | [X]Intent self harm by unspec mean occ sch/ins/pub adm area |
| U2z3. | [X]Intent self harm unspecif means occurrn sport/athlet area |
| U2z4. | [X]Intent self harm by unspecif means occurrn street/highway |
| U2z5. | [X]Intent self harm unspecif means occurrn trade/servce area |
| U2z6. | [X]Intent self harm unspecif mean occurrn indust/constr area |
| U2z7. | [X]Intentional self harm by unspecif means occurrn on farm |
| U2zy. | [X]Intent self harm by unspecif means occ oth specif place |
| U2zz. | [X]Intent self harm by unspecif means occ at unspecif place |
| U41.. | [X]Hanging strangulation + suffocation undetermined intent |
| U44.. | [X]Rifle shotgun+larger firearm discharge undetermin intent |
| U45.. | [X]Other+unspecified firearm discharge undetermined intent |
| U4B.. | [X]Falling jumping/pushed from high place undeterm intent |
| U4Bz. | [X]Fall jump/push frm high plce undt intnt occ unspecif plce |
| U72.. | [X]Sequel intentn self-harm assault+event of undeterm intent |
| U720. | [X]Sequelae of intentional self-harm |
| ZRLfC12 | Health of the Nation Outcome Scales item 2– nonaccidental self-injury |
| ZX... | Self-harm |
| ZX1.. | Self-injurious behaviour |
| ZX11. | Biting self |
| ZX12. | Burning self |
| ZX13. | Cutting self |
| ZX15. | Drowning self |
| ZX18. | Hanging self |
| ZX19. | Hitting self |
| ZX191 | Punching self |
| ZX192 | Slapping self |
| ZX1B. | Jumping from height |
| ZX1B1 | Jumping from building |
| ZX1B2 | Jumping from bridge |
| ZX1B3 | Jumping from cliff |
| ZX1C. | Nipping self |
| ZX1E. | Pinching self |
| ZX1G. | Scratches self |
| ZX1H. | Self-asphyxiation |
| ZX1H1 | Self-strangulation |
| ZX1H2 | Self-suffocation |
| ZX1I. | Self-scalding |
| ZX1J. | Self-electrocution |
| ZX1K. | Self-incineration |

| | |
|---|---|
| ZX1L. | Self-mutilation |
| ZX1L1 | Self-mutilation of hands |
| ZX1L2 | Self-mutilation of genitalia |
| ZX1L3 | Self-mutilation of penis |
| ZX1L6 | Self-mutilation of ears |
| ZX1LD | [X]Self mutilation |
| ZX1M. | Shooting self |
| ZX1N. | Stabbing self |
| ZX1Q. | Throwing self in front of train |
| ZX1R. | Throwing self in front of vehicle |
| ZX1S. | Throwing self onto floor |

### Table 5: Depression related ICD10 codes

| **ICD10 codes** | **Descriptions** |
|---|---|
| F251 | Schizoaffective disorder depressive type |
| F32 | Depressive episode |
| F320 | Mild depressive episode |
| F321 | Moderate depressive episode |
| F322 | Severe depressive episode without psychotic symptoms |
| F323 | Severe depressive episode with psychotic symptoms |
| F328 | Other depressive episodes |
| F329 | Depressive episode, unspecified |
| F33 | Recurrent depressive disorder |
| F330 | Recurrent depressive disorder, current episode mild |
| F331 | Recurrent depressive disorder, current episode moderate |
| F332 | Recurrent depressive disorder, current episode severe without psychotic symptoms |
| F333 | Recurrent depressive disorder, current episode severe with psychotic symptoms |
| F334 | Recurrent depressive disorder, currently in remission |
| F338 | Other recurrent depressive disorders |
| F339 | Recurrent depressive disorder, unspecified |
| F341 | Dysthymia |

### Table 6: Depression related READ codes

| RAED codes | Descriptions |
|---|---|
| 1B17. | Depressed |
| 1B1U. | Symptoms of depression |
| 1BQ.. | Loss of capacity for enjoyment |
| 1BT.. | Depressed mood |
| 1BU.. | Loss of hope for the future |
| 2257 | O/E - depressed |

| | |
|---|---|
| E0013 | Presenile dementia with depression |
| E0021 | Senile dementia with depression |
| E112. | Single major depressive episode |
| E1120 | Single major depressive episode, unspecified |
| E1121 | Single major depressive episode, mild |
| E1122 | Single major depressive episode, moderate |
| E1123 | Single major depressive episode, severe, without psychosis |
| E1124 | Single major depressive episode, severe, with psychosis |
| E1125 | Single major depressive episode, partial or unspec remission |
| E1126 | Single major depressive episode, in full remission |
| E112z | Single major depressive episode NOS |
| E113. | Recurrent major depressive episode |
| E1130 | Recurrent major depressive episodes, unspecified |
| E1131 | Recurrent major depressive episodes, mild |
| E1132 | Recurrent major depressive episodes, moderate |
| E1133 | Recurrent major depressive episodes, severe, no psychosis |
| E1134 | Recurrent major depressive episodes, severe, with psychosis |
| E1135 | Recurrent major depressive episodes,partial/unspec remission |
| E1136 | Recurrent major depressive episodes, in full remission |
| E1137 | Recurrent depression |
| E113z | Recurrent major depressive episode NOS |
| E118. | Seasonal affective disorder |
| E11y2 | Atypical depressive disorder |
| E11z2 | Masked depression |
| E130. | Reactive depressive psychosis |
| E135. | Agitated depression |
| E2003 | Anxiety with depression |
| E204. | Neurotic depression reactive type |
| E291. | Prolonged depressive reaction |
| E2B.. | Depressive disorder NEC |
| E2B0. | Postviral depression |
| E2B1. | Chronic depression |
| Eu204 | [X]Post-schizophrenic depression |
| Eu251 | [X]Schizoaffective disorder, depressive type |
| Eu32. | [X]Depressive episode |
| Eu320 | [X]Mild depressive episode |
| Eu321 | [X]Moderate depressive episode |
| Eu322 | [X]Severe depressive episode without psychotic symptoms |
| Eu323 | [X]Severe depressive episode with psychotic symptoms |
| Eu324 | [X]Mild depression |
| Eu325 | [X]Major depression, mild |
| Eu326 | [X]Major depression, moderately severe |
| Eu327 | [X]Major depression, severe without psychotic symptoms |
| Eu328 | [X]Major depression, severe with psychotic symptoms |

| | |
|---|---|
| Eu32y | [X]Other depressive episodes |
| Eu32z | [X]Depressive episode, unspecified |
| Eu33. | [X]Recurrent depressive disorder |
| Eu330 | [X]Recurrent depressive disorder, current episode mild |
| Eu331 | [X]Recurrent depressive disorder, current episode moderate |
| Eu332 | [X]Recurr depress disorder cur epi severe without psyc sympt |
| Eu333 | [X]Recurrent depress disorder cur epi severe with psyc symp |
| Eu334 | [X]Recurrent depressive disorder, currently in remission |
| Eu33y | [X]Other recurrent depressive disorders |
| Eu33z | [X]Recurrent depressive disorder, unspecified |
| Eu341 | [X]Dysthymia |
| Eu412 | [X]Mixed anxiety and depressive disorder |

#### Table 7: Depression - medication READ codes

| RAED codes | Descriptions |
|---|---|
| d11.. | CHLORAL HYDRATE |
| d12.. | CLOMETHIAZOLE EDISYLATE [HYPNOTIC] |
| d13.. | *DICHLORALPHENAZONE |
| d14.. | *FLUNITRAZEPAM |
| d15.. | FLURAZEPAM |
| d16.. | LOPRAZOLAM |
| d17.. | LORMETAZEPAM |
| d18.. | NITRAZEPAM |
| d1a.. | TEMAZEPAM [HYPNOTIC] |
| d1b.. | *TRIAZOLAM |
| d1c.. | TRICLOFOS SODIUM |
| d1d.. | ZOPICLONE |
| d1f.. | ZOLPIDEM |
| d1g.. | ZALEPLON |
| d1h.. | MELATONIN |
| d21.. | DIAZEPAM [ANXIOLYTIC] |
| d22.. | ALPRAZOLAM |
| d23.. | BROMAZEPAM |
| d24.. | CHLORDIAZEPOXIDE |
| d25.. | CHLORMEZANONE |
| d26.. | CLOBAZAM |
| d27.. | CLORAZEPATE DIPOTASSIUM |
| d28.. | HYDROXYZINE HCL [ANXIOLYTIC] |
| d29.. | *KETAZOLAM |
| d2a.. | LORAZEPAM [ANXIOLYTIC] |
| d2b.. | *MEDAZEPAM |
| d2c.. | MEPROBAMATE |
| d2d.. | OXAZEPAM |

| | |
|---|---|
| d2e.. | *PRAZEPAM |
| d2f.. | BUSPIRONE HYDROCHLORIDE |
| d2g.. | FLUMAZENIL |
| d71.. | AMITRIPTYLINE HYDROCHLORIDE [ANTIDEPRESSANT] |
| d72.. | *BUTRIPTYLINE |
| d73.. | CLOMIPRAMINE HYDROCHLORIDE |
| d74.. | DESIPRAMINE HYDROCHLORIDE |
| d75.. | DOSULEPIN HYDROCHLORIDE |
| d76.. | DOXEPIN |
| d77.. | IMIPRAMINE HYDROCHLORIDE [ANTIDEPRESSANT] |
| d78.. | IPRINDOLE |
| d79.. | LOFEPRAMINE |
| d7a.. | MAPROTILINE HYDROCHLORIDE |
| d7b.. | MIANSERIN HYDROCHLORIDE |
| d7c.. | NORTRIPTYLINE |
| d7d.. | PROTRIPTYLINE HYDROCHLORIDE |
| d7e.. | TRAZODONE HYDROCHLORIDE |
| d7f.. | TRIMIPRAMINE |
| d7g.. | VILOXAZINE HYDROCHLORIDE |
| d7h.. | AMOXAPINE |
| d81.. | PHENELZINE |
| d83.. | ISOCARBOXAZID |
| d84.. | TRANYLCYPROMINE |
| d85.. | MOCLOBEMIDE |
| d91.. | COMPOUND ANTIDEPRESSANTS A-Z |
| da1.. | FLUPENTIXOL [ANTIDEPRESSANT] |
| da2.. | TRYPTOPHAN |
| da3.. | FLUVOXAMINE MALEATE |
| da4.. | FLUOXETINE HYDROCHLORIDE |
| da5.. | SERTRALINE HYDROCHLORIDE |
| da6.. | PAROXETINE HYDROCHLORIDE |
| da7.. | VENLAFAXINE |
| da9.. | CITALOPRAM |
| daA.. | REBOXETINE |
| daB.. | MIRTAZAPINE |
| daC.. | ESCITALOPRAM |
| daD.. | AGOMELATINE |
| gde.. | DULOXETINE |

#### Table 8: Serious Mental Illness related ICD10 codes

| ICD10 codes | Descriptions |
|---|---|
| F200 | Paranoid schizophrenia |
| F201 | Hebephrenic schizophrenia |

| | |
|---|---|
| F202 | Catatonic schizophrenia |
| F203 | Undifferentiated schizophrenia |
| F204 | Post-schizophrenic depression |
| F205 | Residual schizophrenia |
| F206 | Simple schizophrenia |
| F208 | Other schizophrenia |
| F209 | Schizophreniaunspecified |
| F21X | Schizotypal disorder |
| F220 | Delusional disorder |
| F228 | Other persistent delusional disorders |
| F229 | Persistent delusional disorderunspecified |
| F230 | Acute polymorphic psychot disord without symp of schizoph'a |
| F231 | Acute polymorphic psychot disord with symp of schizophrenia |
| F232 | Acute schizophrenia-like psychotic disorder |
| F233 | Other acute predominantly delusional psychotic disorders |
| F238 | Other acute and transient psychotic disorders |
| F239 | Acute and transient psychotic disorderunspecified |
| F24X | Induced delusional disorder |
| F250 | Schizoaffective disordermanic type |
| F251 | Schizoaffective disorderdepressive type |
| F252 | Schizoaffective disordermixed type |
| F258 | Other schizoaffective disorders |
| F259 | Schizoaffective disorderunspecified |
| F28X | Other nonorganic psychotic disorders |
| F29X | Unspecified nonorganic psychosis |
| F300 | Hypomania |
| F301 | Mania without psychotic symptoms |
| F302 | Mania with psychotic symptoms |
| F308 | Other manic episodes |
| F309 | Manic episodeunspecified |
| F310 | Bipolar affective disordercurrent episode hypomanic |
| F311 | Bipolar affect disord cur epi manic without psychotic symp |
| F312 | Bipolar affect disorder cur epi manic with psychotic symp |
| F313 | Bipolar affect disorder cur epi mild or moderate depression |
| F314 | Bipolar affect disord cur epi sev depres without psyc symp |
| F315 | Bipolar affect disord cur epi severe depres with psyc symp |
| F316 | Bipolar affective disordercurrent episode mixed |
| F317 | Bipolar affective disordercurrently in remission |
| F318 | Other bipolar affective disorders |
| F319 | Bipolar affective disorderunspecified |
| F323 | Severe depressive episode with psychotic symptoms |
| F333 | Recurrent depress disorder cur epi severe with psyc symp |
| F39X | Unspecified mood [affective] disorder |

#### Table 9: Serious mental Illness related READ codes

| RAED codes | Descriptions |
|---|---|
| E10.. | Schizophrenic disorders |
| E100. | Simple schizophrenia |
| E1000 | Unspecified schizophrenia |
| E1001 | Subchronic schizophrenia |
| E1002 | Chronic schizophrenic |
| E1003 | Acute exacerbation of subchronic schizophrenia |
| E1004 | Acute exacerbation of chronic schizophrenia |
| E1005 | Schizophrenia in remission |
| E100z | Simple schizophrenia NOS |
| E101. | Hebephrenic schizophrenia |
| E1010 | Unspecified hebephrenic schizophrenia |
| E1011 | Subchronic hebephrenic schizophrenia |
| E1012 | Chronic hebephrenic schizophrenia |
| E1013 | Acute exacerbation of subchronic hebephrenic schizophrenia |
| E1014 | Acute exacerbation of chronic hebephrenic schizophrenia |
| E1015 | Hebephrenic schizophrenia in remission |
| E101z | Hebephrenic schizophrenia NOS |
| E102. | Catatonic schizophrenia |
| E1020 | Unspecified catatonic schizophrenia |
| E1021 | Subchronic catatonic schizophrenia |
| E1022 | Chronic catatonic schizophrenia |
| E1023 | Acute exacerbation of subchronic catatonic schizophrenia |
| E1024 | Acute exacerbation of chronic catatonic schizophrenia |
| E1025 | Catatonic schizophrenia in remission |
| E102z | Catatonic schizophrenia NOS |
| E103. | Paranoid schizophrenia |
| E1030 | Unspecified paranoid schizophrenia |
| E1031 | Subchronic paranoid schizophrenia |
| E1032 | Chronic paranoid schizophrenia |
| E1033 | Acute exacerbation of subchronic paranoid schizophrenia |
| E1034 | Acute exacerbation of chronic paranoid schizophrenia |
| E1035 | Paranoid schizophrenia in remission |
| E103z | Paranoid schizophrenia NOS |
| E104. | Acute schizophrenic episode |
| E105. | Latent schizophrenia |
| E1050 | Unspecified latent schizophrenia |
| E1051 | Subchronic latent schizophrenia |
| E1052 | Chronic latent schizophrenia |
| E1053 | Acute exacerbation of subchronic latent schizophrenia |
| E1054 | Acute exacerbation of chronic latent schizophrenia |
| E1055 | Latent schizophrenia in remission |
| E105z | Latent schizophrenia NOS |

| | |
|---|---|
| E106. | Residual schizophrenia |
| E107. | Schizo-affective schizophrenia |
| E1070 | Unspecified schizo-affective schizophrenia |
| E1071 | Subchronic schizo-affective schizophrenia |
| E1072 | Chronic schizo-affective schizophrenia |
| E1073 | Acute exacerbation subchronic schizo-affective schizophrenia |
| E1074 | Acute exacerbation of chronic schizo-affective schizophrenia |
| E1075 | Schizo-affective schizophrenia in remission |
| E107z | Schizo-affective schizophrenia NOS |
| E10y. | Other schizophrenia |
| E10y0 | Atypical schizophrenia |
| E10y1 | Coenesthopathic schizophrenia |
| E10yz | Other schizophrenia NOS |
| E10z. | Schizophrenia NOS |
| E110. | Manic disorder, single episode |
| E1100 | Single manic episode, unspecified |
| E1101 | Single manic episode, mild |
| E1102 | Single manic episode, moderate |
| E1103 | Single manic episode, severe without mention of psychosis |
| E1104 | Single manic episode, severe, with psychosis |
| E1105 | Single manic episode in partial or unspecified remission |
| E1106 | Single manic episode in full remission |
| E110z | Manic disorder, single episode NOS |
| E111. | Recurrent manic episodes |
| E1110 | Recurrent manic episodes, unspecified |
| E1111 | Recurrent manic episodes, mild |
| E1112 | Recurrent manic episodes, moderate |
| E1113 | Recurrent manic episodes, severe without mention psychosis |
| E1114 | Recurrent manic episodes, severe, with psychosis |
| E1115 | Recurrent manic episodes, partial or unspecified remission |
| E1116 | Recurrent manic episodes, in full remission |
| E111z | Recurrent manic episode NOS |
| E1124 | Single major depressive episode, severe, with psychosis |
| E1134 | Recurrent major depressive episodes, severe, with psychosis |
| E114. | Bipolar affective disorder, currently manic |
| E1140 | Bipolar affective disorder, currently manic, unspecified |
| E1141 | Bipolar affective disorder, currently manic, mild |
| E1142 | Bipolar affective disorder, currently manic, moderate |
| E1143 | Bipolar affect disord, currently manic, severe, no psychosis |
| E1144 | Bipolar affect disord, currently manic,severe with psychosis |
| E1145 | Bipolar affect disord,currently manic, part/unspec remission |
| E1146 | Bipolar affective disorder, currently manic, full remission |
| E114z | Bipolar affective disorder, currently manic, NOS |
| E115. | Bipolar affective disorder, currently depressed |

| | |
|---|---|
| E1150 | Bipolar affective disorder, currently depressed, unspecified |
| E1151 | Bipolar affective disorder, currently depressed, mild |
| E1152 | Bipolar affective disorder, currently depressed, moderate |
| E1153 | Bipolar affect disord, now depressed, severe, no psychosis |
| E1154 | Bipolar affect disord, now depressed, severe with psychosis |
| E1155 | Bipolar affect disord, now depressed, part/unspec remission |
| E1156 | Bipolar affective disorder, now depressed, in full remission |
| E115z | Bipolar affective disorder, currently depressed, NOS |
| E116. | Mixed bipolar affective disorder |
| E1160 | Mixed bipolar affective disorder, unspecified |
| E1161 | Mixed bipolar affective disorder, mild |
| E1162 | Mixed bipolar affective disorder, moderate |
| E1163 | Mixed bipolar affective disorder, severe, without psychosis |
| E1164 | Mixed bipolar affective disorder, severe, with psychosis |
| E1165 | Mixed bipolar affective disorder, partial/unspec remission |
| E1166 | Mixed bipolar affective disorder, in full remission |
| E116z | Mixed bipolar affective disorder, NOS |
| E117. | Unspecified bipolar affective disorder |
| E1170 | Unspecified bipolar affective disorder, unspecified |
| E1171 | Unspecified bipolar affective disorder, mild |
| E1172 | Unspecified bipolar affective disorder, moderate |
| E1173 | Unspecified bipolar affective disorder, severe, no psychosis |
| E1174 | Unspecified bipolar affective disorder,severe with psychosis |
| E1175 | Unspecified bipolar affect disord, partial/unspec remission |
| E1176 | Unspecified bipolar affective disorder, in full remission |
| E117z | Unspecified bipolar affective disorder, NOS |
| E11y. | Other and unspecified manic-depressive psychoses |
| E11y0 | Unspecified manic-depressive psychoses |
| E11y1 | Atypical manic disorder |
| E11y3 | Other mixed manic-depressive psychoses |
| E11yz | Other and unspecified manic-depressive psychoses NOS |
| E11z. | Other and unspecified affective psychoses |
| E11z0 | Unspecified affective psychoses NOS |
| E11zz | Other affective psychosis NOS |
| E12.. | Paranoid states |
| E120. | Simple paranoid state |
| E121. | Chronic paranoid psychosis |
| E122. | Paraphrenia |
| E123. | Shared paranoid disorder |
| E12y. | Other paranoid states |
| E12y0 | Paranoia querulans |
| E12yz | Other paranoid states NOS |
| E12z. | Paranoid psychosis NOS |
| E13.. | Other nonorganic psychoses |

| | |
|---|---|
| E130. | Reactive depressive psychosis |
| E131. | Acute hysterical psychosis |
| E132. | Reactive confusion |
| E133. | Acute paranoid reaction |
| E134. | Psychogenic paranoid psychosis |
| E13y. | Other reactive psychoses |
| E13y0 | Psychogenic stupor |
| E13y1 | Brief reactive psychosis |
| E13yz | Other reactive psychoses NOS |
| E13z. | Nonorganic psychosis NOS |
| E2122 | Schizotypal personality |
| Eu2.. | [X]Schizophrenia, schizotypal and delusional disorders |
| Eu20. | [X]Schizophrenia |
| Eu200 | [X]Paranoid schizophrenia |
| Eu201 | [X]Hebephrenic schizophrenia |
| Eu202 | [X]Catatonic schizophrenia |
| Eu203 | [X]Undifferentiated schizophrenia |
| Eu204 | [X]Post-schizophrenic depression |
| Eu205 | [X]Residual schizophrenia |
| Eu206 | [X]Simple schizophrenia |
| Eu20y | [X]Other schizophrenia |
| Eu20z | [X]Schizophrenia, unspecified |
| Eu21. | [X]Schizotypal disorder |
| Eu22. | [X]Persistent delusional disorders |
| Eu220 | [X]Delusional disorder |
| Eu221 | [X]Delusional misidentification syndrome |
| Eu222 | [X]Cotard syndrome |
| Eu223 | [X]Paranoid state in remission |
| Eu22y | [X]Other persistent delusional disorders |
| Eu22z | [X]Persistent delusional disorder, unspecified |
| Eu23. | [X]Acute and transient psychotic disorders |
| Eu230 | [X]Acute polymorphic psychot disord without symp of schizoph |
| Eu231 | [X]Acute polymorphic psychot disord with symp of schizophren |
| Eu232 | [X]Acute schizophrenia-like psychotic disorder |
| Eu233 | [X]Other acute predominantly delusional psychotic disorders |
| Eu23y | [X]Other acute and transient psychotic disorders |
| Eu23z | [X]Acute and transient psychotic disorder, unspecified |
| Eu24. | [X]Induced delusional disorder |
| Eu25. | [X]Schizoaffective disorders |
| Eu250 | [X]Schizoaffective disorder, manic type |
| Eu251 | [X]Schizoaffective disorder, depressive type |
| Eu252 | [X]Schizoaffective disorder, mixed type |
| Eu25y | [X]Other schizoaffective disorders |
| Eu25z | [X]Schizoaffective disorder, unspecified |

| | |
|---|---|
| Eu26. | [X]Nonorganic psychosis in remission |
| Eu2y. | [X]Other nonorganic psychotic disorders |
| Eu2z. | [X]Unspecified nonorganic psychosis |
| Eu30. | [X]Manic episode |
| Eu300 | [X]Hypomania |
| Eu301 | [X]Mania without psychotic symptoms |
| Eu302 | [X]Mania with psychotic symptoms |
| Eu30y | [X]Other manic episodes |
| Eu30z | [X]Manic episode, unspecified |
| Eu31. | [X]Bipolar affective disorder |
| Eu310 | [X]Bipolar affective disorder, current episode hypomanic |
| Eu311 | [X]Bipolar affect disorder cur epi manic wout psychotic symp |
| Eu312 | [X]Bipolar affect disorder cur epi manic with psychotic symp |
| Eu313 | [X]Bipolar affect disorder cur epi mild or moderate depressn |
| Eu314 | [X]Bipol aff disord, curr epis sev depress, no psychot symp |
| Eu315 | [X]Bipolar affect dis cur epi severe depres with psyc symp |
| Eu316 | [X]Bipolar affective disorder, current episode mixed |
| Eu317 | [X]Bipolar affective disorder, currently in remission |
| Eu318 | [X]Bipolar affective disorder type I |
| Eu319 | [X]Bipolar affective disorder type II |
| Eu31y | [X]Other bipolar affective disorders |
| Eu31z | [X]Bipolar affective disorder, unspecified |
| Eu323 | [X]Severe depressive episode with psychotic symptoms |
| Eu333 | [X]Recurrent depress disorder cur epi severe with psyc symp |
