## Supplementary material codes 4 for "How does the local area deprivation influence life chances for children in poverty in Wales: A record linkage cohort study"

### Table 1: Substance misuse related ICD10 codes

| **ICD10 codes** | **Descriptions** |
|---|---|
| F11 | Mental and behavioural disorders due to use of opioids |
| F110 | Mental and behavioural disorders due to use of opioids |
| F111 | Mental and behavioural disorders due to use of opioids |
| F112 | Mental and behavioural disorders due to use of opioids |
| F113 | Mental and behavioural disorders due to use of opioids |
| F114 | Mental and behavioural disorders due to use of opioids |
| F115 | Mental and behavioural disorders due to use of opioids |
| F116 | Mental and behavioural disorders due to use of opioids |
| F117 | Mental and behavioural disorders due to use of opioids |
| F118 | Mental and behavioural disorders due to use of opioids |
| F119 | Mental and behavioural disorders due to use of opioids |
| F12 | Mental and behavioural disorders due to use of cannabinoids |
| F120 | Mental and behavioural disorders due to use of cannabinoids |
| F121 | Mental and behavioural disorders due to use of cannabinoids |
| F122 | Mental and behavioural disorders due to use of cannabinoids |
| F123 | Mental and behavioural disorders due to use of cannabinoids |
| F124 | Mental and behavioural disorders due to use of cannabinoids |
| F125 | Mental and behavioural disorders due to use of cannabinoids |
| F126 | Mental and behavioural disorders due to use of cannabinoids |
| F127 | Mental and behavioural disorders due to use of cannabinoids |
| F128 | Mental and behavioural disorders due to use of cannabinoids |
| F129 | Mental and behavioural disorders due to use of cannabinoids |
| F13 | Mental and behavioural disorders due to use of sedatives or hypnotics |
| F130 | Mental and behavioural disorders due to use of sedatives or hypnotics |
| F131 | Mental and behavioural disorders due to use of sedatives or hypnotics |
| F132 | Mental and behavioural disorders due to use of sedatives or hypnotics |
| F133 | Mental and behavioural disorders due to use of sedatives or hypnotics |
| F134 | Mental and behavioural disorders due to use of sedatives or hypnotics |
| F135 | Mental and behavioural disorders due to use of sedatives or hypnotics |
| F136 | Mental and behavioural disorders due to use of sedatives or hypnotics |
| F137 | Mental and behavioural disorders due to use of sedatives or hypnotics |
| F138 | Mental and behavioural disorders due to use of sedatives or hypnotics |
| F139 | Mental and behavioural disorders due to use of sedatives or hypnotics |
| F14 | Mental and behavioural disorders due to use of cocaine |
| F140 | Mental and behavioural disorders due to use of cocaine |
| F141 | Mental and behavioural disorders due to use of cocaine |
| F142 | Mental and behavioural disorders due to use of cocaine |
| F143 | Mental and behavioural disorders due to use of cocaine |
| F144 | Mental and behavioural disorders due to use of cocaine |
| F145 | Mental and behavioural disorders due to use of cocaine |
| F146 | Mental and behavioural disorders due to use of cocaine |

| | |
|---|---|
| F147 | Mental and behavioural disorders due to use of cocaine |
| F148 | Mental and behavioural disorders due to use of cocaine |
| F149 | Mental and behavioural disorders due to use of cocaine |
| F15 | Mental and behavioural disorders due to use of other stimulants, including caffeine |
| F150 | Mental and behavioural disorders due to use of other stimulants, including caffeine |
| F151 | Mental and behavioural disorders due to use of other stimulants, including caffeine |
| F152 | Mental and behavioural disorders due to use of other stimulants, including caffeine |
| F153 | Mental and behavioural disorders due to use of other stimulants, including caffeine |
| F154 | Mental and behavioural disorders due to use of other stimulants, including caffeine |
| F155 | Mental and behavioural disorders due to use of other stimulants, including caffeine |
| F156 | Mental and behavioural disorders due to use of other stimulants, including caffeine |
| F157 | Mental and behavioural disorders due to use of other stimulants, including caffeine |
| F158 | Mental and behavioural disorders due to use of other stimulants, including caffeine |
| F159 | Mental and behavioural disorders due to use of other stimulants, including caffeine |
| F16 | Mental and behavioural disorders due to use of hallucinogens |
| F160 | Mental and behavioural disorders due to use of hallucinogens |
| F161 | Mental and behavioural disorders due to use of hallucinogens |
| F162 | Mental and behavioural disorders due to use of hallucinogens |
| F163 | Mental and behavioural disorders due to use of hallucinogens |
| F164 | Mental and behavioural disorders due to use of hallucinogens |
| F165 | Mental and behavioural disorders due to use of hallucinogens |
| F166 | Mental and behavioural disorders due to use of hallucinogens |
| F167 | Mental and behavioural disorders due to use of hallucinogens |
| F168 | Mental and behavioural disorders due to use of hallucinogens |
| F169 | Mental and behavioural disorders due to use of hallucinogens |
| F18 | Mental and behavioural disorders due to use of volatile solvents |
| F180 | Mental and behavioural disorders due to use of volatile solvents |
| F181 | Mental and behavioural disorders due to use of volatile solvents |
| F182 | Mental and behavioural disorders due to use of volatile solvents |
| F183 | Mental and behavioural disorders due to use of volatile solvents |
| F184 | Mental and behavioural disorders due to use of volatile solvents |
| F185 | Mental and behavioural disorders due to use of volatile solvents |
| F186 | Mental and behavioural disorders due to use of volatile solvents |
| F187 | Mental and behavioural disorders due to use of volatile solvents |
| F188 | Mental and behavioural disorders due to use of volatile solvents |
| F189 | Mental and behavioural disorders due to use of volatile solvents |
| F19 | Mental and behavioural disorders due to multiple drug use and use of other psychoactive substances |
| F190 | Mental and behavioural disorders due to multiple drug use and use of other psychoactive substances |
| F191 | Mental and behavioural disorders due to multiple drug use and use of other psychoactive substances |
| F192 | Mental and behavioural disorders due to multiple drug use and use of other psychoactive substances |
| F193 | Mental and behavioural disorders due to multiple drug use and use of other psychoactive substances |

| | |
|---|---|
| F194 | Mental and behavioural disorders due to multiple drug use and use of other psychoactive substances |
| F195 | Mental and behavioural disorders due to multiple drug use and use of other psychoactive substances |
| F196 | Mental and behavioural disorders due to multiple drug use and use of other psychoactive substances |
| F197 | Mental and behavioural disorders due to multiple drug use and use of other psychoactive substances |
| F198 | Mental and behavioural disorders due to multiple drug use and use of other psychoactive substances |
| F199 | Mental and behavioural disorders due to multiple drug use and use of other psychoactive substances |
| O355 | Maternal care for (suspected) damage to fetus by drugs |
| R781 | Finding of opiate drug in blood |
| R782 | Finding of cocaine in blood |
| R783 | Finding of hallucinogen in blood |
| R784 | Finding of other drugs of addictive potential in blood |
| R785 | Finding of psychotropic drug in blood |
| T40 | Poisoning by narcotics and psychodysleptics [hallucinogens] |
| T400 | Poisoning: Opium |
| T401 | Poisoning: Heroin |
| T402 | Poisoning: Other opioids |
| T403 | Poisoning: Methadone |
| T404 | Poisoning: Other synthetic narcotics |
| T405 | Poisoning: Cocaine |
| T406 | Poisoning: Other and unspecified narcotics |
| T407 | Poisoning: Cannabis (derivatives) |
| T408 | Poisoning: Lysergide [LSD] |
| T409 | Poisoning: Other and unspecified psychodysleptics [hallucinogens] |
| T436 | Poisoning: Psychostimulants with abuse potential |
| X42 | Accidental poisoning by and exposure to narcotics and psychodysleptics [hallucinogens], not elsewhere classified |
| X420 | Accidental poisoning by and exposure to narcotics and psychodysleptics [hallucinogens], not elsewhere classified |
| X421 | Accidental poisoning by and exposure to narcotics and psychodysleptics [hallucinogens], not elsewhere classified |
| X422 | Accidental poisoning by and exposure to narcotics and psychodysleptics [hallucinogens], not elsewhere classified |
| X423 | Accidental poisoning by and exposure to narcotics and psychodysleptics [hallucinogens], not elsewhere classified |
| X424 | Accidental poisoning by and exposure to narcotics and psychodysleptics [hallucinogens], not elsewhere classified |
| X425 | Accidental poisoning by and exposure to narcotics and psychodysleptics [hallucinogens], not elsewhere classified |
| X426 | Accidental poisoning by and exposure to narcotics and psychodysleptics [hallucinogens], not elsewhere classified |
| X427 | Accidental poisoning by and exposure to narcotics and psychodysleptics [hallucinogens], not elsewhere classified |

| | |
|---|---|
| X428 | Accidental poisoning by and exposure to narcotics and psychodysleptics [hallucinogens], not elsewhere classified |
| X429 | Accidental poisoning by and exposure to narcotics and psychodysleptics [hallucinogens], not elsewhere classified |
| X62 | Intentional self-poisoning by and exposure to narcotics and psychodysleptics [hallucinogens], not elsewhere classified |
| X620 | Intentional self-poisoning by and exposure to narcotics and psychodysleptics [hallucinogens], not elsewhere classified |
| X621 | Intentional self-poisoning by and exposure to narcotics and psychodysleptics [hallucinogens], not elsewhere classified |
| X622 | Intentional self-poisoning by and exposure to narcotics and psychodysleptics [hallucinogens], not elsewhere classified |
| X623 | Intentional self-poisoning by and exposure to narcotics and psychodysleptics [hallucinogens], not elsewhere classified |
| X624 | Intentional self-poisoning by and exposure to narcotics and psychodysleptics [hallucinogens], not elsewhere classified |
| X625 | Intentional self-poisoning by and exposure to narcotics and psychodysleptics [hallucinogens], not elsewhere classified |
| X626 | Intentional self-poisoning by and exposure to narcotics and psychodysleptics [hallucinogens], not elsewhere classified |
| X627 | Intentional self-poisoning by and exposure to narcotics and psychodysleptics [hallucinogens], not elsewhere classified |
| X628 | Intentional self-poisoning by and exposure to narcotics and psychodysleptics [hallucinogens], not elsewhere classified |
| X629 | Intentional self-poisoning by and exposure to narcotics and psychodysleptics [hallucinogens], not elsewhere classified |
| Y12 | Poisoning by and exposure to narcotics and psychodysleptics [hallucinogens], not elsewhere classified, undetermined intent |
| Y120 | Poisoning by and exposure to narcotics and psychodysleptics [hallucinogens], not elsewhere classified, undetermined intent |
| Y121 | Poisoning by and exposure to narcotics and psychodysleptics [hallucinogens], not elsewhere classified, undetermined intent |
| Y122 | Poisoning by and exposure to narcotics and psychodysleptics [hallucinogens], not elsewhere classified, undetermined intent |
| Y123 | Poisoning by and exposure to narcotics and psychodysleptics [hallucinogens], not elsewhere classified, undetermined intent |
| Y124 | Poisoning by and exposure to narcotics and psychodysleptics [hallucinogens], not elsewhere classified, undetermined intent |
| Y125 | Poisoning by and exposure to narcotics and psychodysleptics [hallucinogens], not elsewhere classified, undetermined intent |
| Y126 | Poisoning by and exposure to narcotics and psychodysleptics [hallucinogens], not elsewhere classified, undetermined intent |
| Y127 | Poisoning by and exposure to narcotics and psychodysleptics [hallucinogens], not elsewhere classified, undetermined intent |
| Y128 | Poisoning by and exposure to narcotics and psychodysleptics [hallucinogens], not elsewhere classified, undetermined intent |
| Y129 | Poisoning by and exposure to narcotics and psychodysleptics [hallucinogens], not elsewhere classified, undetermined intent |
| Z503 | Drug rehabilitation |
| Z715 | Drug abuse counselling and surveillance |

| | |
|---|---|
| Z722 | Drug use |

### Table 2: Substance misuse related READ codes

| READ codes | Descriptions |
|---|---|
| 13c.. | Drug user |
| 13c0. | Injecting drug user |
| 13c1. | Intravenous drug user |
| 13c2. | Never injecting drug user |
| 13c3. | Intramuscular drug user |
| 13c4. | Intranasal drug user |
| 13c5. | Substance misuse increased |
| 13c6. | Substance misuse decreased |
| 13c7. | Current drug user |
| 13c8. | Reduced drugs misuse |
| 13c9. | Subcutaneous drug user |
| 13cA. | Smokes drugs |
| 13cB. | Misuses drugs orally |
| 13cC. | Continuous use of drugs |
| 13cD. | Episodic use of drugs |
| 13cE. | Prolong high dose use cannabis |
| 13cF. | Preoccup with substance misuse |
| 13cG. | Drug tolerance |
| 13cG0 | Opioid tolerant |
| 13cG1 | Opioid naive |
| 13cH. | Persistent substance misuse |
| 13cJ. | Previously injecting drug user |
| 13cK. | Current non recreat drug user |
| 13cL. | Has never injected drugs |
| 13cM. | Substance misuse |
| 13cM0 | Novl psychactve sbstnce misuse |
| 13cM1 | Opioid analgesic dependence |
| 13cN. | Has nvr shrd drg injctn equipt |
| 13cQ. | Behavioural tolerance to drug |
| 13cR. | Physical tolerance to drug |
| 13cS. | Psychological drug tolerance |
| 13cT. | Reverse tolerance to drug |
| 1463 | H/O: drug dependency |
| 146C. | Failed heroin detoxification |
| 146E. | H/O: recreational drug use |
| 146F. | H/O: drug abuse |
| 1P30. | Compul uncontrollable drug tak |
| 1P31. | Compulsive drug taking |
| 1T... | History of substance misuse |

| | |
|---|---|
| 1T0.. | H/O heroin misuse |
| 1T00. | H/O daily heroin misuse |
| 1T01. | H/O weekly heroin misuse |
| 1T02. | Prev history of heroin misuse |
| 1T03. | H/O infrequent heroin misuse |
| 1T1.. | H/O methadone misuse |
| 1T10. | H/O daily methadone misuse |
| 1T11. | H/O weekly methadone misuse |
| 1T12. | H/O infrequent methadone misus |
| 1T13. | Prev history methadone misuse |
| 1T2.. | H/O ecstasy misuse |
| 1T20. | H/O daily ecstasy misuse |
| 1T21. | H/O weekly ecstasy misuse |
| 1T22. | H/O infrequent ecstasy misuse |
| 1T23. | Prev history of ecstasy misuse |
| 1T3.. | H/O benzodiazepine misuse |
| 1T30. | H/O daily benzodiazepin misuse |
| 1T31. | H/O weekly benzodiazep misuse |
| 1T32. | H/O infreq benzodiazep misuse |
| 1T33. | Prev H/O benzodiazepine misuse |
| 1T4.. | H/O amphetamine misuse |
| 1T40. | H/O daily amphetamine misuse |
| 1T41. | H/O weekly amphetamine misuse |
| 1T42. | H/O infrequent amphetam misuse |
| 1T43. | Prev H/O amphetamine misuse |
| 1T5.. | H/O cocaine misuse |
| 1T50. | H/O daily cocaine misuse |
| 1T51. | H/O weekly cocaine misuse |
| 1T52. | H/O infrequent cocaine misuse |
| 1T53. | Prev H/O cocaine misuse |
| 1T6.. | H/O crack cocaine misuse |
| 1T60. | H/O daily crack cocaine misuse |
| 1T61. | H/O weekly crack cocain misuse |
| 1T62. | H/O infrequ crack cocain misus |
| 1T63. | Prev H/O crack cocaine misuse |
| 1T7.. | H/O hallucinogen misuse |
| 1T70. | H/O daily hallucinogen misuse |
| 1T71. | H/O weekly hallucinogen misuse |
| 1T72. | H/O infrequ hallucinog misuse |
| 1T73. | Prev H/O hallucinogen misuse |
| 1T8.. | H/O cannabis misuse |
| 1T80. | H/O daily cannabis misuse |
| 1T81. | H/O weekly cannabis misuse |
| 1T82. | H/O infrequent cannabis misuse |

| | |
|---|---|
| 1T83. | Prev H/O cannabis misuse |
| 1T9.. | H/O solvent misuse |
| 1T90. | H/O daily solvent misuse |
| 1T91. | H/O weekly solvent misuse |
| 1T92. | H/O infrequent solvent misuse |
| 1T93. | Prev history of solvent misuse |
| 1TA.. | H/O barbiturate misuse |
| 1TA0. | H/O daily barbiturate misuse |
| 1TA1. | H/O weekly barbiturate misuse |
| 1TA2. | H/O infrequ barbiturate misuse |
| 1TA3. | Prev H/O barbiturate misuse |
| 1TB.. | H/O major tranquilliser misuse |
| 1TB0. | H/O daily maj tranquilli misus |
| 1TB1. | H/O weekl maj tranquilli misus |
| 1TB2. | H/O infreq maj trnquillis miss |
| 1TB3. | Prev H/O major tranq misuse |
| 1TC.. | H/O anti-depressant misuse |
| 1TC0. | H/O daily anti-depress misuse |
| 1TC1. | H/O weekly anti-depress misuse |
| 1TC2. | H/O infreq anti-depress misuse |
| 1TC3. | Prev H/O anti-depressnt misuse |
| 1TD.. | H/O opiate misuse |
| 1TD0. | H/O daily opiate misuse |
| 1TD1. | H/O weekly opiate misuse |
| 1TD2. | H/O infrequent opiate misuse |
| 1TD3. | Prev history of opiate misuse |
| 1TE.. | Uses heroin on top subst ther |
| 1TF.. | Dsnt use heroin top subst ther |
| 1TG.. | H/O nov psychoact subst misuse |
| 1V... | Drug misuse behaviour |
| 1V0.. | Misuses drugs |
| 1V00. | Occasional drug user |
| 1V01. | Long-term drug misuser |
| 1V02. | Poly-drug misuser |
| 1V03. | Misuses drugs sublingually |
| 1V04. | Misuses drugs rectally |
| 1V05. | Misuses drugs vaginally |
| 1V06. | Uses drug paraphernalia |
| 1V07. | Notified addict |
| 1V08. | Smokes drugs in cigarette form |
| 1V09. | Smokes drugs through a pipe |
| 1V0A. | Chases the dragon |
| 1V0B. | Sniffs drugs |
| 1V0C. | Drug addict |

| | |
|---|---|
| 1V0D. | Am spent per day on drug habit |
| 1V0E. | Health prob sec to drug misuse |
| 1V1.. | Time devotd drug rel activties |
| 1V10. | Time spent obtaining drugs |
| 1V11. | Time spent taking drugs |
| 1V12. | Time spent recover from drugs |
| 1V2.. | Frequency of drug misuse |
| 1V22. | Age at starting drug misuse |
| 1V23. | Time since stopped drug misuse |
| 1V24. | Total time drugs misused |
| 1V26. | Misused drugs in past |
| 1V3.. | Drug injection behaviour |
| 1V30. | Injects drugs subcutaneously |
| 1V31. | Injects drugs intramuscularly |
| 1V32. | Neck injector |
| 1V33. | Groin injector |
| 1V34. | Does not inject drugs |
| 1V35. | Shares drug equipment |
| 1V36. | Frontloading |
| 1V37. | Drug inject equipment hygiene |
| 1V38. | Sharing drug inject equipment |
| 1V3A. | Not share drug inject equipmen |
| 1V3B. | Shares syringes |
| 1V3C. | Shares needles |
| 1V3D. | Cleaning of needles |
| 1V3E. | Cleans own needles |
| 1V3F. | Cleans needles with bleach |
| 1V3G. | Does not clean needles |
| 1V3H. | Obtains clean needles |
| 1V3J. | Uses needle exchange scheme |
| 1V3K. | Obtains clean syringes |
| 1V3L. | Needle syringe exch scheme use |
| 1V3M. | Needle + syringe exch not used |
| 1V3N. | Needle and syringe exch used |
| 1V4.. | Priority of drug activity |
| 1V40. | No priority to drug activities |
| 1V41. | Priority to drug activities |
| 1V42. | Drug priority ov social obligs |
| 1V43. | Drug priority over family |
| 1V44. | Drug priority ov finance oblig |
| 1V5.. | Routine drug-related activity |
| 1V50. | No routine of drug activities |
| 1V51. | Has routine of drug activities |
| 1V52. | Same drug routine every day |

| | |
|---|---|
| 1V53. | Drug-related rituals |
| 1V54. | Follows drug-related rituals |
| 1V55. | Not follow drug-relate rituals |
| 1V6.. | Drug-relat offending behaviour |
| 1V64. | Illicit drug use |
| 1V65. | Heroin misuse |
| 1V66. | Ecstasy misuse |
| 677T. | Subst misuse structurd counsel |
| 7P220 | Delivery rehab drug addiction |
| 8AA.. | Drug abuse monitoring |
| 8B23. | Drug addiction therapy |
| 8B230 | Drug add maint ther naltrexone |
| 8B231 | Drug add maint ther lofexidine |
| 8B2M. | Buprenorphine maintenance ther |
| 8B2N. | Drug add detox ther methadone |
| 8B2P. | Drug add maint ther methadone |
| 8B2Q. | Drug add maint ther buprenorph |
| 8B2R. | Drug add detox ther buprenorph |
| 8B2S. | Opioid agonist substitut thera |
| 8B2T. | Opioid antagonist therapy |
| 8BA9. | Detoxification dependence drug |
| 8BAW. | Drug depen self detoxification |
| 8BAX. | Drug depen home detoxification |
| 8BAc. | Subs mis mgt stop - self withd |
| 8BAd. | Opiate dependence detoxificatn |
| 8BAt. | Drug relapse prevention |
| 8BAv. | Drug harm reduction programme |
| 8BAx. | Drug twelve step programme |
| 8BE.. | Maintenance therapy |
| 8BE0. | Reinduct methadone maint thera |
| 8BE1. | Reinduct buprenorph maint ther |
| 8CR9. | Benzodiazepi clinical mgt plan |
| 8FB.. | Drug rehabilitation |
| 8FB0. | Drug detox programme completed |
| 8H7x. | Refer to drug abuse counsellor |
| 8HHL. | Ref to comm drug dependen team |
| 8HHd. | Referral to drug treatment cen |
| 8HHe. | Referral to com drug alco team |
| 8Hh1. | Self refer substanc misus serv |
| 8HkF. | Refer substance misuse service |
| 8Hl5. | Referral to drugs therapist |
| 8Hl6. | Referral to drugs worker |
| 8Hq.. | Admsn substnc misuse detox cnt |
| 8I2N. | Drug depend home detox contra |

| | |
|---|---|
| 8IE7. | Substance misuse assess declin |
| 9G2.. | Drug addiction notification |
| 9G21. | Drug addict notific to CMO |
| 9G22. | Drug addict re-notific due |
| 9G23. | Drug addict re-notif to CMO |
| 9G2Z. | Drug addiction notif NOS |
| 9HC.. | Substance misuse monitoring |
| 9HC0. | Initial substance misuse asses |
| 9HC1. | Follow up substa misuse assess |
| 9HC2. | Subst mis clin man plan agreed |
| 9HC3. | Subst mis clin man plan review |
| 9HC4. | Sub misuse treatment withdrawn |
| 9HC5. | Sub misus treat prog completed |
| 9HC6. | Substance misuse treatm declin |
| 9HC7. | Subst misuse treat not availbl |
| 9HC8. | Decl to give subst misuse hist |
| 9HC9. | Snc mse tmnt gvn othr hcr prdr |
| 9HCA. | Sbstnce misuse mntr 6 mnth rvw |
| 9HCB. | Substance misuse mntr annl rvw |
| 9HCC. | On substance misuse programme |
| 9N0Z. | Seen in drug rehab centre |
| 9N1yJ | Seen in drug misuse clinic |
| 9N6a. | Refer by drug statutor service |
| 9N6b. | Ref by drug non-statutory serv |
| 9N6g. | Refer by syringe excha service |
| 9NN1. | Under care community drug team |
| 9NX2. | In-house subs misuse treatment |
| 9NdN. | Declnd consnt notif drug misus |
| 9No5. | Seen in substance misuse clinc |
| 9k5.. | Drug misuse - enhan serv admin |
| 9k50. | Drug misuse - enh serv complet |
| 9k51. | Share care drug misu trt - ESA |
| 9k52. | Drug misus trt prim care - ESA |
| 9k53. | Phrmcy attend drug misus - ESA |
| 9kS.. | Drug mis asse decl - enha serv |
| E02.. | Drug psychoses |
| E020. | Drug withdrawal syndrome |
| E021. | Drug-induced paranoia/hallucin |
| E0210 | Drug-induced paranoid state |
| E0211 | Drug-induced hallucinosis |
| E021z | Drug-induc.paranoia/halluc NOS |
| E022. | Pathological drug intoxication |
| E02y. | Other drug psychoses |
| E02y0 | Drug-induced delirium |

| | |
|---|---|
| E02y1 | Drug-induced dementia |
| E02y2 | Drug-induced amnestic syndrome |
| E02y3 | Drug-induced depressive state |
| E02y4 | Drug-induced personality dis. |
| E02yz | Other drug psychoses NOS |
| E02z. | Drug psychosis NOS |
| E24.. | Drug dependence |
| E240. | Opioid type drug dependence |
| E2400 | Opioid dependence-unspecified |
| E2401 | Opioid dependence-continuous |
| E2402 | Opioid dependence - episodic |
| E2403 | Opioid dependence-in remission |
| E240z | Opioid drug dependence NOS |
| E241. | Hypnotic/anxiolytic dependence |
| E2410 | Hypnotic/anxiol.depend.-unspec |
| E2411 | Hypnot/anxiol.dep.-continuous |
| E2412 | Hypnot/anxiol.dep.-episodic |
| E2413 | Hypnot/anxiol.dep-in remission |
| E241z | Hypnotic/anxiolytic depend.NOS |
| E242. | Cocaine type drug dependence |
| E2420 | Cocaine dependence-unspecified |
| E2421 | Cocaine dependence-continuous |
| E2422 | Cocaine dependence-episodic |
| E2423 | Cocaine depend. - in remission |
| E242z | Cocaine drug dependence NOS |
| E243. | Cannabis type drug dependence |
| E2430 | Cannabis dependence-unspecif. |
| E2431 | Cannabis dependence-continuous |
| E2432 | Cannabis dependence-episodic |
| E2433 | Cannabis depend.- in remission |
| E243z | Cannabis drug dependence NOS |
| E244. | Amphetamine/psychostim.depend. |
| E2440 | Amphetamine depend.-unspecif. |
| E2441 | Amphetamine depend.-continuous |
| E2442 | Amphetamine depend.-episodic |
| E2443 | Amphetamine dep.-in remission |
| E244z | Amphetamine dependence NOS |
| E245. | Hallucinogen dependence |
| E2450 | Hallucinogen depend.-unspecif. |
| E2451 | Hallucinogen depend-continuous |
| E2452 | Hallucinogen depend.-episodic |
| E2453 | Hallucinogen dep.-in remission |
| E245z | Hallucinogen dependence NOS |
| E246. | Glue sniffing dependence |

| | |
|---|---|
| E2460 | Glue sniffing - unspecified |
| E2461 | Glue sniffing - continuous |
| E2462 | Glue sniffing - episodic |
| E2463 | Glue sniffing - in remission |
| E246z | Glue sniffing dependence NOS |
| E247. | Other specified drug dependen. |
| E2470 | Other drug dependence unspecif |
| E2471 | Other drug depend.-continuous |
| E2472 | Other drug depend.-episodic |
| E2473 | Other drug dep.-in remission |
| E247z | Other drug dependence NOS |
| E248. | Combined opioid+other drug dep |
| E2480 | Opioid+other drug dep. unspec. |
| E2481 | Continuous opioid+other depen. |
| E2482 | Episodic opioid+other depend. |
| E2483 | In remission-opioid+other dep. |
| E248z | Opioid+other drug depend. NOS |
| E249. | Combined drug dep. excl.opioid |
| E2490 | Comb.drug dep ex opioid-unspec |
| E2491 | Comb.drug dep ex opioid-contin |
| E2492 | Comb.drug dep ex opioid-episod |
| E2493 | Comb.drug dep ex opioid-in rem |
| E249z | Comb.drug dep ex opioid NOS |
| E24A. | Ecstasy type drug dependence |
| E24z. | Drug dependence NOS |
| E25.. | Nondependent abuse of drugs |
| E252. | Nondependent cannabis abuse |
| E2520 | Nondep cannabis abuse - unspec |
| E2521 | Nondep cannabis abuse - contin |
| E2522 | Nondep cannabis abuse - episod |
| E2523 | Nondep cannabis abuse in remis |
| E252z | Nondep cannabis abuse NOS |
| E253. | Nondependen hallucinogen abuse |
| E2530 | Nondep hallucinogen abuse-unsp |
| E2531 | Nondep hallucinogen abuse-cont |
| E2532 | Nondep hallucinogen abuse-epis |
| E2533 | Nondep hallucin abuse-in remis |
| E253z | Nondep hallucinogen abuse NOS |
| E254. | Nondep hypnot/anxiolytic abuse |
| E2540 | Nondep hypnot/anxio.abuse-unsp |
| E2541 | Nondep hypnot/anxio.abuse-cont |
| E2542 | Nondep hypnot/anxio.abuse-epis |
| E2543 | Nondep hypn/anxio.abuse-in rem |
| E254z | Nondep hypnot/anxiol abuse NOS |

| | |
|---|---|
| E255. | Nondependent opioid abuse |
| E2550 | Nondep opioid abuse - unspecif |
| E2551 | Nondep opioid abuse - continuo |
| E2552 | Nondep opioid abuse - episodic |
| E2553 | Nondep opioid abuse - in remis |
| E255z | Nondependent opioid abuse NOS |
| E256. | Nondependent cocaine abuse |
| E2560 | Nondep cocaine abuse - unspec. |
| E2561 | Nondep cocaine abuse - contin. |
| E2562 | Nondep cocaine abuse - episod. |
| E2563 | Nondep cocaine abuse -in remis |
| E256z | Nondependent cocaine abuse NOS |
| E257. | Nondep amphetamine type abuse |
| E2570 | Nondep amphet type abuse -unsp |
| E2571 | Nondep amphet type abuse -cont |
| E2572 | Nondep amphet type abuse -epis |
| E2573 | Nondep amph. type abuse-in rem |
| E257z | Nondep amphet. type abuse NOS |
| E258. | Nondep antidepress type abuse |
| E2580 | Nondep antidep type abuse-unsp |
| E2581 | Nondep antidep type abuse-cont |
| E2582 | Nondep antidep type abuse-epis |
| E2583 | Nondep antidep tp abuse-in rem |
| E258z | Nondep antidep type abuse NOS |
| E259. | Nondependent mixed drug abuse |
| E2590 | Nondep mixed drug abuse-unspec |
| E2591 | Nondep mixed drug abuse-contin |
| E2592 | Nondep mixed drug abuse-episod |
| E2593 | Nondep mixed drug abuse-in rem |
| E2594 | Misuse of prescription drugs |
| E259z | Nondep mixed drug abuse NOS |
| E25y. | Nondependent other drug abuse |
| E25y0 | Nondep other drug abuse-unspec |
| E25y1 | Nondep other drug abuse-contin |
| E25y2 | Nondep other drug abuse-episod |
| E25y3 | Nondep other drug abuse-in rem |
| E25yz | Nondep other drug abuse NOS |
| E25z. | Misuse of drugs NOS |
| Eu1.. | [X]Mental dis, psychoact subst |
| Eu11. | [X]Mental dis due to opioids |
| Eu110 | [X]Acute opioid intoxication |
| Eu111 | [X]Harmful use of opioids |
| Eu112 | [X]Opioid dependence syndrome |
| Eu113 | [X]Opioid withthdrawal state |

| | |
|---|---|
| Eu114 | [X]Opioid withdrawal delirium |
| Eu115 | [X]Psychot dis due to opioids |
| Eu116 | [X]Amnesic synd due to opioids |
| Eu117 | [X]Resid psychotic due opioid |
| Eu11y | [X]Oth ment/beh dis due opioid |
| Eu11z | [X]Uns ment/beh dis due opioid |
| Eu12. | [X]Mental dis due cannabinoids |
| Eu120 | [X]Acute cannabis intoxication |
| Eu121 | [X]Harmful use of cannabis |
| Eu122 | [X]Cannabis dependence syndrom |
| Eu123 | [X]Cannabis withdrawal state |
| Eu124 | [X]Cannabis withdrawl delirium |
| Eu125 | [X]Psychot dis due to cannabis |
| Eu126 | [X]Amnesic synd due cannabis |
| Eu127 | [X]Resid psychot due cannabis |
| Eu12y | [X]Oth ment/beh dis cannabinds |
| Eu12z | [X]Unsp mnt/beh dis cannabinds |
| Eu13. | [X]Mental dis due sedat/hypnot |
| Eu130 | [X]Acute sedat/hypnotic intox |
| Eu131 | [X]Harmful use sedat/hypnotic |
| Eu132 | [X]Sedat/hypnotic depend syndr |
| Eu133 | [X]Sedat/hypnot withdraw state |
| Eu134 | [X]Sed/hypn withdraw delirium |
| Eu135 | [X]Psychot dis due sedat/hypn |
| Eu136 | [X]Amnesic synd due sedat/hypn |
| Eu137 | [X]Resid psychot due sed/hypn |
| Eu13y | [X]Oth ment/beh dis sed/hypnot |
| Eu13z | [X]Uns ment/beh dis sed/hypnot |
| Eu14. | [X]Mental dis due use cocaine |
| Eu140 | [X]Acute cocaine intoxication |
| Eu141 | [X]Harmful use of cocaine |
| Eu142 | [X]Cocaine dependence syndrome |
| Eu143 | [X]Cocaine withdrawal state |
| Eu144 | [X]Cocaine withdrawal delirium |
| Eu145 | [X]Psychot dis due to cocaine |
| Eu146 | [X]Amnesic synd due to cocaine |
| Eu147 | [X]Resid psychot due cocaine |
| Eu14y | [X]Ot ment/beh dis due cocaine |
| Eu14z | [X]Uns ment/bh dis due cocaine |
| Eu15. | [X]Ment dis oth stimul/caffein |
| Eu150 | [X]Acute intoxic oth stimulant |
| Eu151 | [X]Harmful use other stimulant |
| Eu152 | [X]Oth stimulant dependen synd |
| Eu153 | [X]Oth stimulant withdr state |

| | |
|---|---|
| Eu154 | [X]Oth stimulant withdr delir |
| Eu155 | [X]Psychotic dis oth stimulant |
| Eu156 | [X]Amnesic syndr oth stimulant |
| Eu157 | [X]Resid psychot oth stimulant |
| Eu15y | [X]Oth ment/beh dis stimulant |
| Eu15z | [X]Uns ment/beh dis stimulant |
| Eu16. | [X]Mental disord hallucinogens |
| Eu160 | [X]Acute hallucinogen intoxic |
| Eu161 | [X]Harmful use hallucinogens |
| Eu162 | [X]Hallucinogen depend synd |
| Eu163 | [X]Hallucinogen withdraw state |
| Eu164 | [X]Hallucin withdraw delirium |
| Eu165 | [X]Psychotic due hallucinogen |
| Eu166 | [X]Amnesic synd due hallucinog |
| Eu167 | [X]Resid psychot hallucinogen |
| Eu16y | [X]Oth ment/beh dis hallucinog |
| Eu16z | [X]Uns ment/beh dis hallucinog |
| Eu18. | [X]Ment dis volatile solvents |
| Eu180 | [X]Acute solvent intoxication |
| Eu181 | [X]Harmful use of solvents |
| Eu182 | [X]Solvent dependence syndrome |
| Eu183 | [X]Solvent withdrawal state |
| Eu184 | [X]Solvent withdrawal delirium |
| Eu185 | [X]Psychotic dis due solvent |
| Eu186 | [X]Amnesic syndr due solvent |
| Eu187 | [X]Resid psychotic due solvent |
| Eu18y | [X]Ot ment/beh dis due solvent |
| Eu18z | [X]Uns ment/beh due solvent |
| Eu19. | [X]Ment disord multi drug use |
| Eu190 | [X]Acute intox multi drug use |
| Eu191 | [X]Harmful use multiple drugs |
| Eu192 | [X]Multiple drug dependence |
| Eu193 | [X]Multiple drug withdrawal |
| Eu194 | [X]Multi drug withdr delirium |
| Eu195 | [X]Psychotic due multi drugs |
| Eu196 | [X]Amnesic syn due multi drugs |
| Eu197 | [X]Resid psychotic multi drugs |
| Eu19y | [X]Ot ment/beh due multi drugs |
| Eu19z | [X]Un ment/beh due multi drugs |
| Eu1A. | [X]Men behav dis due crack coc |
| Eu1A0 | [X]Acute crack cocaine intoxic |
| Eu1A1 | [X]Harmful use crack cocaine |
| Eu1A2 | [X]Crack cocaine depend synd |
| Eu1A3 | [X]Crack cocaine withdraw stat |

| | |
|---|---|
| Eu1A4 | [X]Crack coc withdraw stat del |
| Eu1A5 | [X]Crack cocaine psychotic dis |
| Eu1A6 | [X]Crack cocaine amnesic synd |
| Eu1A7 | [X]Cra coc res late-on psy dis |
| Eu1Ay | [X]Crac coc other ment beh dis |
| Eu1Az | [X]Crack coc unsp men beh dis |
| L183. | Pregnancy+drug dependence |
| L1830 | Preg.+drug dependence unspecif |
| L1831 | Preg.+drug dependence-deliver. |
| L1832 | Preg.+drug depend-del+p/n comp |
| L1833 | Preg.+drug depend-not deliver. |
| L1834 | Preg.+drug depend.+p/n complic |
| L183z | Preg.+drug dependence NOS |
| L255. | Fetus+drug damage |
| L2550 | Fetus+drug damage unspecified |
| L2551 | Fetus+drug damage-delivered |
| L2552 | Fetus+drug damage+a/n problem |
| L255z | Fetus+drug damage NOS |
| R10B0 | [D]Finding of cocain in blood |
| R10B1 | [D]Find hallucinogen in blood |
| R10B2 | [D]Find psychotrop drug blood |
| R10B4 | [D]Finding, opiate drug in bld |
| Ryu86 | [X]Find ot drg addic poten,bld |
| SL50. | Opiate/narcotic poisoning |
| SL500 | Unspecified opium poisoning |
| SL501 | Heroin poisoning |
| SL502 | Methadone poisoning |
| SL50z | Opiate/narcotic poisoning NOS |
| SL850 | Cocaine poisoning |
| SL96. | Hallucinogen poisoning |
| SL960 | Cannabis poisoning |
| SL961 | Lysergide (LSD) poisoning |
| SL963 | Mescaline poisoning |
| SL964 | Psilocybin poisoning |
| SL96z | Hallucinogen poisoning NOS |
| SL97. | Psychostimulant poisoning |
| SL970 | Amfetamine poisoning |
| SL972 | Ecstasy poisoning |
| SL97z | Psychostimulant poisoning NOS |
| SyuFB | [X]Poisoning by other opioids |
| SyuFC | [X]Poisoning by oth synth narc |
| SyuFD | [X]Poisoning by oth/unsp narc |
| SyuFE | [X]Pois,oth/un psychodysl/hall |
| T800. | Accid.pois.- heroin |

| | |
|---|---|
| T801. | Accid.pois.- methadone |
| T8023 | Accid.pois.- opium |
| T841. | Accid.pois.- hallucinogens |
| T8410 | Accid.pois.- cannabis derivat. |
| T8413 | Accid.pois.- mescaline |
| T8414 | Accid.pois.- psilocin |
| T8415 | Accid.pois.- psilocybin |
| T842. | Accid.pois.- psychostimulants |
| T8420 | Accid.pois.- amphetamine |
| T8520 | Accid.pois.- cocaine |
| U1A5. | [X]Accident poisoning narcotic |
| U1A50 | [X]Acc poison narcotic home |
| U1A51 | [X]Ac pois narcotic res ins |
| U1A52 | [X]Ac pois narcotic pub ins |
| U1A53 | [X]Ac pois narcotic sport ar |
| U1A54 | [X]Ac pois narcotic on hway |
| U1A55 | [X]Ac pois narcotic trade ar |
| U1A56 | [X]Ac pois narcotic indus ar |
| U1A57 | [X]Ac pois narcotic on farm |
| U1A5y | [X]Ac pois narcotic OS place |
| U1A5z | [X]Ac pois narcotic unsp pl |
| U1A6. | [X]Acc poisoning hallucinogens |
| U1A60 | [X]Acc poison hallucinog home |
| U1A61 | [X]Ac pois hallucinog res ins |
| U1A62 | [X]Ac pois hallucinog pub ins |
| U1A63 | [X]Ac pois hallucinog sport ar |
| U1A64 | [X]Ac pois hallucinog on hway |
| U1A65 | [X]Ac pois hallucinog trade ar |
| U1A66 | [X]Ac pois hallucinog indus ar |
| U1A67 | [X]Ac pois hallucinog on farm |
| U1A6y | [X]Ac pois hallucinog OS place |
| U1A6z | [X]Ac pois hallucinog unsp pl |
| U205. | [X]Intent self poison narcotic |
| U2050 | [X]Self pois narcotic home |
| U2051 | [X]S/pois narcotic res ins |
| U2052 | [X]S/pois narcotic pub ins |
| U2053 | [X]S/pois narcotic sport ar |
| U2054 | [X]S/pois narcotic on hway |
| U2055 | [X]S/pois narcotic trade ar |
| U2056 | [X]S/pois narcotic indus ar |
| U2057 | [X]S/pois narcotic on farm |
| U205y | [X]S/pois narcotic OS place |
| U205z | [X]S/pois narcotic unsp pl |
| U206. | [X]Int s/poising hallucinogens |

| | |
|---|---|
| U2060 | [X]Self pois hallucinog home |
| U2061 | [X]S/pois hallucinog res ins |
| U2062 | [X]S/pois hallucinog pub ins |
| U2063 | [X]S/pois hallucinog sport ar |
| U2064 | [X]S/pois hallucinog on hway |
| U2065 | [X]S/pois hallucinog trade ar |
| U2066 | [X]S/pois hallucinog indus ar |
| U2067 | [X]S/pois hallucinog on farm |
| U206y | [X]S/pois hallucinog OS place |
| U206z | [X]S/pois hallucinog unsp pl |
| U405. | [X]Poisoning ?intent narcotic |
| U4050 | [X]Pois ?intent narcotic home |
| U4051 | [X]Pois ?int narcotic resid |
| U4052 | [X]Pois ?int narcotic pub ins |
| U4053 | [X]Pois ?int narcotic sport ar |
| U4054 | [X]Pois ?int narcotic on hway |
| U4055 | [X]Pois ?int narcotic trade ar |
| U4056 | [X]Pois ?int narcotic indus ar |
| U4057 | [X]Pois ?int narcotic on farm |
| U405y | [X]Pois ?int narcotic OS place |
| U405z | [X]Pois ?int narcotic unsp pl |
| U406. | [X]Poison ?intent hallucinogen |
| U4060 | [X]Pois ?intent hallucin home |
| U4061 | [X]Pois ?int hallucinog resid |
| U4062 | [X]Pois ?int hallucin pub ins |
| U4063 | [X]Pois ?int hallucin sport ar |
| U4064 | [X]Pois ?int hallucin on hway |
| U4065 | [X]Pois ?int hallucin trade ar |
| U4066 | [X]Pois ?int hallucin indus ar |
| U4067 | [X]Pois ?int hallucin on farm |
| U406y | [X]Pois ?int hallucin OS place |
| U406z | [X]Pois ?int hallucin unsp pl |
| ZV114 | [V]Pers hist subst abuse |
| ZV4K1 | [V]Drug use |
| ZV6D7 | [V]Drug abuse counsel+surveiln |
| dj36. | SUBUTEX 400micrograms s/l tabs |
| dj37. | SUBUTEX 2mg sublingual tablets |
| dj38. | SUBUTEX 8mg sublingual tablets |
| dj3D. | BUPRNRPHNE+NALOXN 2/0.5mg tabs |
| dj3E. | SUBOXONE 2mg/0.5mg s/l tabs |
| dj3F. | BUPRNRPHNE+NALOXN 8mg/2mg tabs |
| dj3G. | SUBOXONE 8mg/2mg s/l tabs |
| dj3K. | NATZON 400micrograms s/l tabs |
| dj3L. | NATZON 2mg sublingual tablets |

| | |
|---|---|
| dj3M. | NATZON 8mg sublingual tablets |
| dj3N. | GABUP 400micrograms s/l tabs |
| dj3O. | GABUP 1mg sublingual tablets |
| dj3P. | GABUP 2mg sublingual tablets |
| dj3Q. | GABUP 4mg sublingual tablets |
| dj3R. | GABUP 6mg sublingual tablets |
| dj3S. | GABUP 8mg sublingual tablets |
| dj3T. | BUPRENORPHINE 1mg s/l tabs |
| dj3U. | BUPRENORPHINE 4mg s/l tabs |
| dj3V. | BUPRENORPHINE 6mg s/l tabs |
| dj3c. | PREFIBIN 400mcg sublingual tab |
| dj3d. | PREFIBIN 2mg sublingual tabs |
| dj3e. | PREFIBIN 8mg sublingual tabs |
| dj3u. | BUPRENORPHINE 2mg s/l tabs |
| dj3v. | BUPRENORPHINE 8mg s/l tabs |
| djc.. | METHADONE HCL [ANALGESIC] |
| djc1. | PHYSEPTONE 5mg tablets |
| djc2. | PHYSEPTONE 10mg/1mL injection |
| djc3. | METHADONE 1mg/1mL mixture |
| djc4. | METHADONE HCL 50mg/5mL s/f liq |
| djc5. | MARTINDALE METHADONE DTF mixt |
| djc6. | METHODEX 1mg/1mL mixture |
| djc7. | METHADOSE 10mg/mL s/f liq |
| djc8. | METHADONE HCL 20mg/mL s/f liq |
| djc9. | METHADOSE 20mg/mL s/f liq |
| djcA. | METHADONE DILUENT liquid |
| djcB. | METHADOSE DILUENT liquid |
| djcC. | METHADONE 1mg/1mL s/f mixt |
| djcD. | METHAROSE 1mg/1mL s/f soln |
| djcE. | *PINADONE 1mg/1mL mixture |
| djcF. | *PINADONE 1mg/1mL s/f mixt |
| djcG. | PHYSEPTONE 20mg/2mL injection |
| djcH. | PHYSEPTONE 35mg/3.5mL inj |
| djcJ. | PHYSEPTONE 50mg/5mL injection |
| djcK. | PHYSEPTONE 1mg/1mL s/f mixture |
| djcL. | PHYSEPTONE 1mg/1mL mixture |
| djcM. | SYNASTONE 10mg/1mL injection |
| djcN. | SYNASTONE 20mg/2mL injection |
| djcO. | SYNASTONE 35mg/3.5mL injection |
| djcP. | SYNASTONE 50mg/5mL injection |
| djcQ. | SYNASTONE 50mg/2mL injection |
| djcR. | SYNASTONE 50mg/1mL injection |
| djcS. | PHYSEPTONE 50mg/2mL injection |
| djcT. | PHYSEPTONE 50mg/1mL injection |

| | |
|---|---|
| djcU. | EPTADONE 1mg/mL oral solution |
| djcV. | EPTADONE 5mg/mL oral solution |
| djcW. | EPTADONE 20mg/20mL oral soln |
| djcX. | EPTADONE 40mg/40mL oral soln |
| djcY. | EPTADONE 60mg/60mL oral soln |
| djcZ. | EPTADONE 100mg/20mL oral soln |
| djco. | METHADONE 20mg/20mL oral soln |
| djcp. | METHADONE 40mg/40mL oral soln |
| djcq. | METHADONE 60mg/60mL oral soln |
| djcr. | METHADONE 100mg/20mL oral soln |
| djcs. | METHADONE 5mg/mL oral solution |
| djct. | METHADONE HCL 50mg/2mL inj |
| djcu. | METHADONE HCL 50mg/1mL inj |
| djcv. | METHADONE HCL 20mg/2mL inj |
| djcw. | METHADONE HCL 35mg/3.5mL inj |
| djcx. | METHADONE HCL 50mg/5mL inj |
| djcy. | METHADONE HCL 5mg tablets |
| djcz. | METHADONE HCL 10mg/1mL inj |
| du2.. | NALTREXONE HYDROCHLORIDE |
| du21. | NALTREXONE HCL 50mg tablets |
| du22. | NALOREX 50mg tablets |
| du23. | OPIZONE 50mg tablets |
| du24. | ADEPEND 50mg tablets |
| du4.. | LOFEXIDINE HYDROCHLORIDE |
| du41. | BRITLOFEX 200mcg tablets |
| du42. | LOFEXIDINE HCL 200mcg tablets |
