## Supplementary material codes 5 for "How does the local area deprivation influence life chances for children in poverty in Wales: A record linkage cohort study"

### Table 1: Alcohol related ICD10 codes

| ICD10 Codes | Descriptions |
|---|---|
| E244 | Alcohol-induced pseudo-Cushing's syndrome |
| E512 | Wernicke's encephalopathy |
| F10 | Mental and behavioural disorders due to use of alcohol |
| F100 | Mental and behavioural disorders due to use of alcohol |
| F101 | Mental and behavioural disorders due to use of alcohol |
| F102 | Mental and behavioural disorders due to use of alcohol |
| F103 | Mental and behavioural disorders due to use of alcohol |
| F104 | Mental and behavioural disorders due to use of alcohol |
| F105 | Mental and behavioural disorders due to use of alcohol |
| F106 | Mental and behavioural disorders due to use of alcohol |
| F107 | Mental and behavioural disorders due to use of alcohol |
| F108 | Mental and behavioural disorders due to use of alcohol |
| F109 | Mental and behavioural disorders due to use of alcohol |
| G312 | Degeneration of nervous system due to alcohol |
| G621 | Alcoholic polyneuropathy |
| G721 | Alcoholic myopathy |
| I426 | Alcoholic cardiomyopathy |
| K292 | Alcoholic gastritis |
| K70 | Alcoholic liver disease |
| K700 | Alcoholic fatty liver |
| K701 | Alcoholic hepatitis |
| K702 | Alcoholic fibrosis and sclerosis of liver |
| K703 | Alcoholic cirrhosis of liver |
| K704 | Alcoholic hepatic failure |
| K709 | Alcoholic liver disease, unspecified |
| K852 | Alcohol-induced acute pancreatitis |
| K860 | Alcohol-induced chronic pancreatitis |
| O354 | Maternal care for (suspected) damage to fetus from alcohol |
| R780 | Finding of alcohol in blood |
| T51 | Toxic effect of alcohol |
| T510 | Toxic effect: Ethanol |
| T511 | Toxic effect: Methanol |
| T512 | Toxic effect: 2-Propanol |
| T513 | Toxic effect: Fusel oil |
| T518 | Toxic effect: Other alcohols |
| T519 | Toxic effect: Alcohol, unspecified |
| X45 | Accidental poisoning by and exposure to alcohol |
| X450 | Accidental poisoning by and exposure to alcohol |
| X451 | Accidental poisoning by and exposure to alcohol |
| X452 | Accidental poisoning by and exposure to alcohol |
| X453 | Accidental poisoning by and exposure to alcohol |
| X454 | Accidental poisoning by and exposure to alcohol |

| | |
|---|---|
| X455 | Accidental poisoning by and exposure to alcohol |
| X456 | Accidental poisoning by and exposure to alcohol |
| X457 | Accidental poisoning by and exposure to alcohol |
| X458 | Accidental poisoning by and exposure to alcohol |
| X459 | Accidental poisoning by and exposure to alcohol |
| X65 | Intentional self-poisoning by and exposure to alcohol |
| X650 | Intentional self-poisoning by and exposure to alcohol |
| X651 | Intentional self-poisoning by and exposure to alcohol |
| X652 | Intentional self-poisoning by and exposure to alcohol |
| X653 | Intentional self-poisoning by and exposure to alcohol |
| X654 | Intentional self-poisoning by and exposure to alcohol |
| X655 | Intentional self-poisoning by and exposure to alcohol |
| X656 | Intentional self-poisoning by and exposure to alcohol |
| X657 | Intentional self-poisoning by and exposure to alcohol |
| X658 | Intentional self-poisoning by and exposure to alcohol |
| X659 | Intentional self-poisoning by and exposure to alcohol |
| Y15 | Poisoning by and exposure to alcohol, undetermined intent |
| Y150 | Poisoning by and exposure to alcohol, undetermined intent |
| Y151 | Poisoning by and exposure to alcohol, undetermined intent |
| Y152 | Poisoning by and exposure to alcohol, undetermined intent |
| Y153 | Poisoning by and exposure to alcohol, undetermined intent |
| Y154 | Poisoning by and exposure to alcohol, undetermined intent |
| Y155 | Poisoning by and exposure to alcohol, undetermined intent |
| Y156 | Poisoning by and exposure to alcohol, undetermined intent |
| Y157 | Poisoning by and exposure to alcohol, undetermined intent |
| Y158 | Poisoning by and exposure to alcohol, undetermined intent |
| Y159 | Poisoning by and exposure to alcohol, undetermined intent |
| Y573 | Alcohol deterrents |
| Y900 | Blood alcohol level of less than 20 mg/100 ml |
| Y901 | Blood alcohol level of 20-39 mg/100 ml |
| Y902 | Blood alcohol level of 40-59 mg/100 ml |
| Y903 | Blood alcohol level of 60-79 mg/100 ml |
| Y904 | Blood alcohol level of 80-99 mg/100 ml |
| Y905 | Blood alcohol level of 100-119 mg/100 ml |
| Y906 | Blood alcohol level of 120-199 mg/100 ml |
| Y907 | Blood alcohol level of 200-239 mg/100 ml |
| Y908 | Blood alcohol level of 240 mg/100 ml or more |
| Y909 | Presence of alcohol in blood, level not specified |
| Y910 | Mild alcohol intoxication |
| Y911 | Moderate alcohol intoxication |
| Y912 | Severe alcohol intoxication |
| Y913 | Very severe alcohol intoxication |
| Y919 | Alcohol involvement, not otherwise specified |
| Z502 | Alcohol rehabilitation |

| | |
|---|---|
| Z714 | Alcohol abuse counselling and surveillance |
| Z721 | Alcohol use |

### Table 2: Alcohol related READ codes

| READ codes | Descriptions |
|---|---|
| 136.. | Alcohol consumption |
| 1362. | Trivial drinker - <1u/day |
| 1363. | Light drinker - 1-2u/day |
| 1364. | Moderate drinker - 3-6u/day |
| 1365. | Heavy drinker - 7-9u/day |
| 1366. | Very heavy drinker - >9u/day |
| 1368. | Alcohol consumption unknown |
| 1369. | Suspect alcohol abuse - denied |
| 136F. | Spirit drinker |
| 136G. | Beer drinker |
| 136H. | Drinks beer and spirits |
| 136I. | Drinks wine |
| 136J. | Social drinker |
| 136K. | Alcohol intake above recommended sensible limits |
| 136L. | Alcohol intake within recommended sensible limits |
| 136N. | Light drinker |
| 136O. | Moderate drinker |
| 136P. | Heavy drinker |
| 136Q. | Very heavy drinker |
| 136R. | Binge drinker |
| 136S. | Hazardous alcohol use |
| 136T. | Harmful alcohol use |
| 136V. | Alcohol units per week |
| 136W. | Alcohol misuse |
| 136X. | Alcohol units consumed on heaviest drinking day |
| 136Y. | Drinks in morning to get rid of hangover |
| 136Z. | Alcohol consumption NOS |
| 136a. | Increasing risk drinking |
| 136b. | Feels should cut down drinking |
| 136c. | Higher risk drinking |
| 136d. | Lower risk drinking |
| 136e. | Declines to state current alcohol consumption |
| 13Y8. | Alcoholics anonymous |
| 13ZY. | Disqualified from driving due to excess alcohol |
| 1462. | H/O: alcoholism |
| 1B1c. | Alcohol induced hallucinations |
| 1F9D. | Replaces meals with drinks |
| 2126C | Alcohol dependence resolved |

| | |
|---|---|
| 2577. | O/E - breath - alcohol smell |
| 388u. | Fast alcohol screening test |
| 38D2. | Single alcohol screening questionnaire |
| 38D3. | Alcohol use disorders identification test |
| 38D4. | Alcohol use disorder identification test consumption questionnaire |
| 38D5. | Alcohol use disorder identification test Piccinelli consumption questionnaire |
| 38Df. | Five-shot questionnaire on heavy drinking |
| 38Dz. | Severity of alcohol dependence questionnaire |
| 38P03 | Health of the Nation Outcome Scale for Children and Adolescents item 4 - alcohol, substance/solvent misuse |
| 38QA. | CIWA-Ar - Clinical Institute Withdrawal Assessment for Alcohol scale, revised |
| 38QE. | Addiction Research Foundation Clinical Institute Withdrawal Assessment for Alcohol |
| 44X3. | Blood ethanol level |
| 66e.. | Alcohol disorder monitoring |
| 66e0. | Alcohol abuse monitoring |
| 6792. | Health ed. - alcohol |
| 67A5. | Pregnancy alcohol advice |
| 67H0. | Lifestyle advice regarding alcohol |
| 67K6. | Cycle of change stage, alcohol |
| 6892. | Alcohol consumption screen |
| 68S.. | Alcohol consumption screen |
| 7P221 | Delivery of rehabilitation for alcohol addiction |
| 8BA8. | Alcohol detoxification |
| 8BAs. | Alcohol relapse prevention |
| 8BAu. | Alcohol harm reduction programme |
| 8BAw. | Alcohol twelve step programme |
| 8CAM. | Patient advised about alcohol |
| 8CAM0 | Advised to abstain from alcohol consumption |
| 8CAv. | Advised to contact primary care alcohol worker |
| 8CE1. | Alcohol leaflet given |
| 8CdK. | Specialist alcohol treatment service signposted |
| 8G32. | Aversion therapy - alcoholism |
| 8H35. | Admitted to alcohol detoxification centre |
| 8H7p. | Referral to community alcohol team |
| 8HHe. | Referral to community drug and alcohol team |
| 8HkG. | Referral to specialist alcohol treatment service |
| 8HkJ. | Referral to alcohol brief intervention service |
| 8IA7. | Alcohol consumption screening test declined |
| 8IAF. | Brief intervention for excessive alcohol consumption declined |
| 8IAJ. | Declined referral to specialist alcohol treatment service |
| 8IAt. | Extended intervention for excessive alcohol consumption declined |
| 8IEA. | Referral to community alcohol team declined |
| 8IH4. | Alcohol Use Disorders Identification Test declined |
| 8W2.. | Referral to mental health services deferred until alcohol misuse resolved |

| | |
|---|---|
| 9EQ.. | HO/RTS-police:venesect alc |
| 9EVD. | Hospital alcohol liaison team report received |
| 9NJz. | In-house alcohol detoxification |
| 9NN2. | Under care of community alcohol team |
| 9NgzH | Withdrawn from alcohol detoxification programme |
| 9Nz9. | Emrgcy dept attn alcohl consum |
| 9NzA. | Hospital attendance related to personal alcohol consumption |
| 9k1.. | Alcohol misuse - enhanced services administration |
| 9k10. | Comm detoxification registered |
| 9k11. | Alcohol consumption counselling |
| 9k12. | Alcohol misuse - enhanced service completed |
| 9k13. | Alcohol questionnaire completed |
| 9k14. | Alcohol counselling by other agencies |
| 9k15. | Alcohol screen - alcohol use disorder identification test completed |
| 9k16. | Alcohol screen - fast alcohol screening test completed |
| 9k17. | Alcohol screen - alcohol use disorder identification test consumption questions completed |
| 9k18. | Alcohol screen - alcohol use disorder identification test Piccinelli consumption questions completed |
| 9k19. | Alcohol assessment declined - enhanced services administration |
| 9k1A. | Brief intervention for excessive alcohol consumption completed |
| 9k1B. | Extended intervention for excessive alcohol consumption completed |
| C1505 | Alcohol-induced pseudo-Cushing's syndrome |
| C253. | Wernickes encephalopathy |
| E01.. | Alcoholic psychoses |
| E010. | Alcohol withdrawal delirium |
| E011. | Alcohol amnestic syndrome |
| E0110 | Korsakov's alcoholic psychosis |
| E0111 | Korsakov's alcoholic psychosis with peripheral neuritis |
| E0112 | Wernicke-Korsakov syndrome |
| E011z | Alcohol amnestic syndrome NOS |
| E012. | Other alcoholic dementia |
| E0120 | Chronic alcoholic brain syndrome |
| E013. | Alcohol withdrawal hallucinosis |
| E014. | Pathological alcohol intoxication |
| E015. | Alcoholic paranoia |
| E01y. | Other alcoholic psychosis |
| E01y0 | Alcohol withdrawal syndrome |
| E01yz | Other alcoholic psychosis NOS |
| E01z. | Alcoholic psychosis NOS |
| E23.. | Alcohol dependence syndrome |
| E230. | Acute alcoholic intoxication in alcoholism |
| E2300 | Acute alcoholic intoxication, unspecified, in alcoholism |
| E2301 | Continuous acute alcoholic intoxication in alcoholism |
| E2302 | Episodic acute alcoholic intoxication in alcoholism |

| | |
|---|---|
| E2303 | Acute alcoholic intoxication in remission, in alcoholism |
| E230z | Acute alcoholic intoxication in alcoholism NOS |
| E231. | Chronic alcoholism |
| E2310 | Unspecified chronic alcoholism |
| E2311 | Continuous chronic alcoholism |
| E2312 | Episodic chronic alcoholism |
| E2313 | Chronic alcoholism in remission |
| E231z | Chronic alcoholism NOS |
| E23z. | Alcohol dependence syndrome NOS |
| E250. | Nondependent alcohol abuse |
| E2500 | Nondependent alcohol abuse, unspecified |
| E2501 | Nondependent alcohol abuse, continuous |
| E2502 | Nondependent alcohol abuse, episodic |
| E2503 | Nondependent alcohol abuse in remission |
| E250z | Nondependent alcohol abuse NOS |
| Eu10. | [X]Mental and behavioural disorders due to use of alcohol |
| Eu100 | [X]Mental and behavioural disorders due to use of alcohol: acute intoxication |
| Eu101 | [X]Mental and behavioural disorders due to use of alcohol: harmful use |
| Eu102 | [X]Mental and behavioural disorders due to use of alcohol: dependence syndrome |
| Eu103 | [X]Mental and behavioural disorders due to use of alcohol: withdrawal state |
| Eu104 | [X]Mental and behavioural disorders due to use of alcohol: withdrawal state with delirium |
| Eu105 | [X]Mental and behavioural disorders due to use of alcohol: psychotic disorder |
| Eu106 | [X]Mental and behavioural disorders due to use of alcohol: amnesic syndrome |
| Eu107 | [X]Mental and behavioural disorders due to use of alcohol: residual and late-onset psychotic disorder |
| Eu108 | [X]Alcohol withdrawal-induced seizure |
| Eu10y | [X]Mental and behavioural disorders due to use of alcohol: other mental and behavioural disorders |
| Eu10z | [X]Mental and behavioural disorders due to use of alcohol: unspecified mental and behavioural disorder |
| F11x0 | Cerebral degeneration due to alcoholism |
| F1440 | Cerebellar ataxia due to alcoholism |
| F25B. | Alcohol-induced epilepsy |
| F375. | Alcoholic polyneuropathy |
| F3941 | Alcoholic myopathy |
| G555. | Alcoholic cardiomyopathy |
| G8523 | Oesophageal varices in alcoholic cirrhosis of the liver |
| J153. | Alcoholic gastritis |
| J610. | Alcoholic fatty liver |
| J611. | Acute alcoholic hepatitis |
| J612. | Alcoholic cirrhosis of liver |
| J6120 | Alcoholic fibrosis and sclerosis of liver |
| J613. | Alcoholic liver damage unspecified |
| J6130 | Alcoholic hepatic failure |

| | |
|---|---|
| J617. | Alcoholic hepatitis |
| J6170 | Chronic alcoholic hepatitis |
| J6708 | Alcohol-induced acute pancreatitis |
| J6710 | Alcohol-induced chronic pancreatitis |
| L2553 | Maternal care for (suspected) damage to fetus from alcohol |
| PK80. | Fetal alcohol syndrome |
| PK83. | Fetus and newborn affected by maternal use of alcohol |
| Q0071 | Fetus or neonate affected by placental or breast transfer of alcohol |
| R103. | [D]Alcohol blood level excessive |
| SLH3. | Alcohol deterrent poisoning |
| SM0.. | Alcohol causing toxic effect |
| SM00. | Ethyl alcohol causing toxic effect |
| SM000 | Ethanol causing toxic effect |
| SM001 | Denatured alcohol-toxic effect |
| SM002 | Grain alcohol causing toxic effect |
| SM00z | Ethyl alcohol causing toxic effect NOS |
| SM01. | Methyl alcohol - toxic effect |
| SM010 | Methanol - toxic effect |
| SM011 | Wood alcohol - toxic effect |
| SM01z | Methyl alcohol-toxic eff.NOS |
| SM02. | Isopropyl alcohol-toxic effect |
| SM020 | Dimethyl carbinol-toxic effect |
| SM021 | Isopropanol - toxic effect |
| SM022 | Rubbing alcohol - toxic effect |
| SM02z | Isopropyl alcohol-tox.eff.NOS |
| SM03. | Fusel oil - toxic effect |
| SM030 | Amyl alcohol - toxic effect |
| SM031 | Butyl alcohol - toxic effect |
| SM032 | Propyl alcohol - toxic effect |
| SM03z | Fusel oil - toxic effect NOS |
| SM0y. | Other alcohol - toxic effect |
| SM0z. | Alcohol causing toxic effect NOS |
| SyuG0 | [X]Toxic eff of oth alcohols |
| T90.. | Accidental poisoning by alcohol, NEC |
| T900. | Accidental poisoning by alcoholic beverages |
| T901. | Accidental poisoning by other ethyl alcohol and its products |
| T9010 | Accid.pois.- denatured alcohol |
| T9011 | Accid.pois.- methylated spirit |
| T9012 | Accidental poisoning by grain alcohol NOS |
| T901z | Accidental poisoning by ethyl alcohol NOS |
| T902. | Accid.pois.- methyl alcohol |
| T903. | Accid.pois.- isopropyl alcohol |
| T9032 | Accid.pois.- rubbing alc.subst |
| T904. | Accid.pois.- fusel oil |

| | |
|---|---|
| T90z. | Accidental poisoning by alcohol NOS |
| TJH3. | Adverse reaction to alcohol deterrents |
| U1A9. | [X]Accidental poisoning by and exposure to alcohol |
| U1A90 | [X]Accidental poisoning by and exposure to alcohol, occurrence at home |
| U1A91 | [X]Accidental poisoning by and exposure to alcohol, occurrence in residential institution |
| U1A92 | [X]Accidental poisoning by and exposure to alcohol, occurrence at school, other institution and public administrative area |
| U1A93 | [X]Accidental poisoning by and exposure to alcohol, occurrence at sports and athletics area |
| U1A94 | [X]Accidental poisoning by and exposure to alcohol, occurrence on street and highway |
| U1A95 | [X]Accidental poisoning by and exposure to alcohol, occurrence at trade and service area |
| U1A96 | [X]Accidental poisoning by and exposure to alcohol, occurrence at industrial and construction area |
| U1A97 | [X]Accidental poisoning by and exposure to alcohol, occurrence on farm |
| U1A9y | [X]Accidental poisoning by and exposure to alcohol, occurrence at other specified place |
| U1A9z | [X]Accidental poisoning by and exposure to alcohol, occurrence at unspecified place |
| U209. | [X]Intentional self poisoning by and exposure to alcohol |
| U2090 | [X]Intentional self poisoning by and exposure to alcohol, occurrence at home |
| U2091 | [X]Intentional self poisoning by and exposure to alcohol, occurrence in residential institution |
| U2092 | [X]Intentional self poisoning by and exposure to alcohol, occurrence at school, other institution and public administrative area |
| U2093 | [X]Intentional self poisoning by and exposure to alcohol, occurrence at sports and athletics area |
| U2094 | [X]Intentional self poisoning by and exposure to alcohol, occurrence on street and highway |
| U2095 | [X]Intentional self poisoning by and exposure to alcohol, occurrence at trade and service area |
| U2096 | [X]Intentional self poisoning by and exposure to alcohol, occurrence at industrial and construction area |
| U2097 | [X]Intentional self poisoning by and exposure to alcohol, occurrence on farm |
| U209y | [X]Intentional self poisoning by and exposure to alcohol, occurrence at other specified place |
| U209z | [X]Intentional self poisoning by and exposure to alcohol, occurrence at unspecified place |
| U409. | [X]Poisoning ?intent alcohol |
| U4090 | [X]Poison ?intent alcohol home |
| U4091 | [X]Pois ?int alcohol res ins |
| U4092 | [X]Pois ?int alcohol pub inst |
| U4093 | [X]Pois ?int alcohol sport ar |
| U4094 | [X]Pois ?int alcohol on hway |
| U4095 | [X]Pois ?int alcohol trade ar |
| U4096 | [X]Pois ?int alcohol indust ar |

| | |
|---|---|
| U4097 | [X]Poisoning by and exposure to alcohol, occurrence on farm, undetermined intent |
| U409y | [X]Pois ?int alcohol OS place |
| U409z | [X]Pois ?int alcohol unsp plc |
| U60H3 | [X]Alcohol deterrents causing adverse effects in therapeutic use |
| U8... | [X]Supplementary factors related to causes of morbidity and mortality classified elsewhere |
| U80.. | [X]Evidence of alcohol involvement determined by blood alcohol level |
| U800. | [X]Evidence of alcohol involvement determined by blood alcohol level of less than 20 mg/100 ml |
| U801. | [X]Evidence of alcohol involvement determined by blood alcohol level of 20-39 mg/100 ml |
| U802. | [X]Evidence of alcohol involvement determined by blood alcohol level of 40-59 mg/100 ml |
| U803. | [X]Evidence of alcohol involvement determined by blood alcohol level of 60-79 mg/100 ml |
| U804. | [X]Evidence of alcohol involvement determined by blood alcohol level of 80-99 mg/100 ml |
| U805. | [X]Evidence of alcohol involvement determined by blood alcohol level of 100-119 mg/100 ml |
| U806. | [X]Evidence of alcohol involvement determined by blood alcohol level of 120-199 mg/100 ml |
| U807. | [X]Evidence of alcohol involvement determined by blood alcohol level of 200-239 mg/100 ml |
| U808. | [X]Evidence of alcohol involvement determined by blood alcohol level of 240 mg/100 ml or more |
| U80z. | [X]Evidence of alcohol involvement determined by presence of alcohol in blood, level not specified |
| U81.. | [X]Evidence of alcohol involvement determined by level of intoxication |
| U810. | [X]Evidence of alcohol involvement determined by level of intoxication, mild alcohol intoxication |
| U811. | [X]Evidence of alcohol involvement determined by level of intoxication, moderate alcohol intoxication |
| U812. | [X]Evidence of alcohol involvement determined by level of intoxication, severe alcohol intoxication |
| U813. | [X]Evidence of alcohol involvement determined by level of intoxication, very severe alcohol intoxication |
| U814. | [X]Evidence of alcohol involvement determined by level of intoxication, alcohol involvement, not otherwise specified |
| ZV113 | [V]Personal history of alcoholism |
| ZV4KC | [V] Alcohol use |
| ZV57A | [V]Alcohol rehabilitation |
| ZV6D6 | [V]Alcohol abuse counselling and surveillance |
| ZV704 | [V]Medicolegal examination |
| ZV70L | [V]Blood-alcohol and blood-drug test |
| ZV791 | [V]Screening for alcoholism |
| du1.. | DISULFIRAM |
| du11. | DISULFIRAM 200mg tablets |
| du12. | ANTABUSE 200mg tablets |

| | |
|---|---|
| du5.. | ACAMPROSATE CALCIUM |
| du51. | ACAMPROSATE CAL 333mg e/c tabs |
| du52. | CAMPRAL EC 333mg e/c tablets |
