## Supplementary material Figure 1 for "How does the local area deprivation influence life chances for children in poverty in Wales: A record linkage cohort study"

**Supplementary Figure 1: the percentage of the children (FSM and non-FSM) who are doing well across all area level deprivation scores**

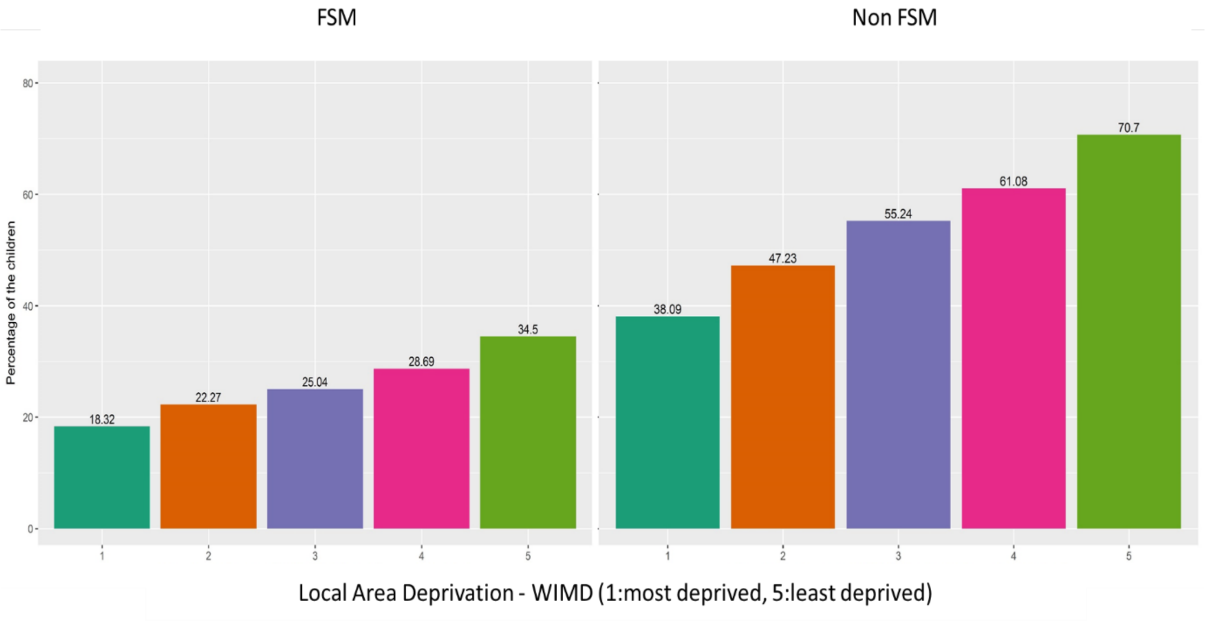
