## Supplementary material Figure 2 for "How does the local area deprivation influence life chances for children in poverty in Wales: A record linkage cohort study"

**Supplementary Figure 2: Significant factors associated with overall doing well among the FSM children**

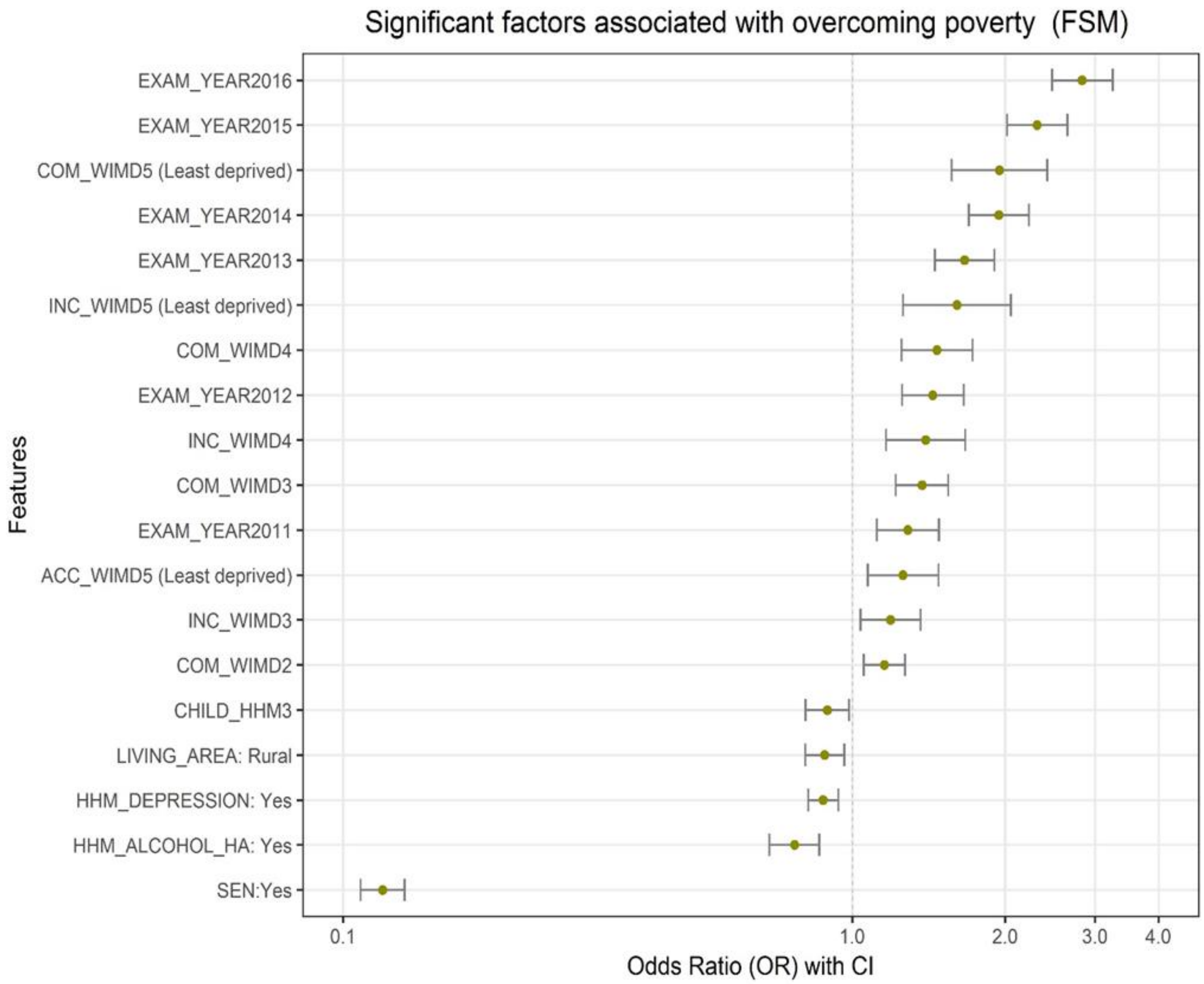


*Note: EXAM_YAER = Exam year (between 2009 and 2016), COM_WIMD = Community safety WIMD, INC_WIMD = Income WIMD, ACC_WIMD= Access to service WIMD, CHILD_HHM = Number of children in the household, LIVING_AREA = Living area, HHM_DEPRESSION = Living with someone who had depression, HHM_ALCOHOL= Living with someone who had depression, SEN = Special Education Need*
