## Supplementary material Figure 3 for "How does the local area deprivation influence life chances for children in poverty in Wales: A record linkage cohort study"

**Supplementary Figure 3: Significant factors associated with overall doing well among the non-FSM children**

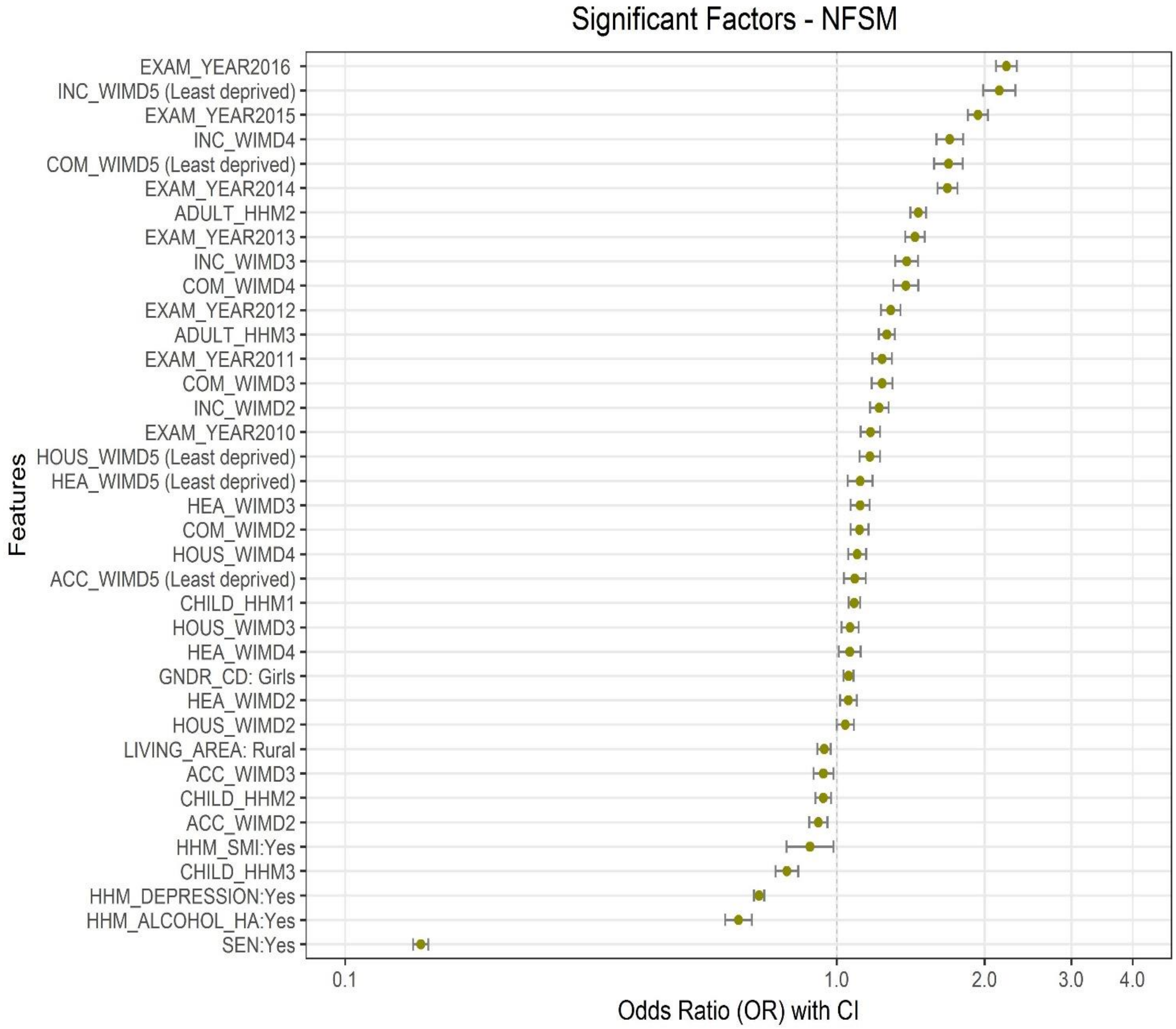


*Note: EXAM_YAER = Exam year (between 2009 and 2016), INC_WIMD = Income WIMD, COM_WIMD = Community safety WIMD, ADULT_HHM = Number of adults in the household, HOUS_WIMD = Housing WIMD, HEA_WIMD = Health WIMD, ACC_WIMD= Access to service WIMD, CHILD_HHM = Number of children in the household, GNDR_CD = Gender, LIVING_AREA = Living area, HHM_SMI = Living with someone who had serious mental illness, HHM_DEPRESSION = Living with someone who had depression, HHM_ALCOHOL= Living with someone who had depression, SEN = Special Education Need*
